# The use and impact of surveillance-based technology initiatives in inpatient and acute mental health settings: A systematic review

**DOI:** 10.1101/2024.04.04.24305329

**Authors:** Jessica L. Griffiths, Katherine R. K. Saunders, Una Foye, Anna Greenburgh, Ciara Regan, Ruth E. Cooper, Rose Powell, Ellen Thomas, Geoff Brennan, Antonio Rojas-Garcia, Brynmor Lloyd-Evans, Sonia Johnson, Alan Simpson

**Author notes:** Jessica L. Griffiths and Katherine R. K. Saunders contributed equally to this work and are listed as joint first authors.

## Abstract

**Background:** The use of surveillance technologies is becoming increasingly common in inpatient mental health settings, commonly justified as efforts to improve safety and cost-effectiveness. However, the use of these technologies has been questioned in light of limited research conducted and the sensitivities, ethical concerns and potential harms of surveillance. This systematic review aims to: 1) map how surveillance technologies have been employed in inpatient mental health settings, 2) identify any best practice guidance, 3) explore how they are experienced by patients, staff and carers, and 4) examine evidence regarding their impact.

**Methods:** We searched five academic databases (Embase, MEDLINE, PsycInfo, PubMed and Scopus), one grey literature database (HMIC) and two pre-print servers (medRxiv and PsyArXiv) to identify relevant papers published up to 18/09/2023. We also conducted backwards and forwards citation tracking and contacted experts to identify relevant literature. Quality was assessed using the Mixed Methods Appraisal Tool. Data were synthesised using a narrative approach.

**Results:** A total of 27 studies were identified as meeting the inclusion criteria. Included studies reported on CCTV/video monitoring (n = 13), Vision-Based Patient Monitoring and Management (VBPMM) (n = 6), Body Worn Cameras (BWCs) (n = 4), GPS electronic monitoring (n = 2) and wearable sensors (n = 2). Twelve papers (44.4%) were rated as low quality, five (18.5%) medium quality, and ten (37.0%) high quality. Five studies (18.5%) declared a conflict of interest. We identified minimal best practice guidance. Qualitative findings indicate that patient, staff and carer perceptions and experiences of surveillance technologies are mixed and complex. Quantitative findings regarding the impact of surveillance on outcomes such as self-harm, violence, aggression, care quality and cost-effectiveness were inconsistent or weak.

**Discussion:** There is currently insufficient evidence to suggest that surveillance technologies in inpatient mental health settings are achieving the outcomes they are employed to achieve, such as improving safety and reducing costs. The studies were generally of low methodological quality, lacked lived experience involvement, and a substantial proportion (18.5%) declared conflicts of interest. Further independent coproduced research is needed to more comprehensively evaluate the impact of surveillance technologies in inpatient settings, including harms and benefits. If surveillance technologies are to be implemented, it will be important to engage all key stakeholders in the development of policies, procedures and best practice guidance to regulate their use, with a particular emphasis on prioritising the perspectives of patients.

## Introduction

Inpatient mental health settings are challenging environments, both for those receiving and those delivering mental healthcare. The core purpose of inpatient wards is to provide a physically and psychologically safe place for people experiencing acute mental health difficulties to recover and receive care, however both patients and staff have reported feeling unsafe on wards [1,2,3]. Inpatient mental health patients report (re)traumatising experiences including abuse, coercion, aggression and violence on wards [4,5,6,7,8]. Staff also report abuse and violence on the wards [9,10], as well as having to risk-assess for and respond to incidents of self-harm and suicide attempts, which are prevalent in these settings [11]. In this context, some mental health service providers in the UK are increasing their use of surveillance-based technologies in inpatient settings [12]. Such surveillance technologies include Closed Circuit Television (CCTV), Body Worn Cameras (BWCs), and remote monitoring devices (such as smart watches, Global Positioning System (GPS) trackers and infrared cameras). Use of these technologies is justified on the basis that they may be able to detect or prevent aggressive and violent incidents, reduce self-harm incidents and suicide attempts, improve staff and patient safety, change patient behaviour and staff conduct, provide accurate records to help resolve complaints and to contribute to legal cases, and reduce staffing costs [13,14,15,16,17]. Reducing cost is a driving force for many service providers, and both conflict on wards and providing adequate staffing are costly [18] but interrelated [19,20]; surveillance technologies may therefore appear to offer a cost-effective solution.

The use of video technologies implemented with the stated purpose of improving security is becoming increasingly common. For example, in the UK, BWCs are now used by the police [21], emergency healthcare workers including paramedics [22,23,24], and retail staff [25,26,27]. However, the use of some of these technologies on inpatient wards is controversial [28,29]. Patient and service user groups, as well as advocates and disability rights activists, have consistently called for scrutiny of these technologies regarding potential risks of iatrogenic harm and ethical concerns [30,31]. For example, issues raised by the Stop Oxevision campaign include: i) ethical considerations around use of surveillance technologies and obtaining informed consent (for example, concerns about the ability of services to provide adequate information for informed consent, potential consequences for patients not providing or withdrawing consent, and whether consent can reasonably be given to being filmed or recorded while acutely unwell on an inpatient ward), ii) concerns about data access, storage, security, and human rights violations, iii) distress caused by being recorded or monitored, or the exacerbation of existing paranoia, trauma or distress [14,15,16,17], and iv) fears that it could result in reductions in staffing and one-to-one contact between staff and patients on wards.

In order to plan effective and safe mental health service delivery, it is important to determine whether evidence supports the use of surveillance technologies, and to review best practice and ethical considerations. However, a comprehensive review of the evidence underpinning the use of surveillance technologies in inpatient settings has not yet been undertaken. Therefore, we conducted, to our knowledge, the first systematic review of a range of surveillance technologies in inpatient mental health settings. Both quantitative and qualitative evidence is synthesised to answer the following overarching research question: how are surveillance-based technology initiatives being used and implemented in inpatient mental healthcare settings, and what is their impact? Our specific four research objectives were: 1a) how are surveillance-based technologies in inpatient mental health settings being implemented and what are the related implementation outcomes? 1b) what is current best practice, including the consideration of ethical issues, in the implementation of surveillance-based technologies in inpatient mental health settings? 2a) how are surveillance-based technologies in inpatient mental health settings experienced (e.g., by patients, staff, carers, visitors)? 2b) what is the effect, including benefits, harms and unintended consequences, of surveillance-based technologies in inpatient mental health settings for outcomes such as patient and staff safety and patient clinical improvement?

## Methods

We conducted a systematic review following the Preferred Reporting Items for Systematic Reviews and Meta-Analyses (PRISMA) statement [32]. The PRISMA checklist can be found in Appendix A. The protocol for our review was registered with PROSPERO (CRD42023463993). This review was conducted by the National Institute for Health and Care Research (NIHR) Policy Research Unit in Mental Health (MHPRU) based at King’s College London and University College London, which conducts research in response to policymaker need (e.g., in the Department for Health and Social Care or NHS England). Our working group met weekly, and included academic and lived experience researchers, and clinicians.

### Lived experience involvement

The working group included five lived experience researchers, who took part in all stages of the research from design, screening and extraction to analysis and write-up. The lived experience researchers included people with experience of inpatient care; conducting patient-led ward inspections; peer advocacy and support; being a carer; and direct experience of surveillance technologies during admission to inpatient mental health services. Some of the lived experience researchers were in liaison with service user groups and patients with experience of surveillance technologies. Due to the sensitive nature of the topic and related experiences, some lived experience researchers in the group have chosen to remain anonymous. Another expert by experience, who was not part of the working group, and who had direct experience of surveillance of surveillance technologies in an inpatient mental health setting, contributed only to the lived experience commentary.

### Search strategy

We searched five electronic databases (Embase, MEDLINE, PsycInfo, PubMed and Scopus) for peer-reviewed literature relevant to our research objectives. We searched for grey literature relevant to research objective 2a on a grey literature database (the Health Management Information Consortium) and two pre-print servers (medRxiv and PsyArXiv). Database searches were conducted between 17/09/2023 and 18/09/2023, with no date or language restrictions. Screening of non-English language papers was conducted using Google Translate; extraction and quality appraisal of full texts was conducted by someone with knowledge of the language. We contacted experts (including from NHS England, the Care Quality Commission, and research experts internationally) to request additional literature we may not have identified. Our lived experience networks supported the identification of additional grey literature. We also reference list screened and citation tracked included studies and relevant systematic reviews. Our search strategy included key terms relating to surveillance and inpatient mental health settings, as detailed in Appendix B.

### Screening

Title and abstract and full text screening were conducted in Rayyan [33]. Title and abstract screening was conducted by seven researchers (KS, UF, JG, AG, CR, and two NIHR MHPRU Lived Experience Researchers). 100% of titles and abstracts were independently double screened. Full text screening was conducted by nine researchers (KS, UF, JG, AG, CR, RC and three NIHR MHPRU Lived Experience Researchers). 100% of full texts were independently double screened. Any disagreements were resolved by discussion between KS, UF, JG and AG.

### Inclusion criteria

#### Participants

Mental health patients (of any age, sex, or gender), staff, carers, and visitors to services.

#### Intervention

Surveillance-based technology initiatives including CCTV, remote monitoring initiatives, smart watches, and body-worn cameras.

#### Comparators/controls

Any comparator or control group was eligible to be included.

#### Outcomes

For research objective 1a we included studies which mapped where, when, how, how often and by whom such surveillance initiatives are used and who they are used on. Information related to lived experience involvement in the development, implementation, use and evaluation of the intervention was also included, as were implementation outcomes including appropriateness, feasibility, fidelity, sustainability, penetration and costs.

For research objective 1b, we included studies which reported information relating to best practice guidelines, standards and recommendations in their results sections.

For research objective 2a, we included studies which reported qualitative data on patient, staff, and family/carer pre-implementation perceptions and post-implementation experiences of surveillance technologies.

For research objective 2b, we included quantitative data on outcomes including safety of patients, staff, carers, and visitors, use of restrictive practices and other containment measures, cost-effectiveness, care quality outcomes, clinical mental health outcomes, wellbeing, and satisfaction of patients, staff, carers, and visitors.

#### Setting

Inpatient mental health/psychiatric hospitals (including acute inpatient services, as well as longer-term rehabilitation wards and forensic wards), 136-suites and places of safety.

#### Design

We included all study designs reporting quantitative, qualitative, and mixed methods data. The exceptions are listed under ‘exclusion criteria’. For grey literature to be eligible for inclusion, the sources had to, at least briefly, describe their methodological approach.

### Exclusion criteria

We excluded conference proceedings, abstracts without an associated full text, books, PhD/MSc/BSc theses, opinion pieces, reviews, blog posts and social media content. We also excluded studies based in emergency departments, dementia-specific wards, care/nursing homes, outpatient, and drop-in crisis services. We excluded studies which focused solely on door locking, door security, or key card access practices and policies, without explicit reference to surveillance technologies. No language or location restrictions were imposed during our searches or screening.

### Data extraction

A data extraction sheet was designed in Microsoft Excel and revised based on feedback from the working group and piloting on an eligible paper by JG. The final data extraction sheet can be seen in Supplementary 1. Data extraction was conducted by eight researchers (KS, JG, UF, AG, CR, RC and two NIHR MHPRU Lived Experience Researchers). Data were independently double extracted for 4/27 (14.8%) of the included papers and an expert quantitative researcher (ARG) checked the accuracy and interpretation of all quantitative data extracted.

### Quality appraisal

As the included studies varied in design, we used the Mixed Methods Appraisal Tool (MMAT) to assess quality [34]. We also noted any additional ethical issues, the degree of lived experience involvement in the studies, and conflicts of interest reported in the papers, such as author affiliations with surveillance technology companies or funding received from them. Potential undisclosed conflicts of interest were also investigated through online searches of authors using search engines. Quality appraisal was conducted by eight researchers (KS, JG, UF, AG, CR, RC and two NIHR MHPRU Lived Experience Researchers). Independent double quality appraisal was conducted for 4/27 (14.8%) of the included papers.

### Evidence synthesis

Evidence synthesis was led by JG and UF. The interpretation of data and synthesis of results was supported by KS and the working group. Data were synthesised by research objective, and study characteristics were tabulated. Where possible, results were reported separately by type of surveillance technology.

Synthesis methods by research objective:

#### 1a) Implementation mapping and outcomes

We mapped the way the surveillance-based technologies were used in our settings of interest by technology type, including details (where available) on where, when, how often and by whom surveillance-based technologies are used and who was being surveilled. We tabulated and narratively described [35] implementation outcomes including appropriateness, adoption, feasibility, fidelity, cost, penetration, and sustainability [36].

#### 1b) Best practice

We summarised data on current best practice guidelines, standards and recommendations narratively [35].

#### 2a) Perceptions and experiences

Quantitative and qualitative data documenting perceptions and experiences of surveillance technologies were narratively synthesised [35]. We synthesised data separately according to whether perceptions and experiences were reported pre- or post-implementation of surveillance technologies. Findings were grouped into benefits and potential uses, or concerns and potential harms, and then by respondent (e.g., patients, staff, family/carers) where possible.

#### 2b) Quantitative measures of effect

Quantitative outcome data were tabulated and summarised narratively [36]. This included reporting original measures of effect (e.g., risk ratios, odds ratios, or risk differences for dichotomous outcomes, and mean differences or standardised mean differences for continuous outcomes) and p-values, where available. Results were grouped according to surveillance technology type. We were unable to perform a meta-analysis due to heterogeneity across the types of outcomes, measures of effect, populations, and length of follow up.

## Results

Figure 1 presents the PRISMA flow diagram [32] of the screening and selection process. We identified 27 studies for inclusion. Nearly half of included studies reported on CCTV/video monitoring (n = 13), other studies reported on VBPMM (n = 6), BWCs (n = 4), GPS electronic monitoring (n = 2) or wearable sensors (n = 2). Most studies were conducted in the UK (n = 18), with two conducted in Germany, one multi-country study, and one each conducted in Ireland, Malaysia, Finland, Australia, Israel, and USA. Thirteen (48.1%) studies were quantitative in design, seven (25.9%) qualitative, and seven (25.9%) mixed methods. Most studies reported data from a mix of ward types (n = 8), followed by acute wards (n = 6), low/medium secure wards (n = 5), forensic wards (n = 5) and psychiatric intensive care units (PICUs) (n = 3). Only two studies specified that they included wards with inpatients under the age of 18 [48, 63]. The remaining studies either exclusively focused on inpatient wards for adults or did not specify the age of the inpatient populations.

**Figure 1.**
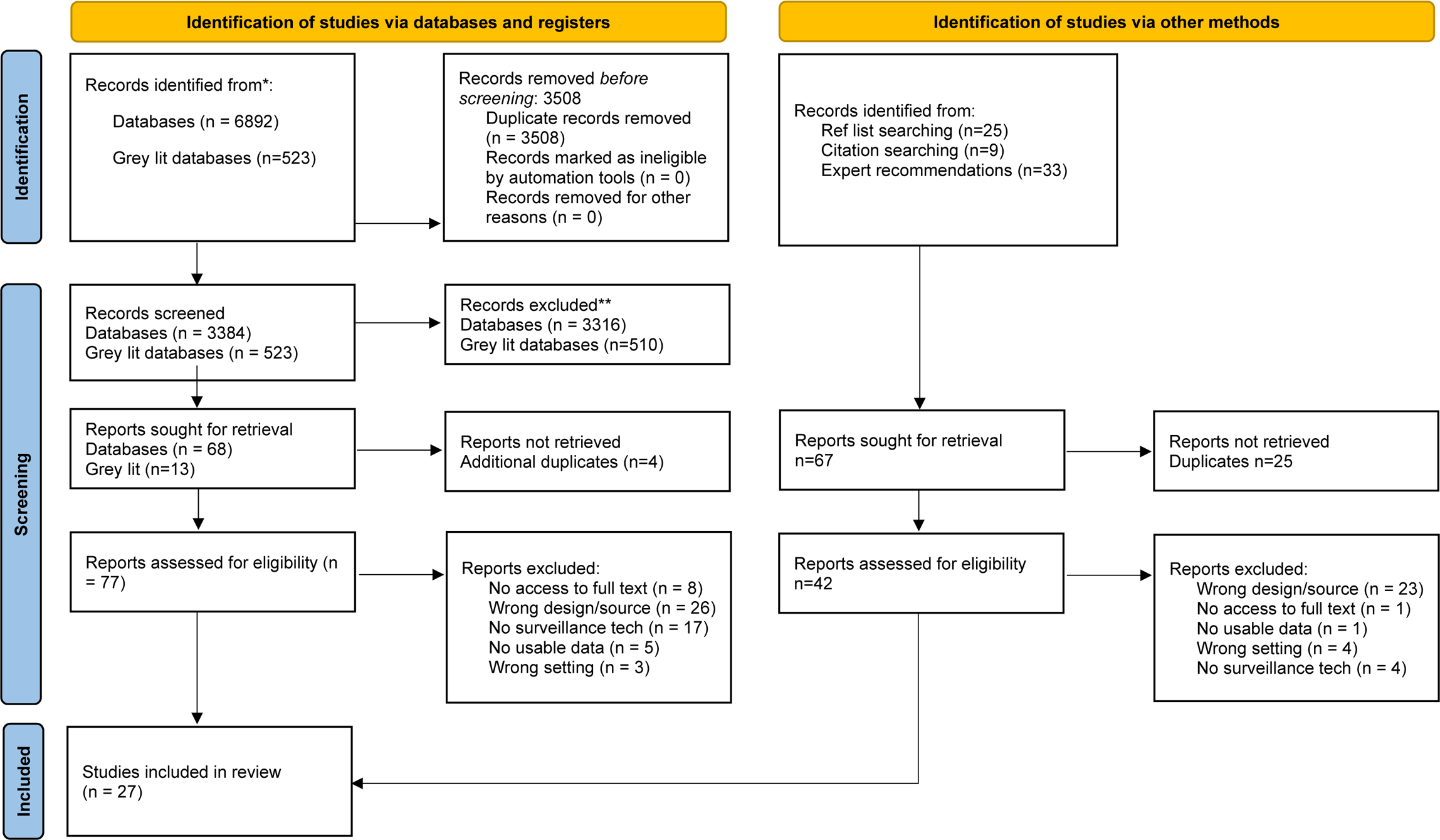
PRISMA 2020 flow diagram for new systematic reviews which included searches of databases, registers and other sources.

Twelve papers (44.4%) were rated as low quality, five studies (18.5%) were rated as medium quality, and ten studies (37.0%) were rated as high quality. For full details on MMAT ratings, see Supplementary 2. Five papers (18.5%) disclosed conflicts of interest. One report produced by a surveillance technology company [53], while other conflicts of interest included the project being funded by a surveillance technology company [40,49], authors’ time being funded by a technology company [44,51] or authors working for a surveillance technology company [40,51]. Out of the 27 studies included in this review, we also identified potential undeclared conflicts of interest in two studies. Study characteristics, including quality ratings, are summarised in Table 1. A more detailed version of this table is provided in Appendix D.

**Table 1.**
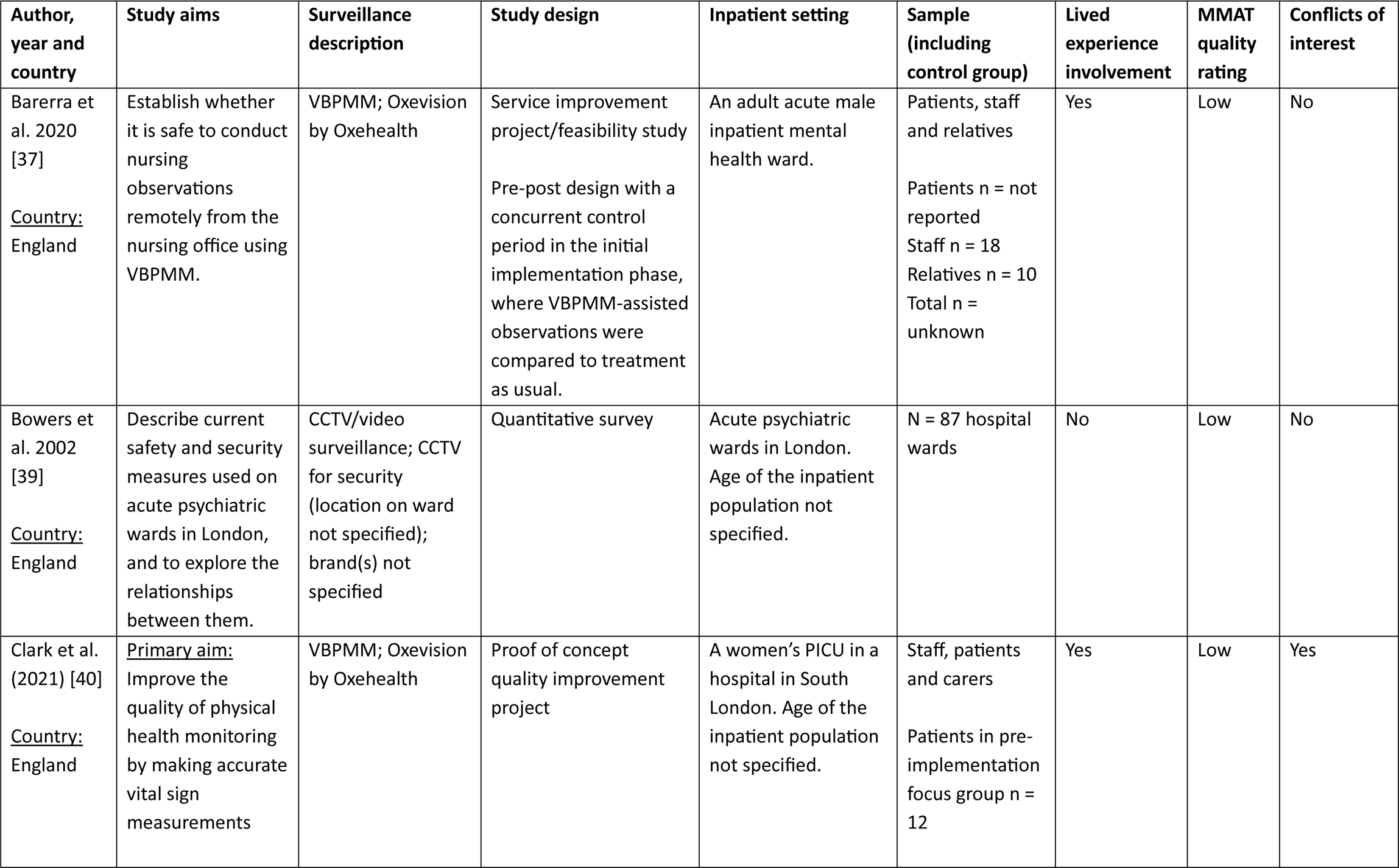
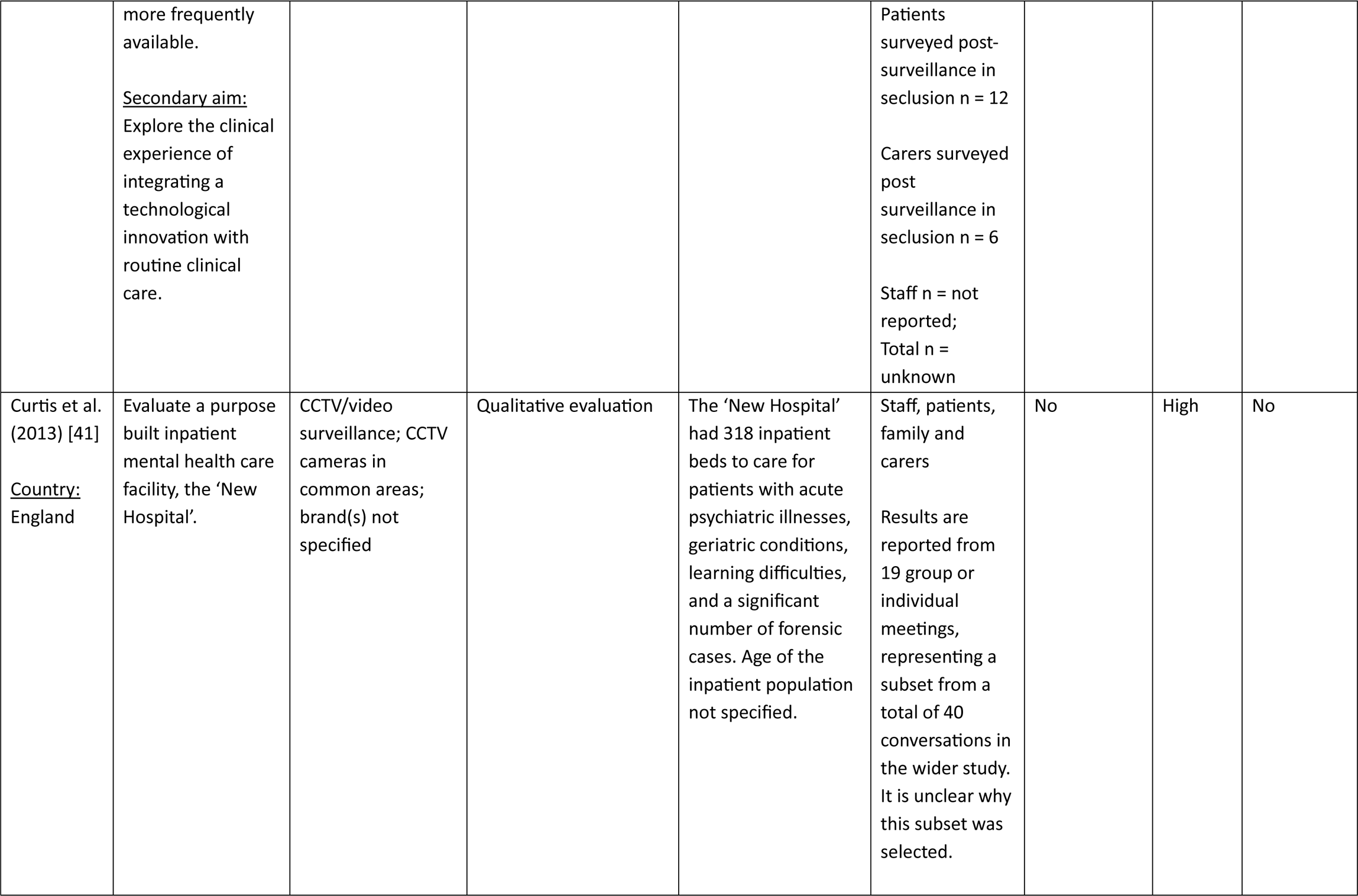
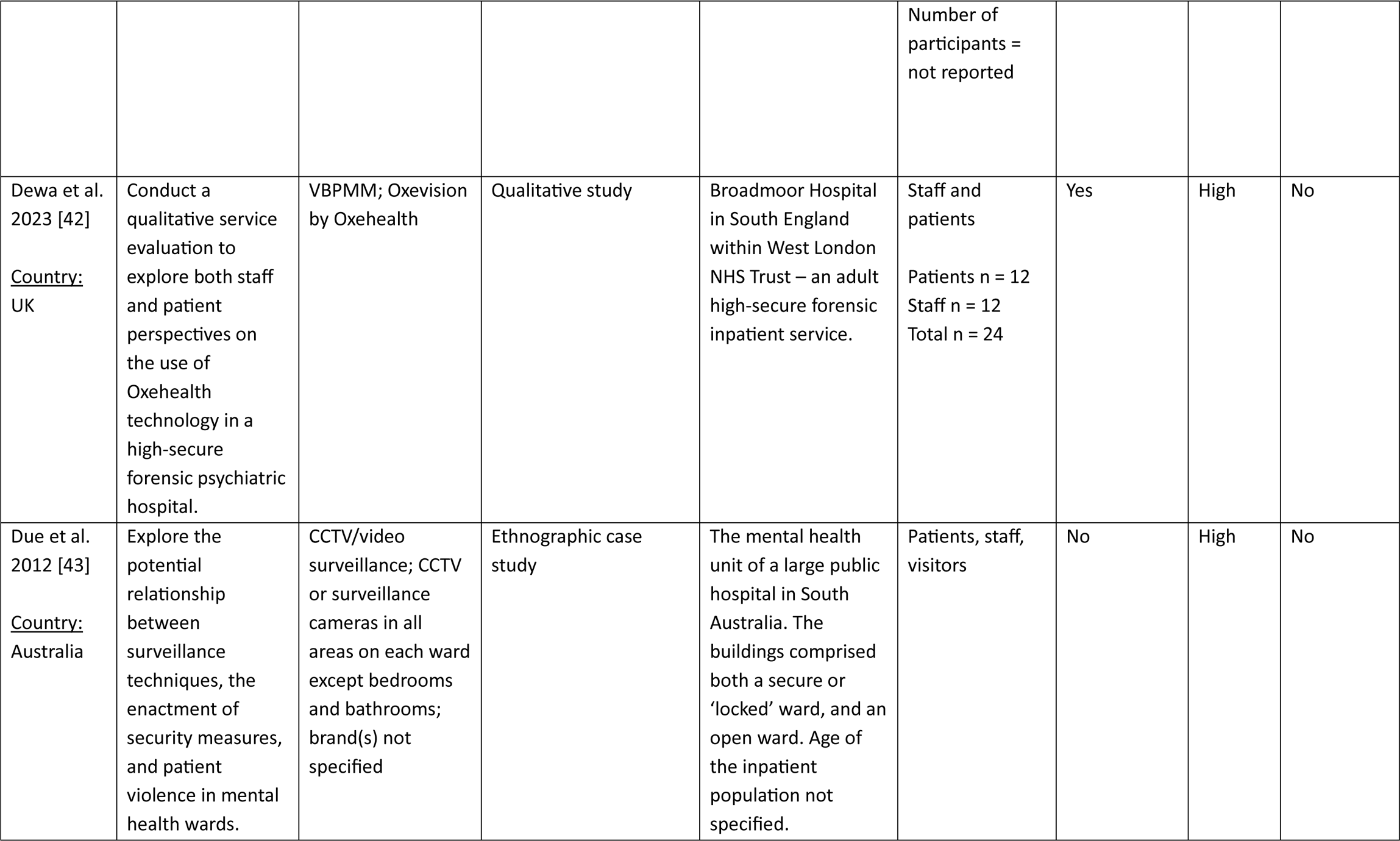
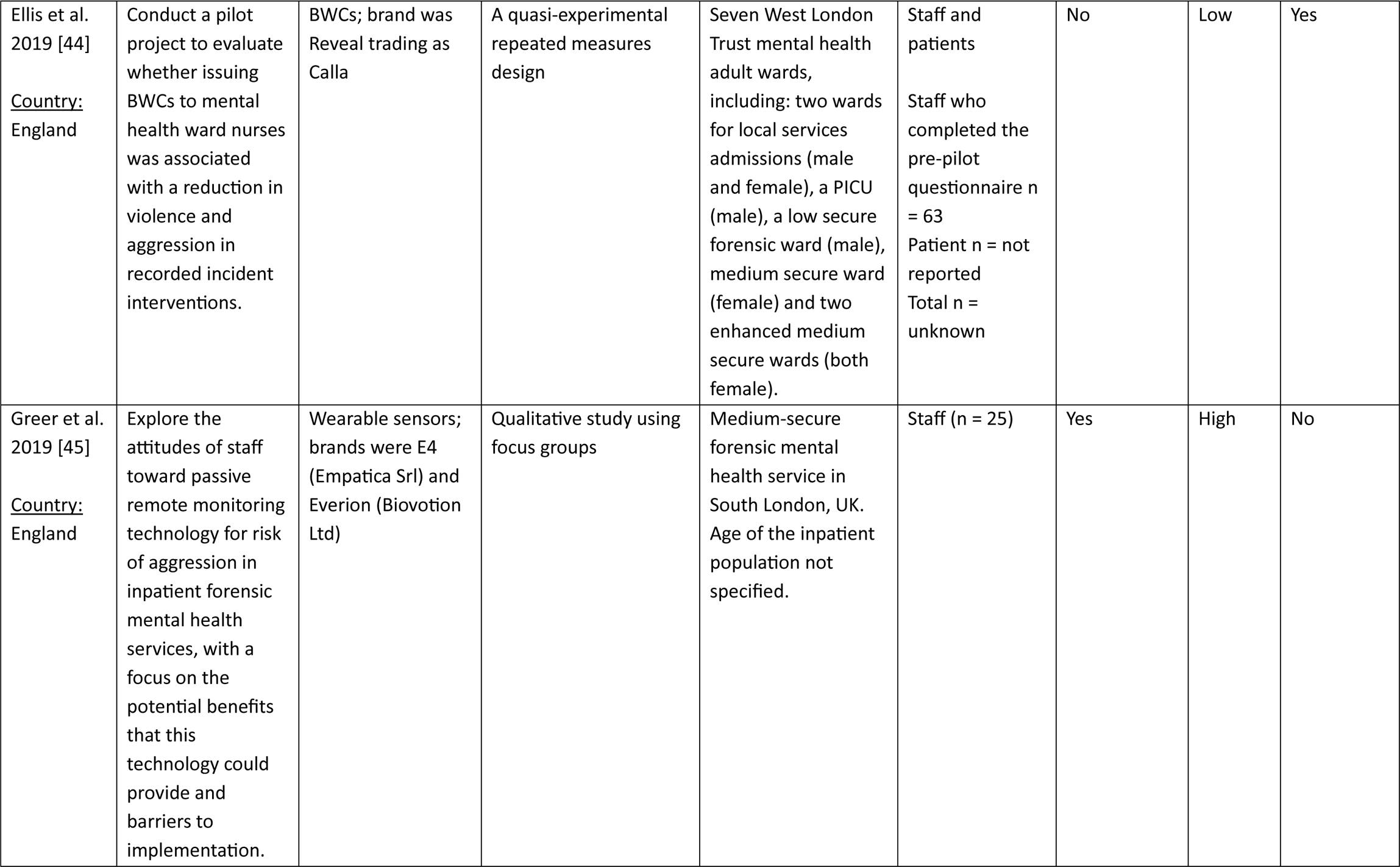
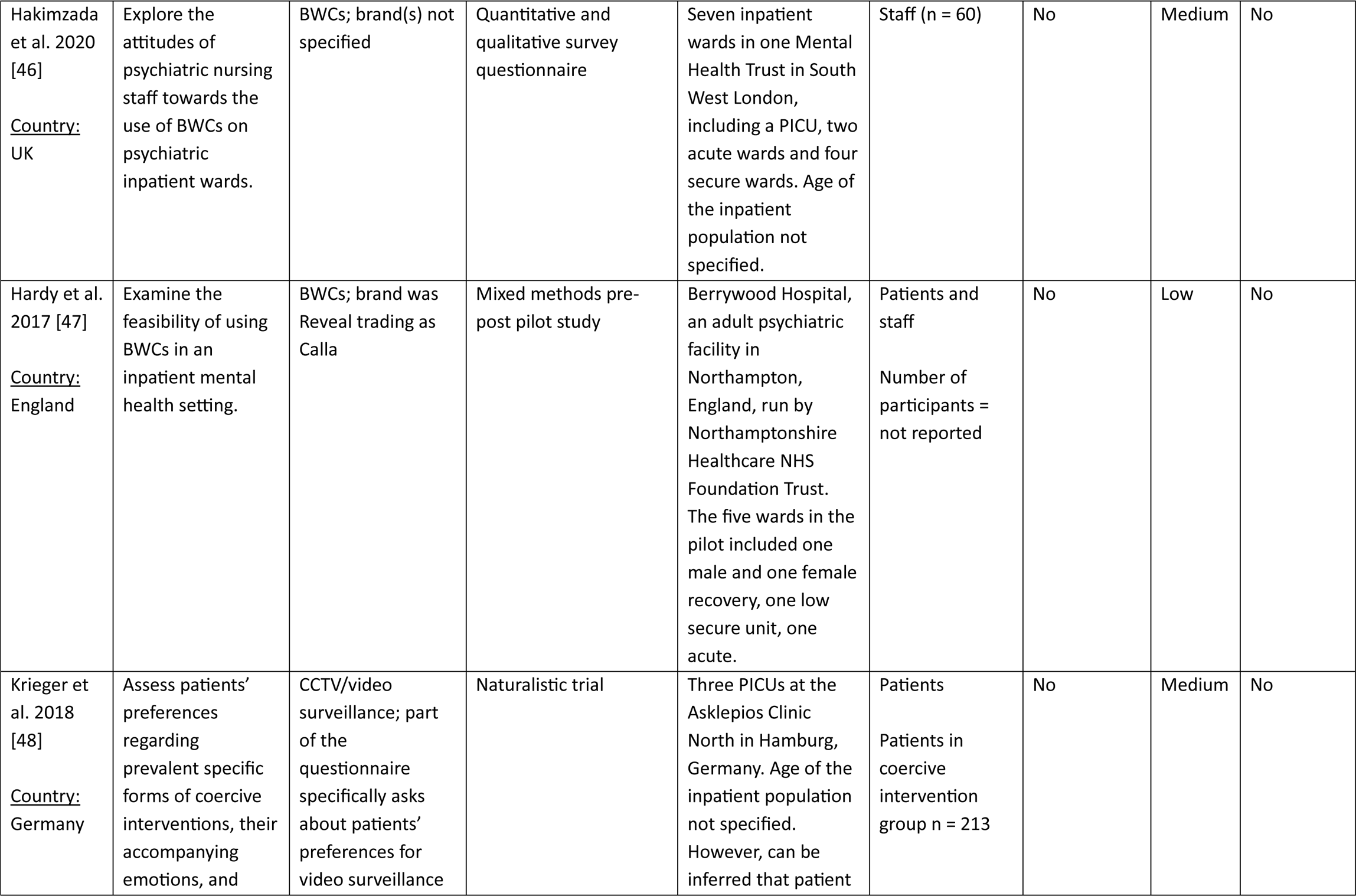
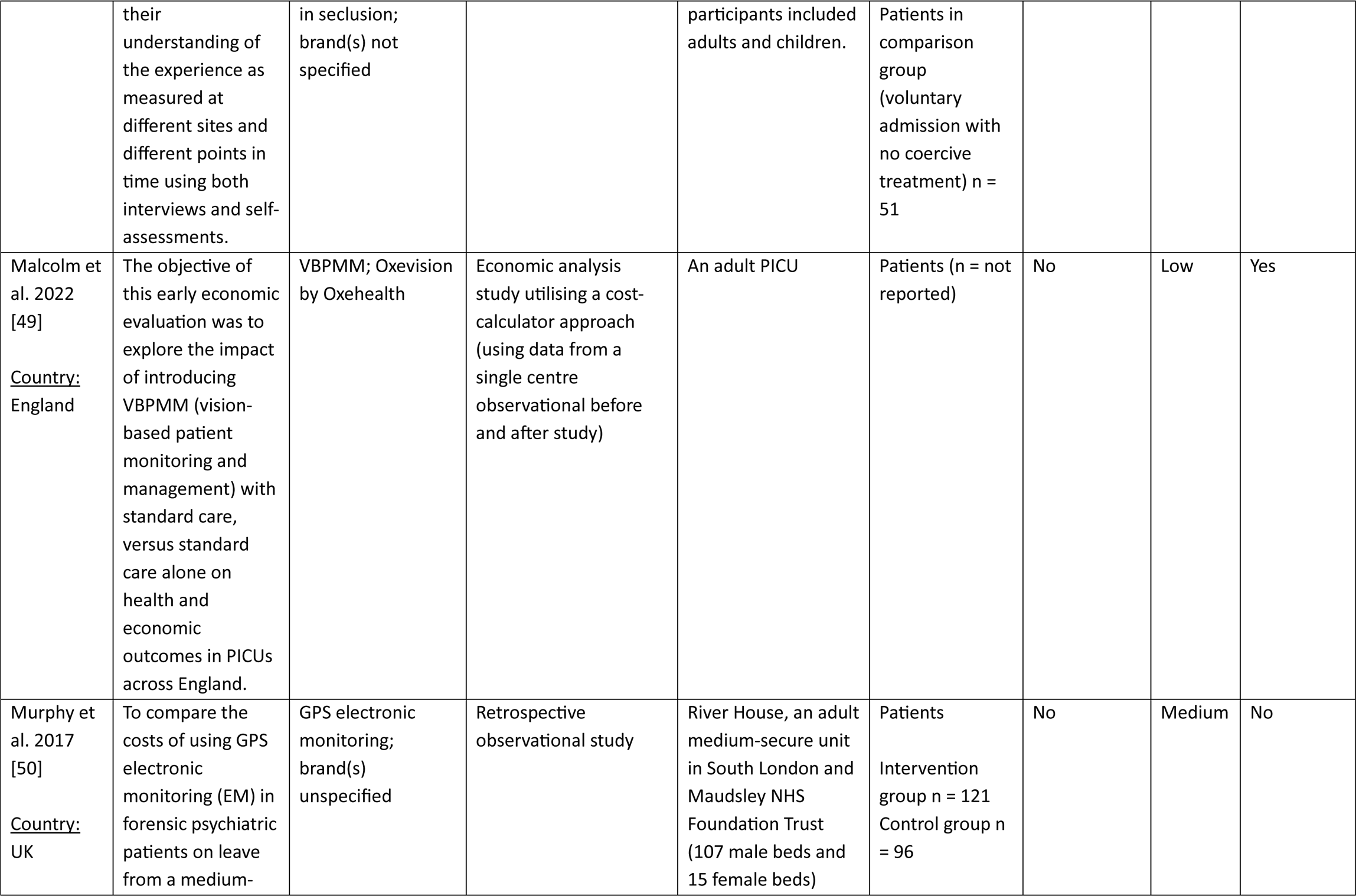
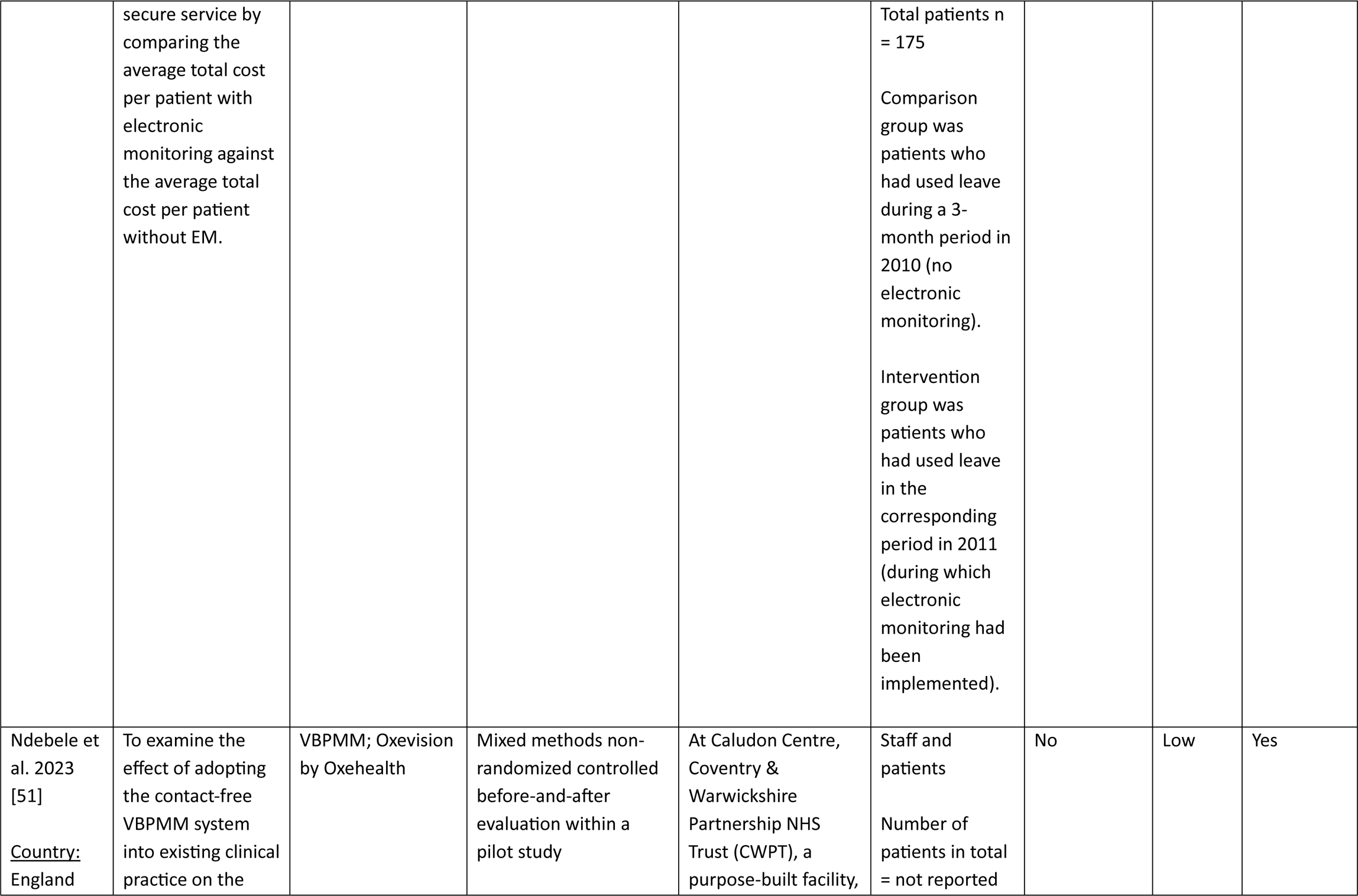
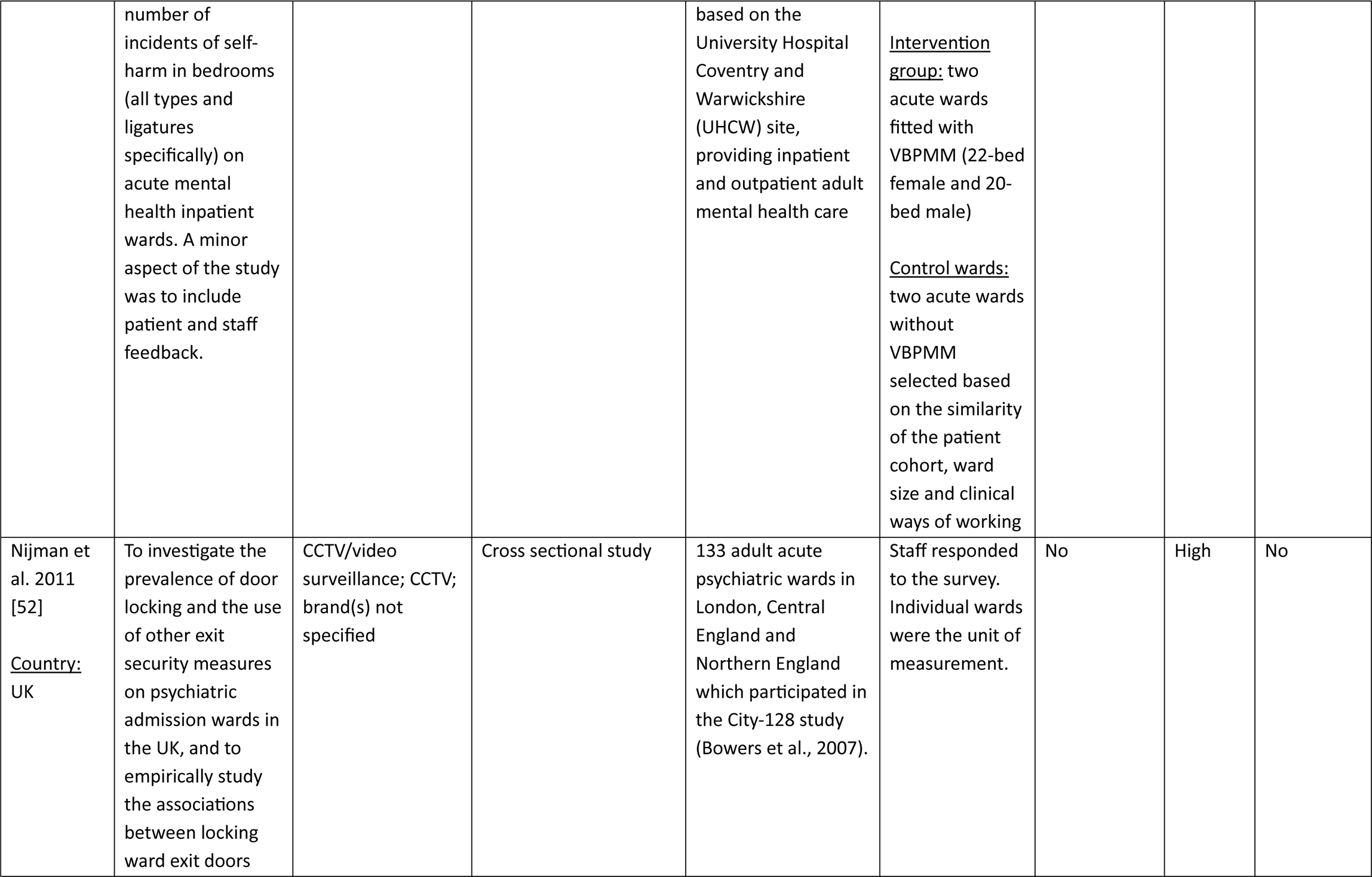
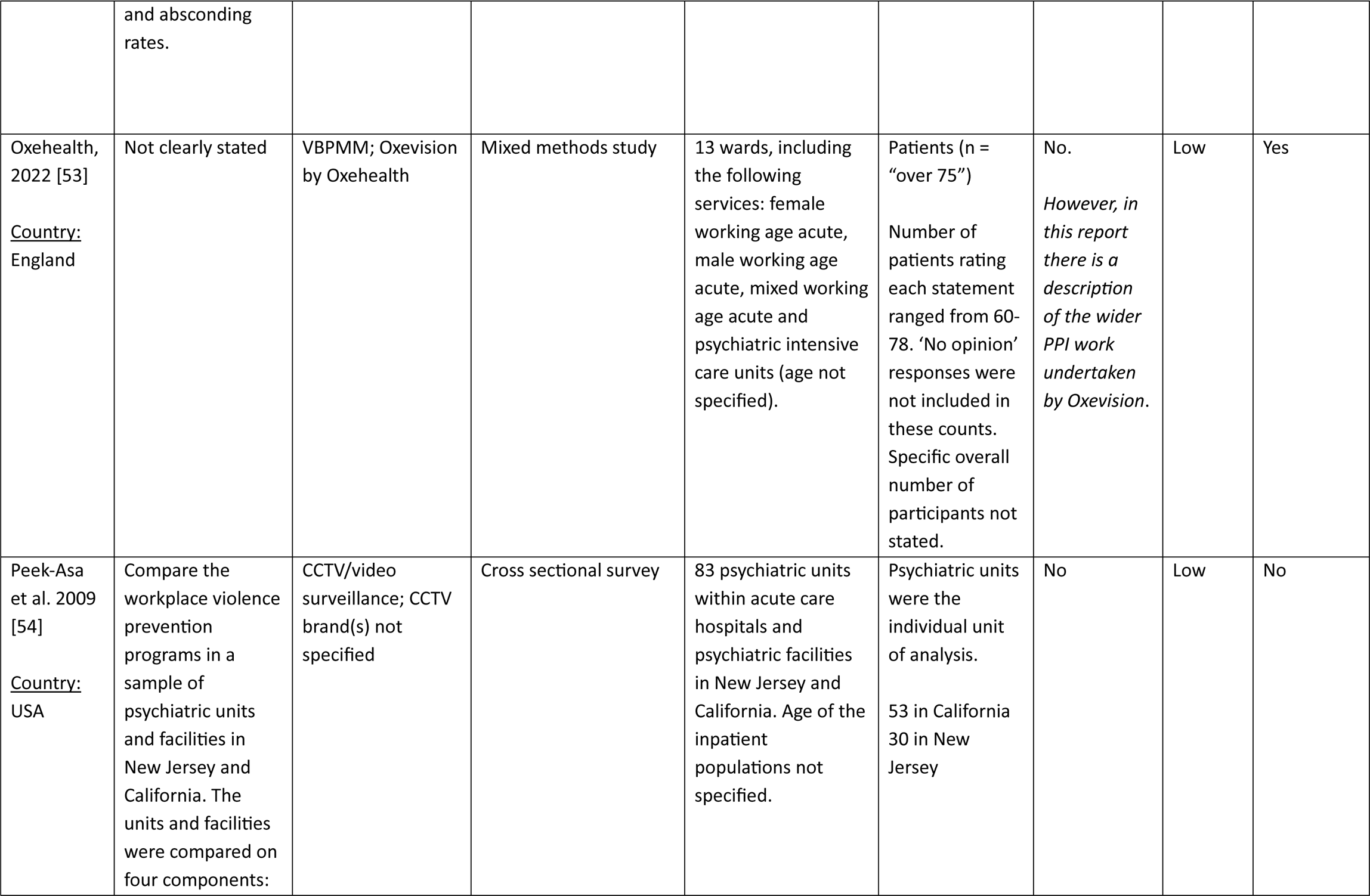
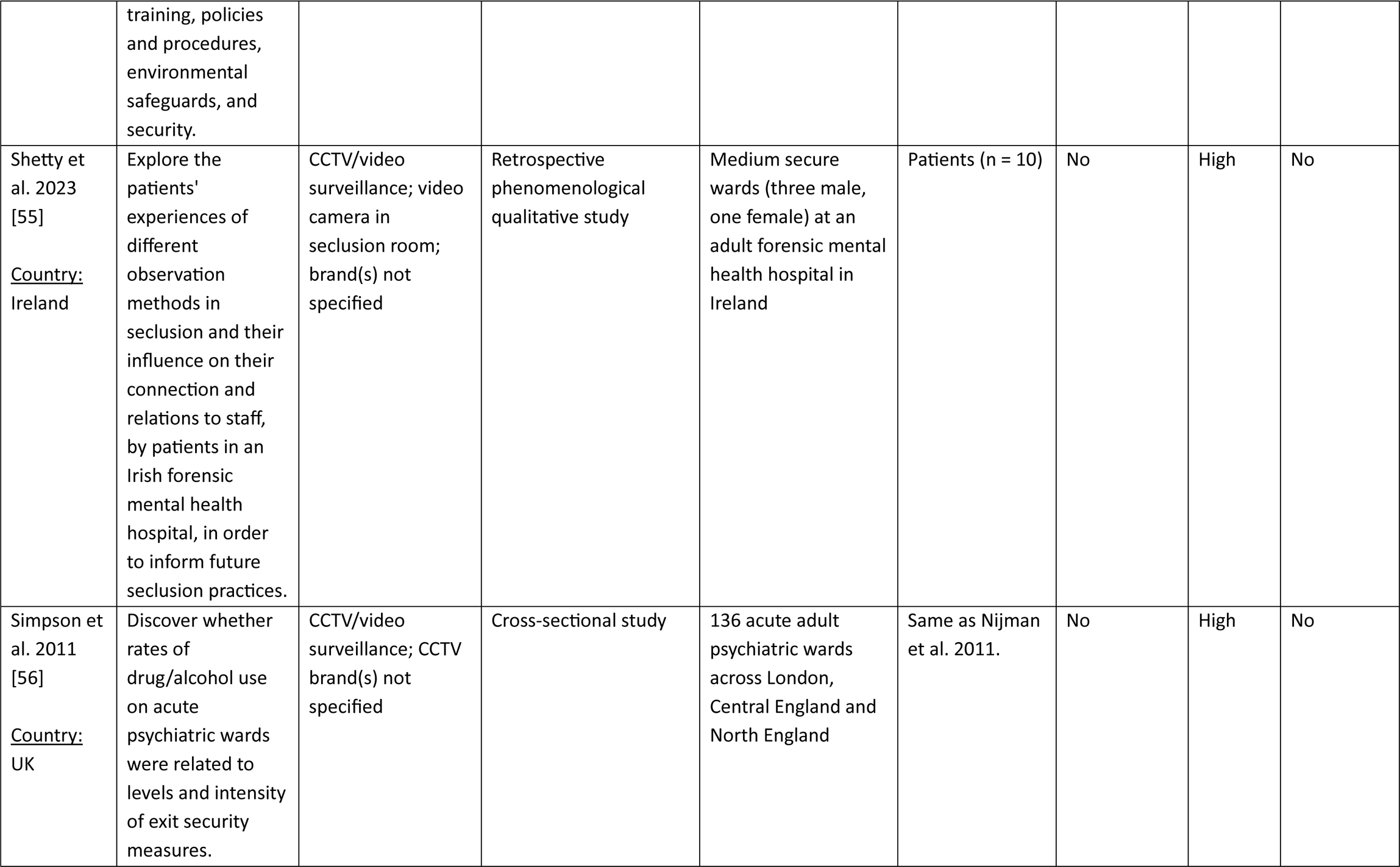
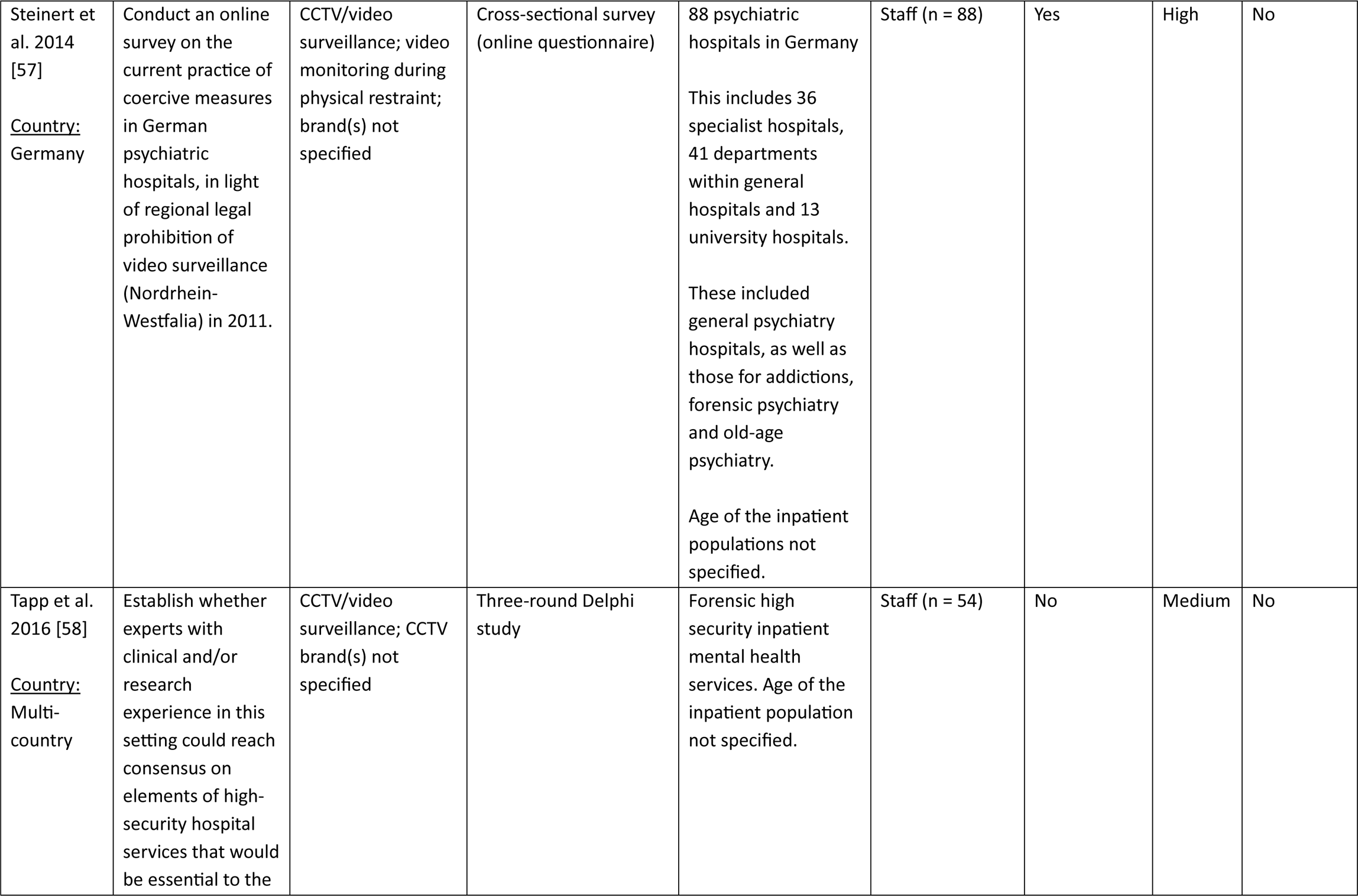
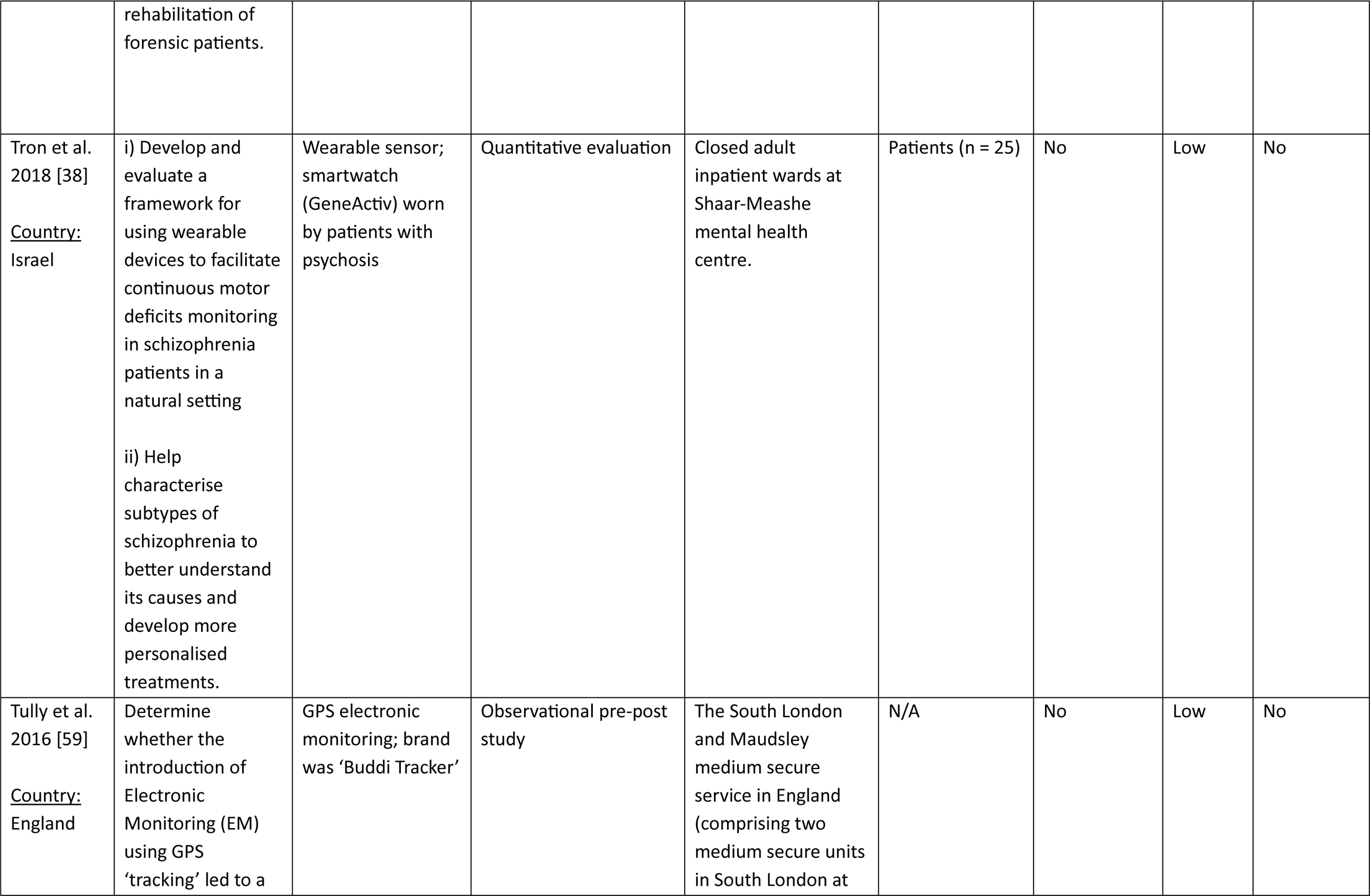
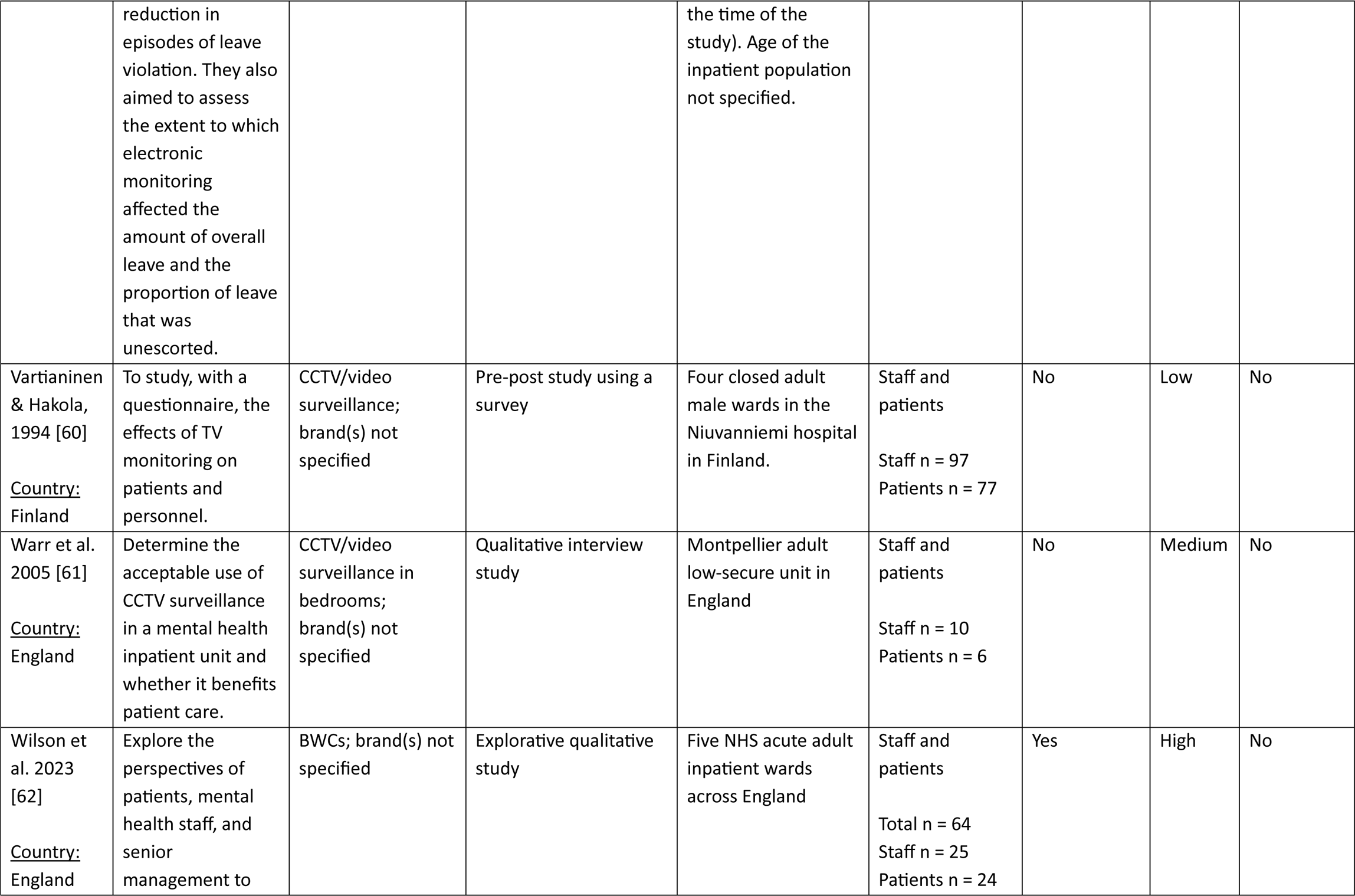
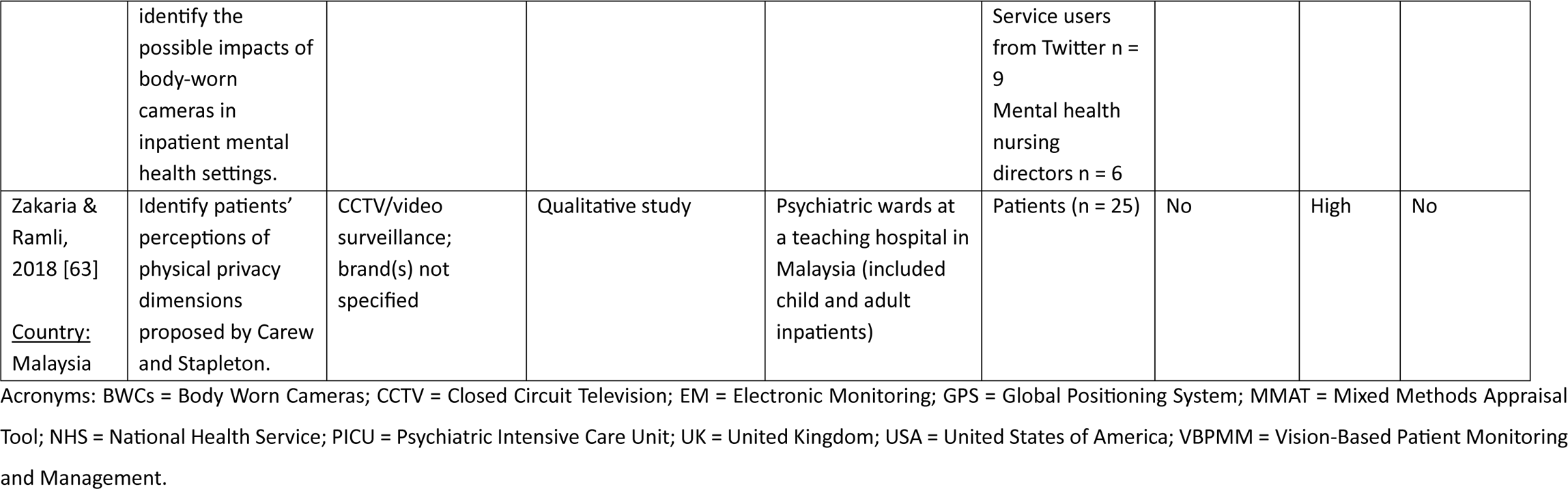
Table of study characteristics.

### Research objective 1a: How are surveillance-based technologies in inpatient mental health settings being implemented and what are the related implementation outcomes?

Below we have summarised how surveillance technologies have been implemented, and reported implementation outcomes, by type of surveillance technology. Full details on implementation process, setting, informed consent procedures and lived experience involvement can be found in Appendix E while implementation outcomes can be found in Table 2.

**Table 2.** Summary of implementation outcomes (appropriateness, feasibility, fidelity, adoption, sustainability, penetration) across the surveillance technologies.

| Surveillance technology | Implementation outcome | Results | MMAT quality rating | Conflicts of interest |
| --- | --- | --- | --- | --- |
| Vision Based Patient Monitoring and Management (VBPM) | Feasibility (n = 1 paper) | • **Ndebele et al. [51]:** Oxevision consent rates were 67% for the female acute ward, and 76% for the male acute ward. [51] | Low | Yes |
|  | Fidelity (n = 1 paper) | **Barerra et al. [37]:**<br>• There were no significant gaps or drops in the use of Oxevision during the four-week evaluation. [37]<br>• On a few nights, usage was slightly lower than expected, so some staff became ‘sensor champions’ to ensure all staff on each night shift were trained to use it. [37]<br>• During the first four night shifts, staff performed and recorded their observations as required. [37] | Low | None |
|  | Penetration (n = 1 paper) | **Barerra et al. [37]:**<br>• 17299 observations over an estimated 755 patient nights had been monitored. After 4 months, 41 patients have spent on average 14.58 (SD 14.55) nights in bedrooms with sensors (minimum of one night and maximum of 86 nights) [37]. | Low | None |
|  | Costs (n = 1 paper) | • **Malcolm et al. [49]** provided the following breakdown of costs of VBPM:<br>Standard care, Oxevision + standard care, Difference<br>Cost of night-time observational hours: £268, £158, –£109<br>Cost of one to one observation hours: £10,749, £9,943, –£806<br>Cost of assaults: £227, £167, –£60<br>Cost of rapid tranquillization event: £562, £338, –£223<br>Cost of VBPM £0, £319, £319<br>Total cost per patient £11,806, £10,926, –£880 | Low | Yes |
|  |  | Total cost per occupied bed day £246, £228, –£18<br>Total cost per average sized ward per year £970,193, £897,907, –£72,286<br>Total cost to the NHS per year £74,381,491, £68,839,567, –£5,541,924<br>This breakdown considered cost of nursing observations, assaults, rapid tranquilisations and the cost of Oxevision. Annual licence fees, installation costs and cabling costs were provided by Oxehealth. Staff training costs were calculated by combining staff costs from the Personal Social Services Research Unit with estimated staff numbers requiring training per ward provided by Oxehealth. A more detailed breakdown of costs is provided in the paper. |  |  |
| CCTV/video surveillance | Appropriateness (n = 1 paper) | • **Krieger et al. [48]:** 44% patients reported understanding why they were under video surveillance at the time, 56% reported understanding 4-5 days after. [48] | Medium | None |
|  | Adoption (n = 3 papers) | • **Nijman et al. [52]:** In a survey of 136 acute psychiatric wards in England, 27 (20%) used CCTV for monitoring who was leaving the ward.<br>• **Steinert et al. [57]:** In a survey of psychiatric hospitals in Germany, in general psychiatry and addictions, 15.9% respondents used video monitoring during mechanical restraint.<br>• **Peek-Asa et al. [54]:** “Surveillance cameras and/or mirrors” were implemented by 90.6% (48/53) of psychiatric inpatient facilities in California, and 100% (30/30) in New Jersey (p = 0.08). | 2 high;<br>1 low [54] | None |
|  | Penetration (n = 1 paper) | • **Krieger et al. [48]** found that 9.4% of patients in their current admission to one of three PICUs in Germany had been monitored via video. | Medium | None |
| Body Worn Cameras (BWCs) | Fidelity (n = 1 paper) | • **Hardy et al. [47]** reported that 68% of patients were aware some nurses were wearing BWCs. The patients who said they had not been made aware were from three of the wards, with half from one ward. | Low | None |
|  | Feasibility<br>(n = 1 paper) | • **Hardy et al. [47]:**<br>• Most (79%) of staff who did <u>not</u> wear BWCs reported observing no operational difficulties.<br>• 64% of 39 staff who <u>did</u> wear BWCs, and 69% of 23 staff who did <u>not</u> wear BWCs reported observing no practical difficulties. The remainder said they were minor and easily resolved, but 9% of staff who did <u>not</u> wear BWCs reported that the wearer needed assistance to continue to use the camera.<br>• The Trust's IT department was not asked to help with any problems during the pilot. There a few minor technical issues reported during the pilot, and these were dealt with by the clinical staff trained to be BWC administrators.<br>• No information governance concerns were raised.<br>The BWC technical/operational difficulties described included:<br>• Difficulties setting up the software<br>• Difficulties connecting to Calla's web servers<br>• Difficulties securely attaching the BWC<br>• The camera switching on if knocked<br>• Problems switching the camera on/off<br>• The camera not turning on or recording (though this was fixed quickly when reported)<br>• Difficulty wearing the harness over a coat or jacket<br>• Having to take the harness off fully to remove a fleece<br>• The harness smelling (and the wash routine to address this weakening the elastic)<br>• Staff were not taking them back to the docking station after use<br>• Staff difficulties adjusting the harness | Low | None |
|  |  | BWCs not turning on again after the first monthly generator test – they had to be disconnected and then re-docked. |  |  |
|  | Costs (n = 1 paper) | **Hardy et al. [47]:**<br>Set-up costs:<br><u>Staff costs to deliver and attend training, and to create and agree policies</u> (cost of this not specified)<br><u>IT costs:</u> The IT technician spent 48.5 hours to set up the service and deal with any problems (cost of this not specified).<br><u>Cost of cameras:</u> BWCs and related equipment were provided free of charge for this project. The costs to purchase were: camera and software £6,540; accessories £1,109.<br>Continuing costs:<br><u>Staff time to upload and review recordings</u> (3hrs/week from a senior Prevention and Management of Violence and Aggression (PMVA) team member)<br><u>Staff costs sorting out problems with cameras</u> (3hrs/week from a junior PMVA team member and 1hr/week from a senior PMVA team member)<br><u>Storage</u> (provided free of charge for this project) would have cost £569 for the 3-month period | Low | None |
| GPS electronic monitoring (EM) | Feasibility (n = 1 paper) | **Tully et al. [59]:** did not explicitly report on feasibility but stated that the technology was still in use at the time of publication. | Low | None |
|  | Fidelity (n = 1 paper) | **Tully et al. [59]:** The technology was used in the way it was intended (in the early stages of a patient being granted leave or transitioning from escorted to unescorted leave). It was<br>only used immediately before discharge in a minority of cases. No data was provided in the paper to support these claims. | Low | None |
|  | Costs (n = 2 papers) | • **Murphy et al. [50]** reported the following electronic monitoring costs:<br>• Total electronic monitoring cost over the 3-month study period for 121 devices: £34,653.<br>• Equates to an average electronic monitoring cost per patient of £286.<br>• Total cost per patient (taking into account electronic monitoring costs, staff costs, leave violation costs) was £195,703 overall in the 3-month study period (equivalent to an average of £1617 per patient).<br><u>Figures these calculations were based on:</u><br>• Hourly cost of escorting staff: £59<br>• Annual electronic monitoring contract costs: £114,336 for up to 70 devices<br>• Cost of additional devices: £119/device<br>• Leave violation costs, taking into account factors such as length of violation, whether police were contacted or involved, whether the Ministry of Justice was contacted, any media reports on local or national news, drug/alcohol use or any offences committed during leave (costs were not reported).<br>• **Tully et al. (2016):** Each GPS electronic monitoring device in this study cost £133. |  |  |
| Wearable sensors | Appropriateness (n = 1 paper) | • **Tron et al. [38]** reported that :<br>• Movement features detected by the smartwatch during the ‘free time’ window (4-5pm) were the most effective in explaining variance in patients’ scores on all factors of the Positive and Negative Syndrome Scale. | NA | NA |
|  |  | Combining data from all time windows (free time, lunch, occupational therapy, full day and full night windows) resulted in substantially higher explained variance than any of the individual windows alone for all factors.<br>They also reported a case where a patient's step count increased during a period where their medication dosage significantly changed. |  |  |

Acronyms: BWCs = Body Worn Cameras; CCTV = Closed Circuit Television; EM = Electronic Monitoring; GPS = Global Positioning System; IT = Information Technology; MMAT = Mixed Methods Appraisal Tool; PICU = Psychiatric Intensive Care Unit; PMVA = Prevention and Management of Violence and Aggression.

#### Vision-Based Patient Monitoring and Management (VBPMM)

##### Description of implementation

Six studies explored VBPMM [37,40,42,49,51,53]. All were UK-based and utilised Oxevision (a VBPMM device by Oxehealth). Four of the six studies reported conflicts of interest [40,49,51,53]. All studies were rated as low quality except one which was rated high quality [42]. This high quality study was one of the two VBPMM papers which did not report any conflicts of interest [42]. Inpatient settings included acute wards [37,51,53], psychiatric intensive care units [40,49] and a high secure forensic inpatient service [42]. VBPMM was used in patients’ bedrooms in all but one study, where it was used in a seclusion room [40].

VBPMM involves an anti-ligature, wall-mounted system equipped with an infrared-sensitive camera (a Class IIa medical device), also referred to as an ‘optical sensor’, which remotely monitors patients’ pulse and breathing rate at regular intervals [51]. It also tracks patients’ movements, generating location and activity-based alerts. Video can be viewed by staff for up to 15 seconds when taking vital sign measurements or responding to an alert. In the latter case, only blurred video is available [51]. Dewa et al. [42] states that de-pixellated video can “only be viewed with express permission in exceptional circumstances” (e.g., if there is potential risk to the patient), though it did not state who provides permission. The VBPMM system can be accessed via monitors in the nurses’ station and portable tablets. It differs from CCTV in that it has additional physical health monitoring functions and video stream viewing is intermittent ‘on-demand’ rather than continuous observation.

Ndebele et al. [51] described how consent for VBPMM use was sought from patients, or from a suitable consultee, such as their carer or the ward’s consultant psychiatrist, in cases where patients lacked capacity to consent. If consent was not given, the system remained switched off in the patient’s bedroom for the duration of their stay. If patients who lacked capacity initially later regained capacity, consent was then sought from them. The remaining papers did not describe patients being able to opt-in or out of VBPMM use.

##### Stated aims of the technology

Reported aims of VBPMM include: reducing staff disturbance to patients by enabling less intrusive observations [37,42]; allowing staff to respond to patient needs more quickly and efficiently [49], aiding monitoring of self-harm risks [51], preventing incidents [42], supporting care-planning [53]; supporting compassionate and dignified care [53] and reducing NHS mental health care costs [49]. VBPMM is reportedly intended as an adjunct to usual care, not as a replacement for therapeutic interactions or physical care [37,53]. However, it is unclear how this adjunctive role is envisioned alongside the stated aim of cost reduction.

##### Lived experience involvement in implementation

Three out of six papers reported lived experience involvement in VBPMM implementation [37,40,53]. This included a pre-implementation patient focus group [40] and meetings with former patients, relatives and nursing staff [37]. The Oxehealth report [53] stated that as an organisation, they have continuous patient and caregiver involvement throughout the implementation process. These descriptions of lived experience involvement lacked methodological detail.

##### Implementation outcomes

Three studies reported VBPMM implementation outcomes [37,49,51]. Barrera et al. [37] reported high fidelity, with no significant gaps in VBPPM use and staff observations being conducted as required, and high penetration, stating that the sensors appeared to be embedded in the ward’s day-to-day clinical practice. Ndebele et al. [49] reported VBPMM consent rates of 68% and 76% on a female and male acute ward, respectively. It was not clear whether any consenting patients later withdrew consent, and whether these figures capture those individuals. Malcolm et al. [49] compared the costs of implementing VBPMM compared to standard care. They calculated that if VBPMM were implemented in addition to standard care for adults admitted to PICUs across England, the total costs per year would be: £10,926 (GBP) per patient, £228 per occupied bed day, £897,907 per average sized ward, and £68,839,567 per year to the NHS in total. These calculations considered factors including cost of nursing observations, staff training, assaults, rapid tranquilization and the costs of the technology.

#### Closed Circuit Television (CCTV)/video surveillance

##### Description of implementation

Thirteen studies explored CCTV/video surveillance [39,41,43,48,52,54,55,56,57,58,60,61,63]. No studies declared conflicts of interest, seven studies were rated as high quality [41,43,52,55,56,57,63], three were rated medium quality [48,58,61] and three low quality [39,54,60]. These studies were based in the UK (n = 3), Germany (n = 2), Australia (n = 1), Finland (n = 1), USA (n = 1), Malaysia (n = 1) and one study recruited experts from a range of countries. CCTV/video surveillance had been implemented in acute wards [41], PICUs [48], and forensic high-secure wards [58]. Curtis et al. [41] described the setting as an inpatient psychiatric facility with beds for acute psychiatric conditions, geriatric conditions, learning difficulties and forensic cases. In six papers, the type of inpatient ward was not specified [39,43,54,57,60,63]. Within wards, CCTV was described as being implemented in communal areas (e.g., ward corridors, exit doors), patients’ bedrooms [61] and seclusion rooms [41,43,52,60]. Some specified that it was not used in private areas such as patient bedrooms [41,43] or bathrooms [43].

##### Stated aims of the technology

The functions of CCTV/video surveillance described in the papers included: monitoring patient behaviour [41,52,57,63] and staff behaviour [41]; monitoring who is leaving the ward [52], monitoring safety during mechanical restraint [57], reducing institutional incidents [58] and preventing violence [57].

##### Lived experience involvement in implementation

None reported in the included papers.

##### Implementation outcomes

Four papers reported CCTV/video surveillance implementation outcomes [48,52,54,57]. Krieger et al. [48] reported that only 44% of patients understood why they were under surveillance at the time, and only 56% understood 4-5 days after surveillance ended. Adoption rates varied between studies (from 15.9% to 100% in different locations across the USA, UK and Germany) [52,54,57]. In terms of penetration, Krieger et al. [48] reported that 9.4% patients in three PICUs in Germany had been monitored via video, though it was unclear whether all the PICUs had video surveillance technology and its location on the wards.

#### Body Worn Cameras (BWCs)

##### Description of implementation

BWCs were investigated in four UK-based studies, one of which was rated high quality [62], one medium quality [46], and two low quality [44,47]. Conflicts of interest were reported in one study [44]. In two studies, the brand was named as Calla [44,47]. Brands were not specified in the other two studies. Inpatient mental health settings included acute wards, low-secure, medium and medium enhanced forensic wards, recovery wards, and a health-based place of safety room at a psychiatric hospital. Hakimzada et al. [46] explored staff perceptions of BWCs in inpatient settings where BWCs had not been implemented, including acute wards, secure wards and a PICU.

BWCs are recording devices worn by trained staff in inpatient settings to document interactions between staff and patients via audio and video recordings. They are manually activated by staff at their discretion. This may generally be signalled by a red flashing light and audible beep, with staff advised to inform patients before recording [46]. In Hardy et al. [47], staff were trained to explain to staff and patients that the camera was for safety, to narrate their actions and intentions to the camera, and inform patients if they stop recording due to it exacerbating the situation. Staff could turn the camera around to record sound only if necessary [47].

BWC footage access was protected by a PIN to prevent data retrieval if the camera was misplaced [47]. In Hardy et al.’s [47] study, BWCs were docked, recharged and data uploaded to a secure cloud via computer in the reception area at the end of each shift. This secure cloud was provided and administered by the BWC manufacturer. Footage is kept for a fixed length of time before being automatically deleted, unless required for a specific purpose, e.g., internal investigation (Ellis et al., 2019).

Methods of informing patients of BWCs were reported in one study and included: displaying information posters in high visibility areas on wards, providing written information, and by staff verbally informing patients about them on admission, in morning meetings, patient experience groups and community meetings [47].

Hardy et al. [47] stated that preparing for BWC implementation involved establishing the necessary policies, IT infrastructure and information governance compliance – e.g., completing a full privacy impact assessment and self-assessment tool from the surveillance camera commissioner. Patients and visitors were informed, and training was delivered to staff by the BWC supplier, which was then cascaded to other ward staff. Certain staff members also received specific training to become administrators [47].

##### Stated aims of the technology

The aims of BWCs were described as: increasing transparency; resolving incidents and complaints by providing accurate incident records; improving staff performance by providing footage for training and monitoring; improving staff conduct and patient behaviour; preventing incidents of aggression; improving safety and to “counter false allegations” [44,46,47,62].

##### Lived experience involvement in implementation

None reported.

##### Implementation outcomes

One low-quality study reported BWC implementation outcomes [47]. Most staff reported no operational or practical difficulties with the BWCs. Where difficulties were reported, most were minor and easily resolved. Only 68% of surveyed patients reported that they had been made aware that some nurses were wearing BWCs [47]. Hardy et al. [47] reported the following purchase costs: camera and software (£6,540), accessories (£1,109) and storage (£569) though these were provided free by the BWC manufacturer for the study. It also provided a breakdown of staff requirements (e.g., to deliver and attend training, create policies, provide IT support, to upload and review recordings and sort out problems with cameras) but did not report the associated costs.

#### Global Positioning System (GPS) electronic monitoring

##### Description of implementation

Two low-quality papers reported on GPS electronic monitoring [50,59]. Neither reported conflicts of interest. One study used the brand Buddi Tracker [59]; the brand was unspecified in the other. Both studies were set in UK-based medium-secure inpatient mental health services.

In both studies, GPS electronic monitoring devices were attached to patients’ ankles when they went on leave. They were only used with consenting patients, with the exception of high-risk patients requiring urgent hospital or court transfer. It was unclear whether the use of GPS electronic monitoring in these instances was court-ordered or the result of a clinical decision. Consent rates were not provided in either study. Clinical decisions about the appropriateness of GPS electronic monitoring were made on an individual basis following a specific risk assessment protocol in Murphy et al. [50]. Tully et al. [59] described how it was primarily intended to be used with patients in the early stages of leave, when risk of leave violation is highest.

The ‘Buddi Tracker’ device [59] employs secure straps with anti-tamper features, transmitting location via GPS signals to monitoring software via a mobile phone network. Geographical parameters (‘geo-fences’) can be set, allowing inclusion and exclusion zones to be created. If a patient breaches a geo-fence, an alarm goes off which causes the device to vibrate and an alert to be sent through the in-built monitoring software. Information from each device is monitored by a security company. Breaches of agreed terms and conditions trigger a predetermined alert to relevant parties and a risk management plan [59].

##### Stated aims of the technology

GPS electronic monitoring tracks patients on leave, with the aim of preventing leave violations such as absconding or failing to return [50]. It was hypothesised to reduce leave violations, increase overall leave and increase the proportion of unescorted leave [50].

##### Lived experience involvement in implementation

Tully et al. [59] states that the introduction of GPS electronic monitoring was discussed with patients and legal advisors, and consent and information forms were developed. However, there is no methodological detail for patient consultation provided. No lived experience involvement in implementation was reported in Murphy et al. [50].

##### Implementation outcomes

Two papers reported GPS electronic monitoring implementation outcomes [50,59]. Though Tully et al. [59] did not directly discuss feasibility, the authors did state that the technology was still in use at the time of publication, suggesting evidence of feasibility. Tully et al. [59] also reported high fidelity; the authors claimed it was mostly used in the early stages of patients being granted leave or transitioning from escorted to unescorted leave and was only used immediately before discharge in a minority of cases. However, data were not provided to evidence this claim [59]. Murphy et al. [50] calculated that the total cost of GPS electronic monitoring over the 3-month study period was £34,653, equating to an average cost of £286 per patient. They estimated that the total cost of implementing GPS electronic monitoring over the 3-month study period, taking into account the costs of escorting staff, technology costs and leave violation costs, was an average of £1617 per patient. Tully et al. [59] simply reported that each GPS electronic monitoring device used in their study cost £133.

#### Wearable sensors

##### Description of implementation

Two papers, one rated as low quality [38] and one as high quality [45] examined wearable sensors. Neither reported conflicts of interest.

Tron et al. [38] evaluated the use of GeneActiv smart watches for monitoring movement in patients with psychosis at a psychiatric inpatient facility in Israel. These smart watches were equipped with accelerometers, light, and temperature sensors. Medical staff managed their placement and removal and uploaded data from the memory card in the device to a central storage location for analysis.

Greer et al. [45] explored staff’s perceptions of using two different remote monitoring devices to conduct real-time monitoring of patients’ psychophysiological signals to predict aggression. One device (E4, Empatica Srl) is worn around the wrist, and the other (Everion, Biovotion Ltd) is worn around the upper arm. Staff were recruited from a medium-secure forensic inpatient service in the UK and did not have prior experience with these devices.

##### Stated aims of the technology

Tron et al. [38] aimed to use the GeneActiv smartwatch to monitor patient movements and correlate them with mental states to better evaluate schizophrenia symptom severity, characterise schizophrenia subtypes and causes, and personalise treatments. In Greer et al. [45] the aim of the devices was to monitor patients’ physical indicators to predict aggression.

##### Lived experience involvement in implementation

Greer et al. [45] stated that the interview topic guide was informed by consultation with two service user–caregiver advisory groups. No lived experience involvement was reported in Tron et al. [38].

##### Implementation outcomes

Tron et al. [38] reported that movement features detected by smartwatches during the ‘free time’ window (4-5pm) were the most effective in explaining variance in patients’ scores on factors of the clinician-administered Positive and Negative Syndrome Scale (PANSS). Combining data from all time windows throughout the day resulted in substantially higher explained variance on all PANSS factors. They also reported a case where a patient’s step count increased during a period where their medication dosage changed. They argue that this evidence suggests the potential of using smartwatches for continuous tracking of schizophrenia-related symptoms and patient states in hospital settings.

### Research objective 1b: What is current best practice, including the consideration of ethical issues, in the implementation of surveillance-based technologies in inpatient mental health settings?

Only two studies explicitly reported findings on best practice and ethical considerations; neither declared a conflict of interest, one was rated medium quality [58] and the other low quality [59]. Tully et al. [59] sought legal advice before implementing GPS electronic monitoring and reported that they were advised that GPS monitoring in this study’s specific context was “legal and not in violation of human rights”. They do not provide any documentation or evidence to support this.

Tapp et al. [58] conducted a Delphi expert consensus study to try to reach consensus on the elements of high security hospital services that would be essential for the rehabilitation of forensic patients. During round one, 82% of staff and academic experts agreed that “CCTV monitoring should be implemented in the secure environment to reduce institutional incidents”, which met the 80% threshold for consensus. In round three, 62.2% of experts rated CCTV as “Important – the element of care is desirable, but its absence would not have a direct effect on the described outcome [institutional incidents]”. This did not meet the threshold for consensus, which the authors concluded meant that CCTV should not be applied prescriptively in high-secure hospital inpatient services.

#### Lived experience involvement

There was no patient or carer representation in the expert group in Tapp et al.’s [58] Delphi study, and no other lived experience involvement in this study. In Tully et al. [59], the introduction of the technology was discussed with patients and legal advisors, who helped develop consent and information forms. No further detail was provided.

### Research objective 2a – pre-implementation: How are surveillance-based technologies in inpatient mental health settings perceived pre-implementation?

#### Vision-Based Patient Monitoring and Management (VBPMM)

One study explored pre-implementation perceptions of VBPMM [40] (see Table 3 for full results). It reported a conflict of interest and was rated low quality. It reported overall positive pre-implementation staff views of VBPMM and mixed patient views. No papers reported carer views.

**Table 3.** Staff, patient and carer pre-implementation perceptions of Vision-Based Patient Monitoring and Management (VBPMM)

| Vision-Based Patient Monitoring & Management (VBPMM) |  | Pre-implementation perceptions of technology | MMAT quality rating | Conflicts of interest |
| --- | --- | --- | --- | --- |
| **Potential uses or benefits** | Staff<br>1 paper [40]<br>staff n = not reported | Staff largely felt VBMM would be a **positive addition**<br>Thought it would **help obtain vital signs** when it might otherwise be difficult to, given a patient's presentation | Low | Yes |
|  | Patients<br>1 paper [40]<br>n = 12 patients & a patient representative | Patients largely felt it would be **positively received**, as it was expected to improve **clinical safety** and **reduce disrupted sleep**<br>A separate patient representative felt it would be a **positive addition to the seclusion suite** | Low | Yes |
| **Concerns and potential harms** | Patients<br>n=1 paper [40]<br>n = 12 patients & a patient representative | One patient was concerned the camera would **emit damaging "rays"**<br>Another patient was **worried the camera would control them** in some way<br>Another patient suggested it might **reduce human interaction in seclusion** | Low | Yes |

Acronyms: MMAT = Mixed Methods Appraisal Tool; VBPM = Vision-Based Patient Monitoring and Management.

##### Staff

Staff were reported to largely feel that VBPMM could be a positive addition to seclusion rooms, as it could facilitate vital sign monitoring [40].

##### Patients

Some patients felt that VBPMM could improve safety and reduce disrupted sleep, whereas others feared that it would reduce human interaction in seclusion, or that the cameras could control or harm them [40].

#### Closed Circuit Television (CCTV)/video surveillance

Two papers explored pre-implementation perceptions of CCTV/video surveillance [41,63] (see Table 4 for full results). Neither paper reported any conflicts of interest, and both were rated high quality. Patient views were mixed, and staff and carer views were negative.

**Table 4.**
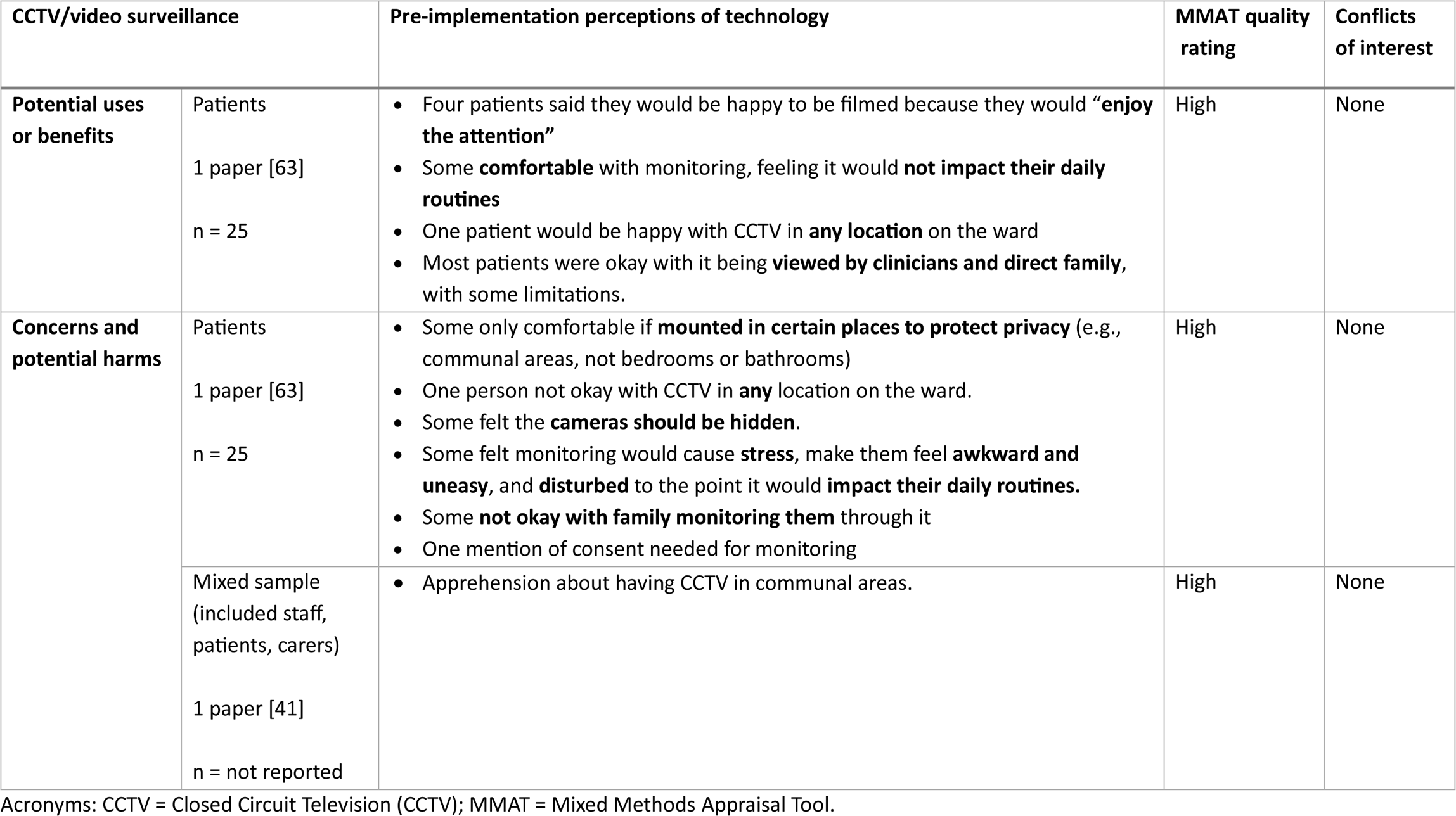
Staff, patient and carer pre-implementation perceptions of Closed Circuit Television (CCTV)/video surveillance.

##### Patients

Whilst some patients felt comfortable with the idea of being video monitored, others felt that it would cause them stress and disrupt their daily routines. Privacy concerns led some patients to prefer cameras to be positioned in communal rather than private areas. Patient preferences varied regarding camera visibility and who should be able to view the footage [63].

##### Mixed sample (patients, staff, carers)

Curtis et al. [41] reported apprehension towards the use of CCTV in communal ward spaces amongst a mixed sample of staff, patients and carers.

#### Body Worn Cameras (BWCs)

Two studies explored pre-implementation perceptions of BWCs [44,46] (see Table 5 for full results). One reported a conflict of interest [44]. One paper was rated medium quality [46] and one low quality [44]. Nursing staff views were mixed. No studies reported pre-implementation patient or family/carer views.

**Table 5.**
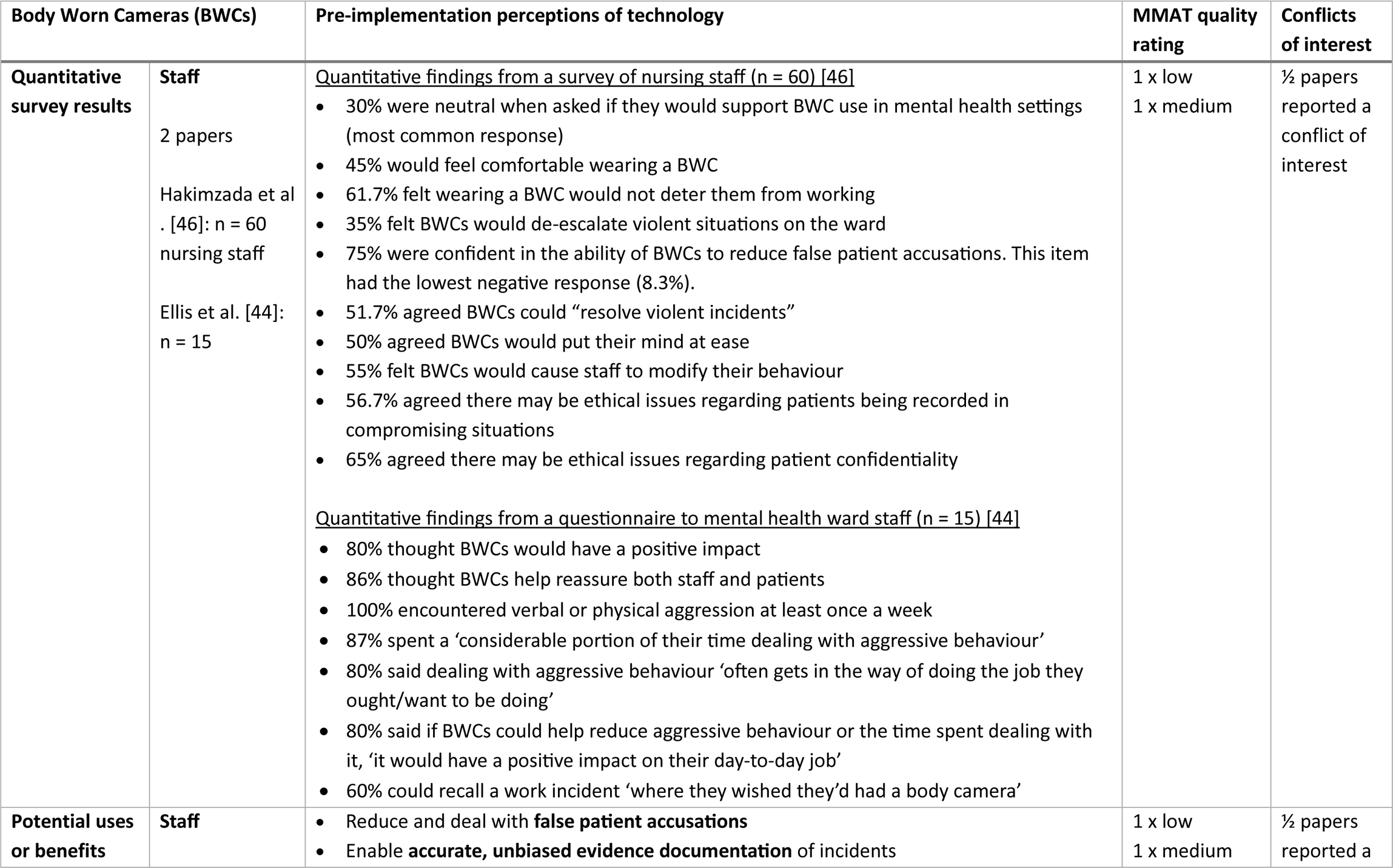
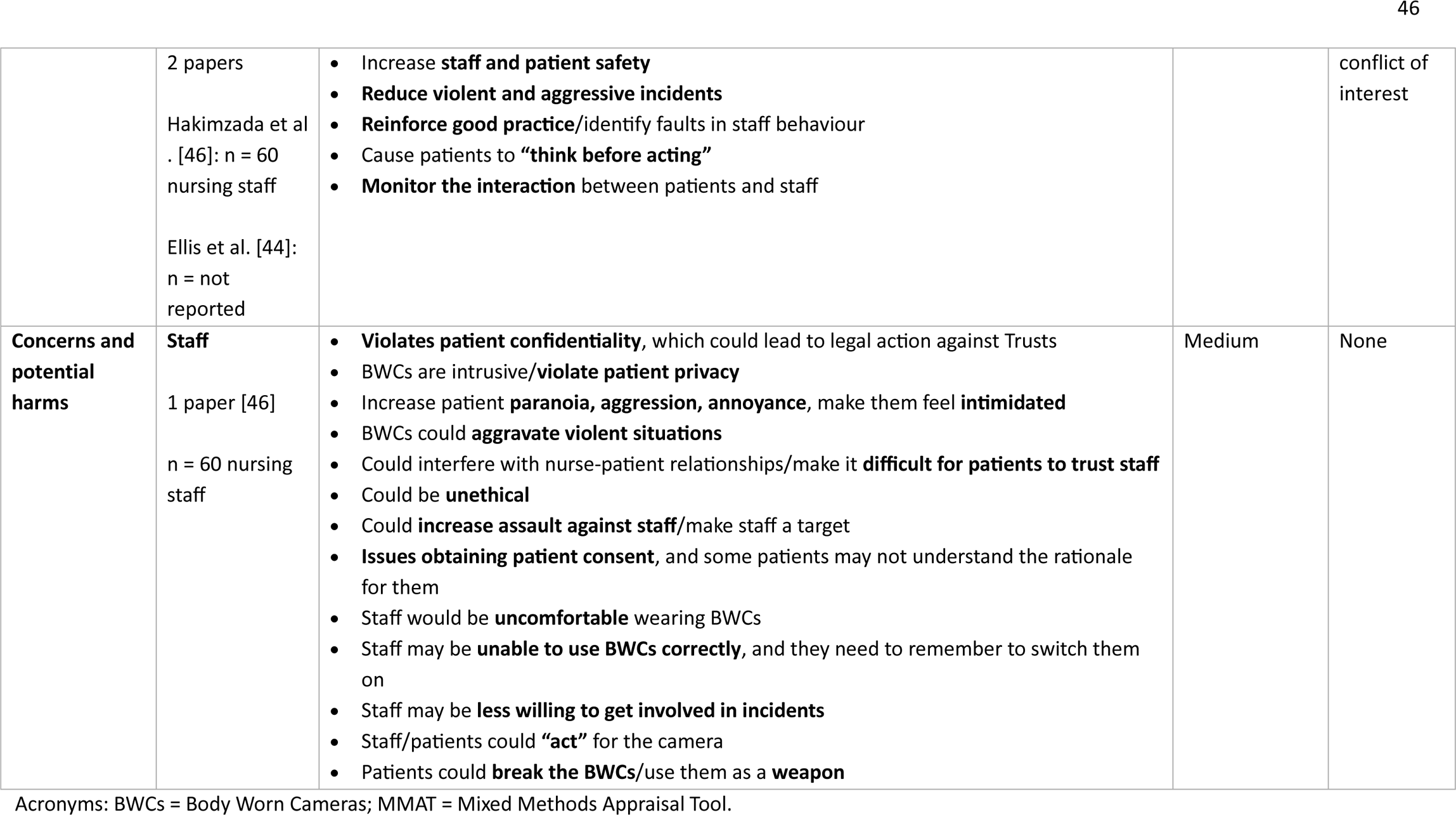
Staff, patient and carer pre-implementation perceptions of Body Worn Cameras (BWCs)

##### Staff

There were mixed views amongst nursing staff about whether they would feel comfortable wearing a BWC, whether it would deter them from working, modify staff behaviour or put their minds at ease. Some felt that BWCs could reinforce good practice and help to identify faults in staff behaviour, though others thought they may make staff less willing to get involved in incidents, or that staff and patients may “act” for the camera. Some nursing staff felt that footage from BWCs could provide accurate, unbiased documentation of incidents, and most felt that they would reduce ‘false patient accusations’. Whilst some believed that BWCs could improve staff and patient safety and help reduce and de-escalate conflict and violent incidents, and so reduce constraints on patients, others thought they could increase and exacerbate violent and aggressive situations. Some also feared that BWCs could be broken and used as weapons by patients. Furthermore, ethical concerns were raised by some staff that BWCs could violate patients’ privacy and confidentiality [44,46].

#### Global Positioning System (GPS) electronic monitoring

No papers reported on staff, patient or carer pre-implementation perceptions of GPS electronic monitoring.

#### Wearable sensors

One paper explored pre-implementation perceptions of wearable sensors [45] (see Table 6 for full results). It did not report any conflicts of interest and was rated high quality. Staff views of wearable sensors were mixed. No studies reported patient or family/carer views.

**Table 6.** Staff, patient and carer pre-implementation perceptions of wearable sensors.

| Wearable sensors |  | Pre-implementation perceptions of technology | MMAT quality rating | Conflicts of interest |
| --- | --- | --- | --- | --- |
| **Potential uses or benefits** | Staff<br>1 paper [45]<br>n = 25 nurses | • Could help **monitor risk** and so **prevent situations escalating, reducing violent incidents**<br>• Provides **information** which patients may otherwise not express or which may not be observable by staff.<br>• Could **foster self-awareness** among patients<br>• **Facilitates less obtrusive monitoring** without the need for physical contact<br>• Factors that could increase patient willingness to engage could include stylish design and having clear benefits to wearing the device (e.g., if it affected their leave status) | High | None |
| **Concerns and potential harms** | Staff<br>1 paper [45]<br>n = 25 nurses | • Device could be used as a **ligature** due to elastic armband<br>• Device could be used as a **weapon** to cause **harm to self or others**<br>• Could **exacerbate patient paranoia**<br>• Concerns about **data security and patient confidentiality**<br>• Could **add to staff's workloads** (e.g., if need to manually upload/analyse data, monitoring patient use of the technology, or if checklists accompany them).<br>• Patients' willingness to use the technology may **change depending on their mental state** | High | None |

Acronyms: MMAT = Mixed Methods Appraisal Tool.

##### Staff

Staff recognised wearable sensors’ potential for facilitating less obtrusive monitoring, increasing patients’ self-awareness and providing information that may not otherwise be shared with staff. Some also felt that they could aid risk-monitoring, reduce violent incidents and prevent situations from escalating. However, concerns included patients misusing them as ligatures or weapons, exacerbating patient paranoia, data security and patient confidentiality issues, fluctuating patient willingness to use them and increased workload for staff.

### Research objective 2a – post-implementation: How are surveillance-based technologies in inpatient mental health settings experienced post-implementation?

#### Vision-Based Patient Monitoring and Management (VBPMM)

Five papers explored post-implementation experiences of VBPMM [37,40,42,51,53] (see Table 7 for full results). Three of these studies reported conflicts of interest [40,51,53]. Four were rated low quality [37,40,51,53], one was rated high quality [42]. Experiences of patients, staff and carers were mixed.

**Table 7.** Staff, patient and carer post-implementation experiences of Vision-Based Patient Monitoring and Management (VBPMM)

| Vision-Based Patient Monitoring & Management (VBPMM) |  | Post-implementation experiences of surveillance | MMAT quality rating | Conflicts of interest |
| --- | --- | --- | --- | --- |
| **Perceived benefits** | Staff (n = 4 papers)<br>[37,40,42,51] | • Positive effect on **patients' sleep**<br>• **Observations easier and quicker** for staff<br>• Perceived **reduction in verbal and physical aggression**<br>• Perceived **improvement to patients' privacy and dignity** when compared to in person observation<br>• Technology helps **identify incidents**<br>• Leads to **better care** for patients<br>• Improved **staff and patient safety**<br>• Improved **assurance for staff managing risk**<br>• Can serve as **an extra safety measure** when staff were unable to perform physical checks on a patient (e.g., if they were behaving aggressively)<br>• Improved physical health monitoring **aiding clinical decision making** | 3 x low<br>1 x high | 2/4 papers reported a conflict of interest |
|  | Patients (n = 3 papers)<br>[40,42,53] | • **Feeling safer** as monitoring leads to staff helping quicker if their health worsens<br>• Technology **aids independence** from staff<br>• **Better nights' sleep** with remote monitoring (as physical checks disturbed sleep)<br>• Monitoring in seclusion aided **feeling connected to others**<br>• Some patients feel **indifferent** about the technology's use, for example, over time forgetting that it was there, paying less attention to it, and accepting that it was there to stay | 2 x low<br>1 x high | 2/3 papers reported a conflict of interest |
|  | Carers (n = 1 paper [40]) | • Carers **had mostly positive perceptions** of monitoring. | Low | Yes |
| **Negative impacts, effects and harms** | Staff (n = 1 paper [42]) | • **Technological glitches** (e.g., poor Wi-Fi, signal issues, poor readings of patient activity)<br>• **Security concerns; data protection** and **physical concerns** about the device e.g., concerns about patients accessing VBPMM data via the code on the back of staff's iPads<br>• **Lack of trust** in technologies **accuracy**<br>• **Insufficient training** to be able to use the technology correctly, and **issues with staff ability** to use the technology<br>• Technology **not a replacement for standard care** and physical observations | High | None |
|  |  | Negative effect on patients' **privacy** including **ethical concerns** regarding watching patients |  |  |
|  | Patients (n = 1 paper [42]) | **Lack of privacy and dignity** felt when monitored<br>Concerns regarding the **impact on human rights**<br>Feelings of **embarrassment, distress and paranoia** regarding being watched (particularly around getting undressed)<br>**Lack of choice** or say about the use of the technology<br>**Less trust in staff** and **impact on relationships** with staff<br>**Increased power imbalance** between staff and patients<br>**Lack of communication** about the technology, including **inaccuracies in explanations** | High | None |
|  | Carers (n = 1 paper [40]) | Concerns regarding the **negative impact on quality of care**<br>Negative perceptions more common amongst patients who had **spent less time in hospital** | Low | Yes |

Acronyms: MMAT = Mixed Methods Appraisal Tool; VBPM = Vision-Based Patient Monitoring and Management.

##### Staff

Benefits of VBPMM perceived by staff included improved sleep and enhanced staff and patient safety (e.g., through improved physical health monitoring and reduced patient aggression). There were mixed perspectives on its impact on patients’ privacy. Staff also flagged concerns about technological issues (e.g., poor Wi-Fi), incorrect use of the technology, insufficient staff training and doubts about its accuracy. Some felt VBPMM should not replace standard care and physical observations [37,40,42,51].

##### Patients

Some patients also felt that VBPMM improved patient safety and sleep. Other benefits reported by patients included increased independence from staff and a greater sense of connection in seclusion. However, patients also raised ethical concerns about VBPMM’s negative impact on their privacy, dignity and human rights. They cautioned about how being monitored can cause distress, exacerbate power imbalances and damage trust between patients and staff. Concerns were also raised about a lack of patient choice, and inadequate or inaccurate communication from staff regarding VBPMM [40,42,53].

##### Carers

One paper reported that carers had mostly positive perceptions of VBPMM, but some had concerns about a negative impact on care quality [40].

#### Closed Circuit Television (CCTV)/video surveillance

Five papers explored post-implementation experiences of CCTV/video surveillance [41,43,55,60,61] (see Table 8 for full results). None of these studies reported any conflicts of interest. Three were rated high quality [41,43,55], one medium quality [61] and one low quality [60]. Three studies explored experiences of CCTV/video surveillance in communal ward areas [41,43,60], one in a seclusion room [55], and one in patients’ bedrooms [61].

**Table 8.**
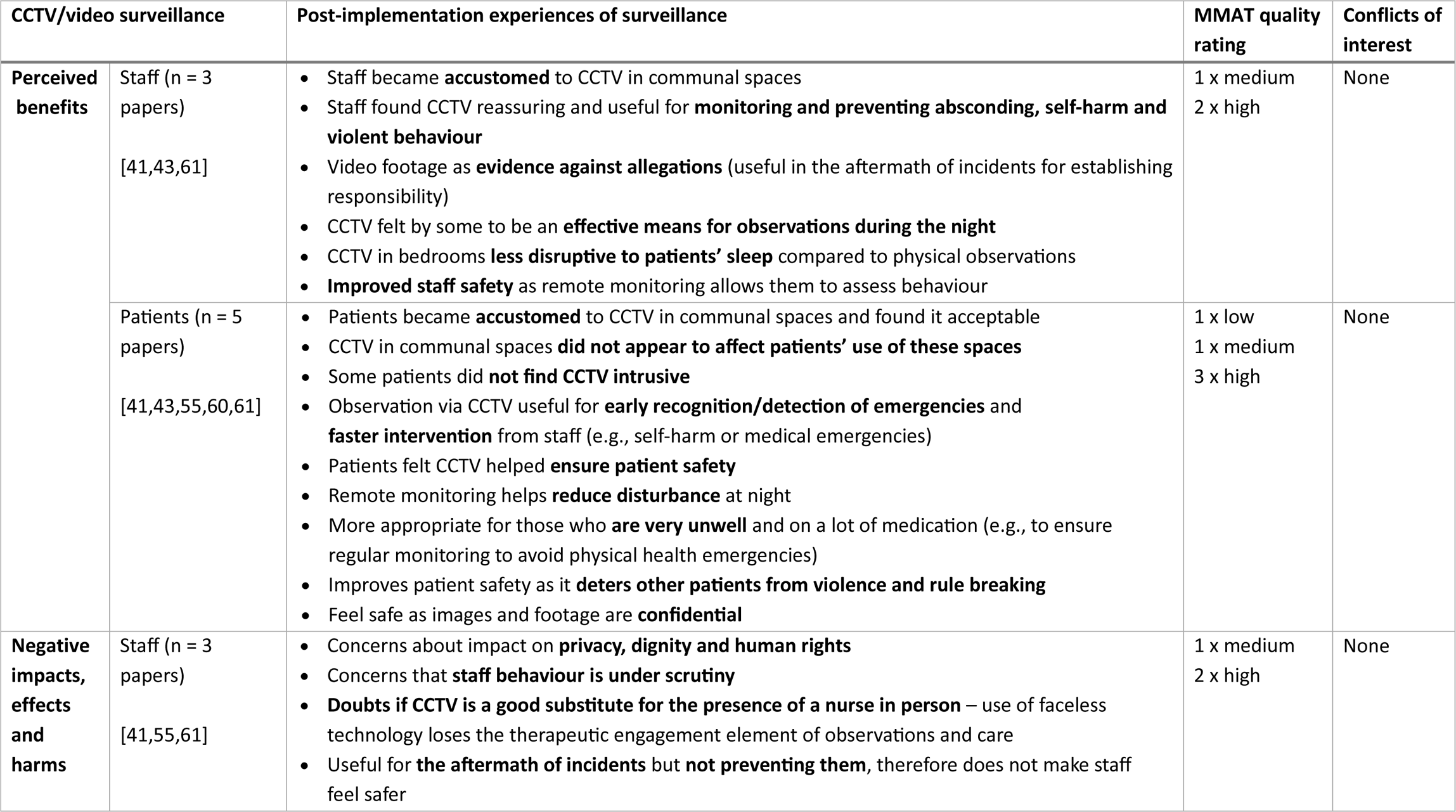
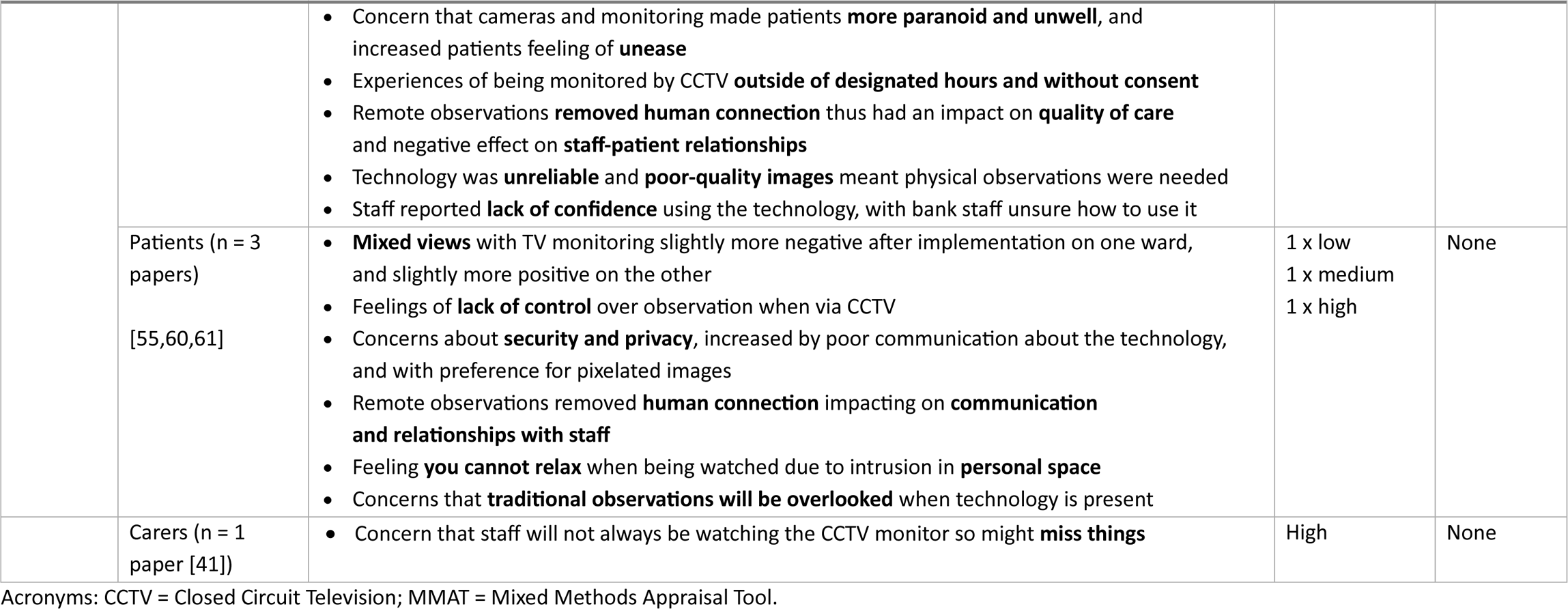
Staff, patient and carer post-implementation experiences of Closed Circuit Television (CCTV)/video surveillance.

##### Staff

Staff’s experiences of CCTV in communal spaces varied [41,43,60]. Some identified benefits including improved staff and patient safety, monitoring of self-harm, violence and absconding. However, others doubted its ability to control behaviour or prevent incidents. Some saw value in using CCTV to provide evidence to investigate incidents and allegations and felt it could be used to scrutinise staff behaviour. Ethical concerns were raised about its impact on patients’ privacy, dignity and human rights, and on therapeutic engagement. Some staff felt CCTV should not be used as a substitute for in-person care [41].

Staff’s views of CCTV use in patients’ bedrooms at night were also mixed [41,43,61]. Perceived benefits included improved monitoring of patients, enhanced staff safety, and reduced disruption of patients’ sleep compared to physical checks. Some staff felt they could rely on CCTV for patient observation, whereas others emphasised the importance of still conducing physical checks. Some staff raised concerns about negative impacts of CCTV in patients’ bedrooms on privacy, increased patient distress and paranoia, and reduced opportunities for therapeutic engagement. There were also reports of staff feeling uncertain about how to use the technology, using it incorrectly, finding it unreliable and it producing low quality images [61].

##### Patients

Patients had mixed views on CCTV monitoring in communal areas. Some felt it enhanced staff and patient safety, while others considered it an invasion of privacy. CCTV use in communal areas did not appear to affect patients’ use of these spaces [43]. In seclusion rooms, some patients believed CCTV could aid staff observations, prevent self-harm, help recognise emergencies and foster a sense of safety. However, concerns included a lack of control, privacy issues and security concerns, worsened by poor communication about the technology [55].

Regarding CCTV use in patients’ bedrooms at night, some patients found it enhanced their sense of safety, for example by deterring other patients from rule-breaking or stealing property. Some considered it less invasive and disruptive to sleep than physical checks since it reduced staff movement and the frequency of staff entering bedrooms for checks. However, others felt it was intrusive, impeded relaxation, negatively impacted therapeutic relationships with staff, and feared that it could result in traditional observations being neglected. Misunderstandings amongst patients about how and when CCTV was being used were reported, and there were also instances where patients were video monitored in their bedrooms outside of designated times or without consent [61].

##### Carers

One study reported that carers had concerns staff would not always be monitoring CCTV and so may miss things [41].

#### Body Worn Cameras (BWCs)

Two papers explored post-implementation experiences of BWCs [47,62] (see Table 9 for full results). Neither reported any conflicts of interest. One was rated high quality [62] and one low quality [47]. Staff and patient experiences were mixed, no carer experiences were reported.

**Table 9.** Staff, patient and carer post-implementation experiences of Body Worn Cameras (BWCs)

| Body Worn Cameras (BWCs) |  | Post-implementation experiences of surveillance | MMAT quality rating | Conflicts of interest |
| --- | --- | --- | --- | --- |
| Perceived benefits | Staff (n = 2 papers)<br>[47,62] | • **Staff wearing cameras were more positive** than other staff about them; **staff not wearing them had more mixed views**<br>• Belief or experience that it **prevents violence and aggression**<br>• Useful as **evidence for complaints/resolving incidents**<br>• Considered a useful tool for **prosecution** following incidents of violence<br>• Staff often felt BWC footage could be used **to protect them against false accusations of misconduct**<br>• **Reassured** in their techniques in restraint when cameras are on<br>• May reveal when **staff are not behaving professionally**<br>• Potential use in **training** with some staff expressing a desire to watch footage of incidents during a debrief with a manager to reflect on their own behaviour and consider what they might do differently in future.<br>• Some staff believed BWCs would be useful **for documenting physical restraint and planned interventions**, potentially **reducing restrictive practice** and **increasing physical safety for both staff and patients**.<br>• Staff tended to believe BWCs could make wards a safer place by **improving staff awareness and reflexive practice**, rather than changing patient behaviour. | 1 x low<br>1 x high | None |
|  | Patients (n = 1 paper [62]) | • Many patients expressed feeling unheard, ignored and not believed by staff – BWCs may make patients feel **safer by providing evidence to back up their claims**.<br>• Patients see the potential for BWCs to **protect them from staff misconduct**.<br>• Patients believed BWCs would be useful for **documenting physical restraint and planned interventions**, potentially **reducing restrictive practice** and **increasing physical safety** for both staff and patients.<br>• Patients tended to believe BWCs could make wards a safer place by **improving staff awareness and reflexive practice**, rather than changing patient behaviour. | High | None |
| **Negative impacts, effects and harms** | Staff (n = 2 papers)<br>[47,62] | Some staff found BWCs caused **discomfort** to wear<br>Concern footage **only captures from time of arrival, not the build up**<br>Staff **feel watched**<br>Patients appear to feel **intimidated** by the technology<br>Some staff feel BWCs **do not prevent violence and aggression** because they treat it as an individual issue **without addressing complex systemic causes**.<br>Some staff were concerned that BWCs would be treated as a **substitute for good care or safe staffing**.<br>Some staff raised **ethical and legal questions** about the role of **patient consent** in deciding when, or if, a BWC is turned on.<br>Some were concerned BWCs would feel like a **punitive measure** that singles out a patient, enhancing existing feelings of **criminalisation** and making the **ward feel less safe**. | 1 x low<br>1 x high | None |
|  | Patients (n = 2 papers)<br>[47,62] | **Negative impact on relationship between staff and patients** with patients expressing hesitation about speaking with staff members wearing a camera, regardless of whether it is on or off.<br>Some patients were concerned that BWCs would be treated as a **substitute for good care or safe staffing**.<br>Some patients raised **ethical and legal questions** about the role of **patient consent** in deciding when, or if, a BWC is turned on.<br>Some patients were concerned BWCs would feel like a **punitive measure** that singles out a patient, enhancing existing feelings of **criminalisation** and making the **ward feel less safe**.<br>Increased feeling of **staff having power and control over patients**.<br>Patients were concerned about being recorded in their **most vulnerable moments** and the impact BWCs might have on their **recovery, dignity and privacy**. | 1 x low<br>1 x high | None |

Acronyms: BWCs = Body Worn Cameras; MMAT = Mixed Methods Appraisal Tool.

##### Staff

Benefits of BWCs perceived by staff included reduced violence, aggression and restrictive practices. Some staff felt that they improved safety by improving staff awareness and reflexive practice, rather than changing patient behaviour. Staff identified various uses for BWC footage including: providing evidence to aid incident and complaint resolution (including ‘false allegations’ against staff) and prosecutions; documenting interventions (e.g., physical restraints); and facilitating debriefing and staff training. However, some staff raised concerns that BWCs only capture footage from the time of arrival, not the preceding events, and doubted their effectiveness in reducing violence and aggression as they do not address their underlying systemic causes. Some staff viewed BWCs as a punitive measure, contributing to patients’ feelings of criminalization and intimidation. They also raised ethical and legal concerns around patient consent and the potential for BWCs to be used as a substitute for good care and safe staffing [47,62].

##### Patients

Whilst some patients reported feeling safer with BWCs due to them providing evidence to support their claims and protect them against staff misconduct, others felt BWCs did not improve safety and negatively impacted their recovery, privacy and dignity. Like staff, some patients felt that BWCs fail to address the systemic causes of violence and aggression, and that any improvements in safety are due to increased staff awareness and reflexivity, rather than changes in patient behaviour. Similar to staff, some patients viewed BWCs as punitive, contributing to feelings of criminalization and exacerbating power imbalances between patients and staff [47,62].

#### GPS electronic monitoring and wearable sensors

None of the included studies explored staff, patient or family/carer post-implementation experiences of GPS electronic monitoring or wearable sensors.

### Research objective 2b: What is the effect, including unintended consequences, harms, and benefits, of surveillance-based technologies in inpatient mental health settings for outcomes such as patient and staff safety and patient clinical improvement?

Eleven studies reported outcomes on the effectiveness of surveillance strategies in inpatient mental health settings [37,40,44,47,49,50,51,56,59,60,61]. Overall, the findings were limited and mixed. The findings below are reported by type of surveillance and tabulated in Table 10.

**Table 10.**
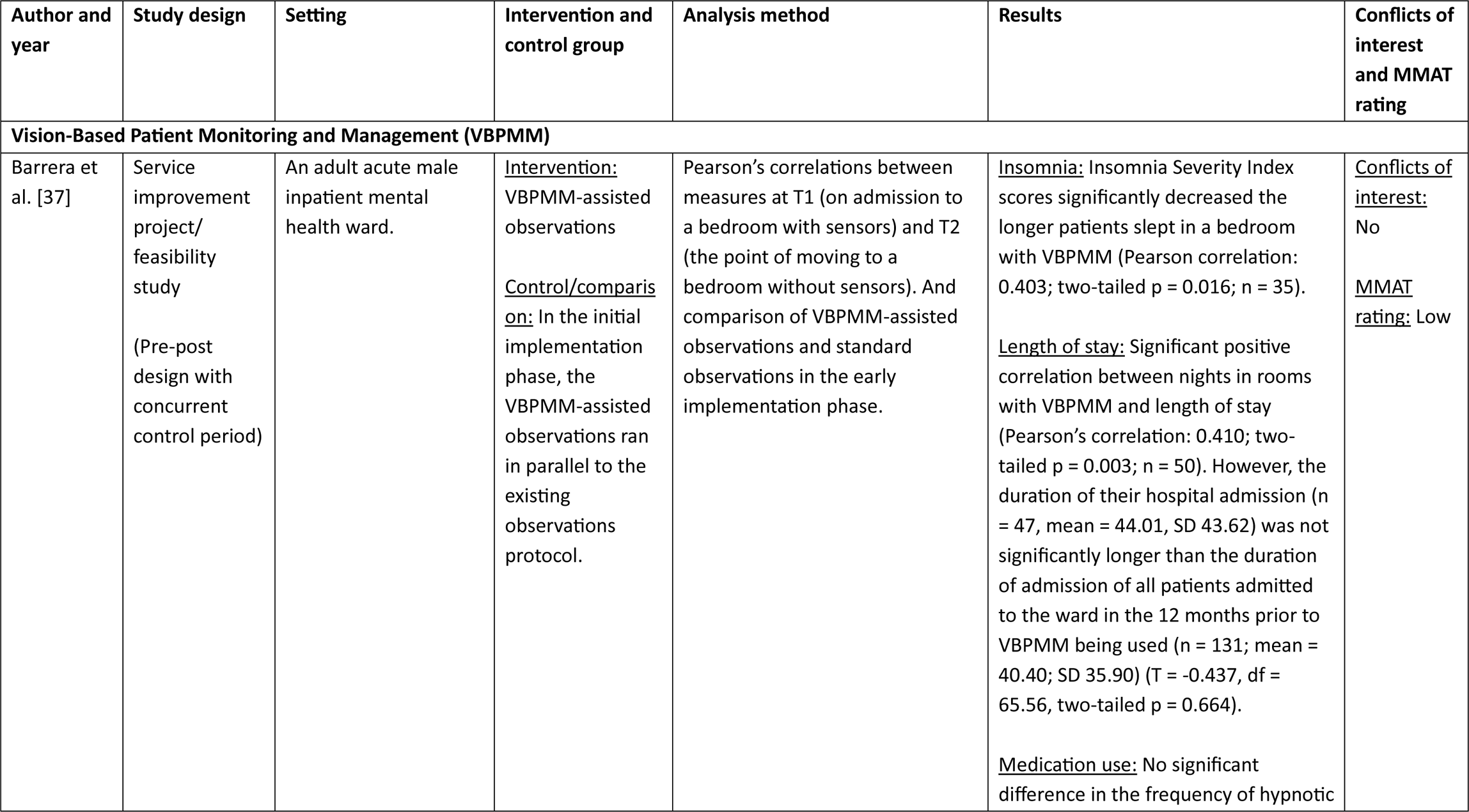
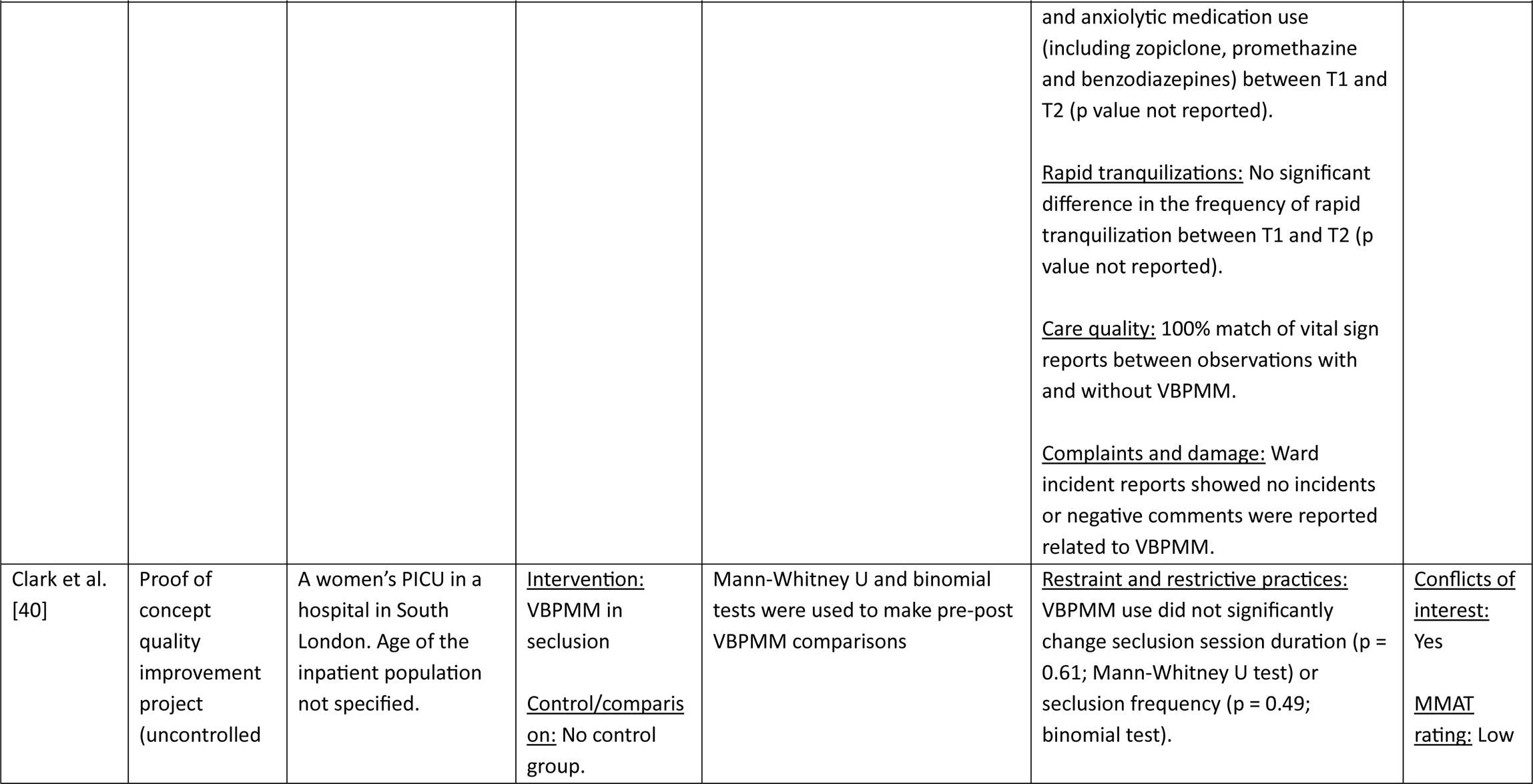
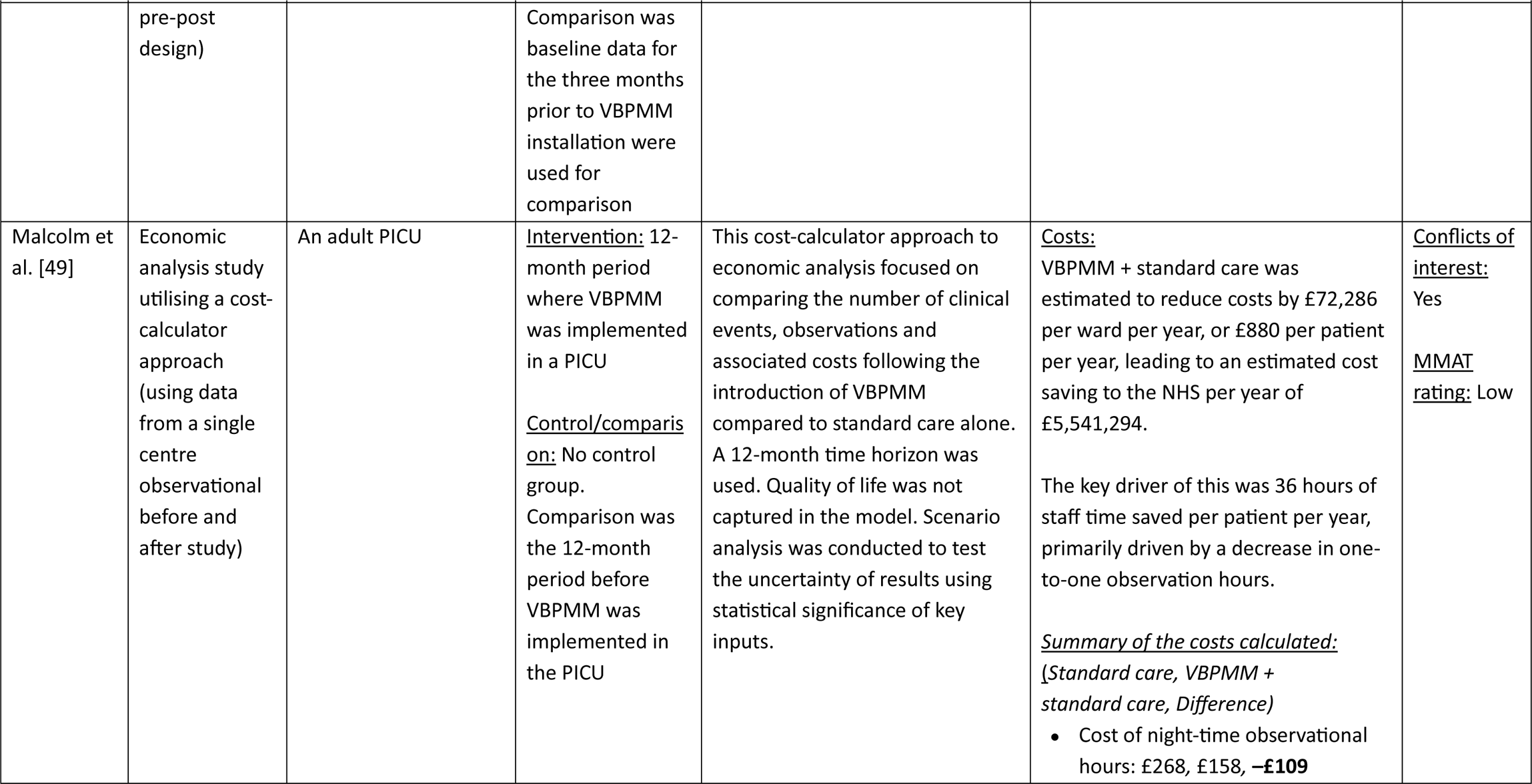
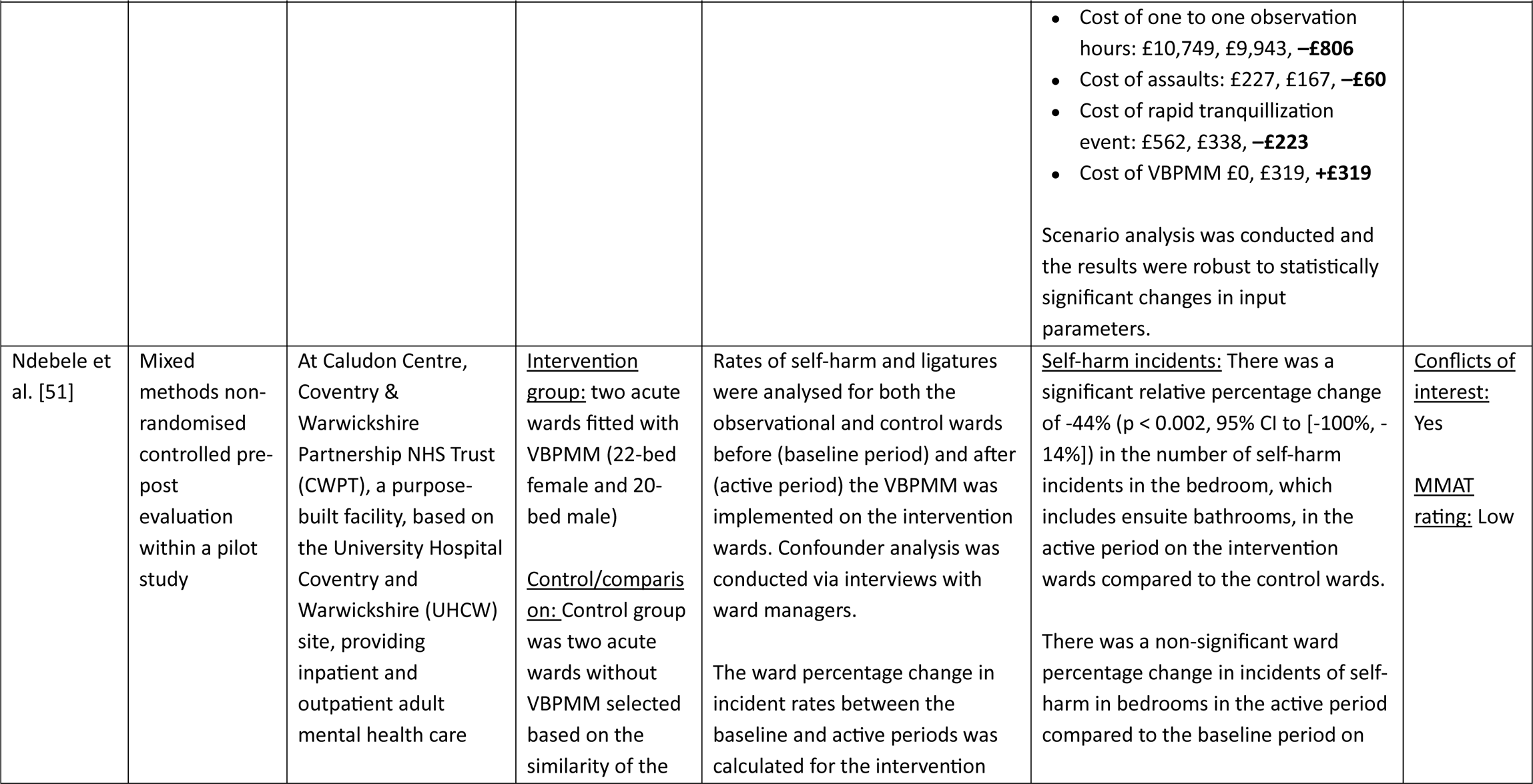
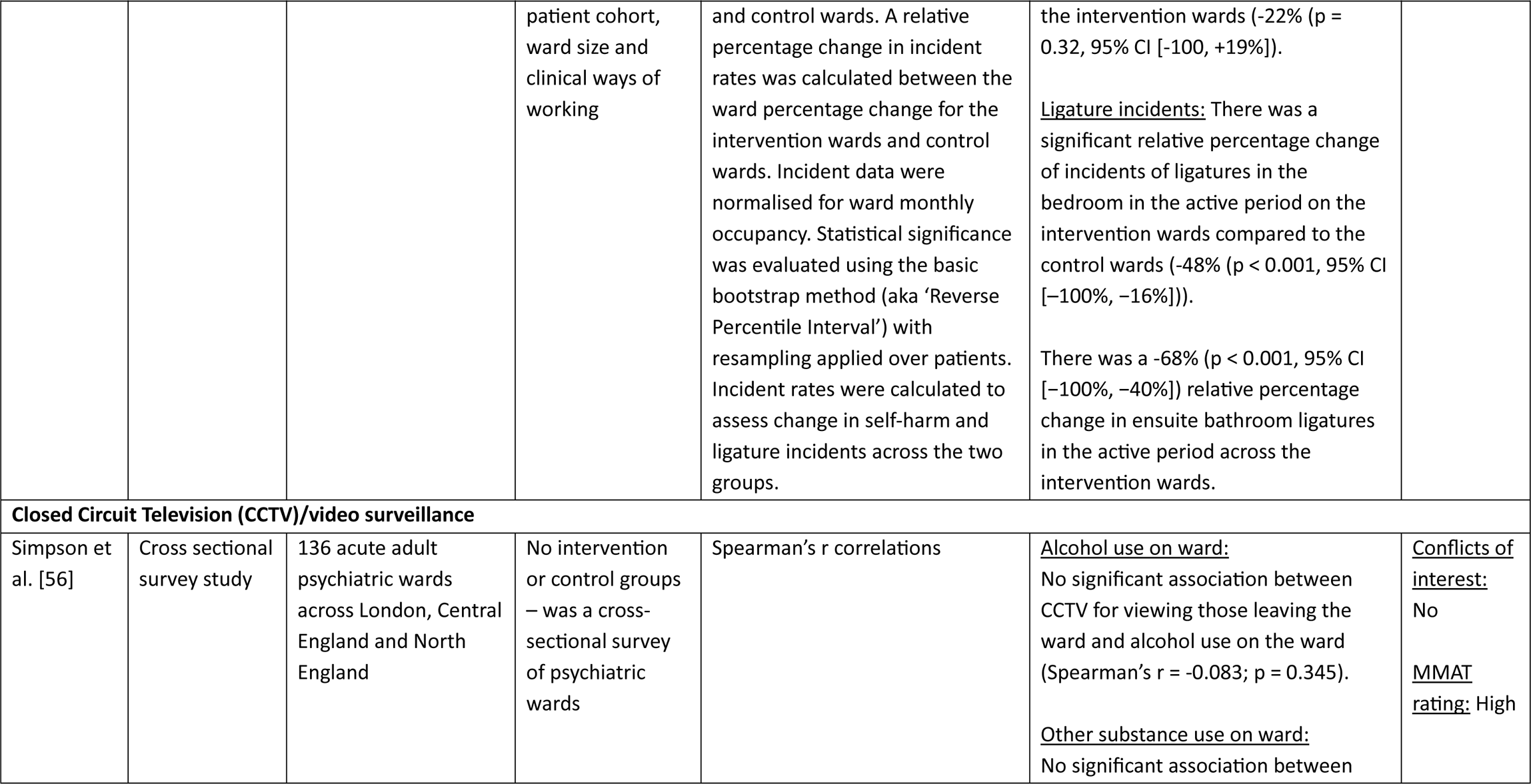
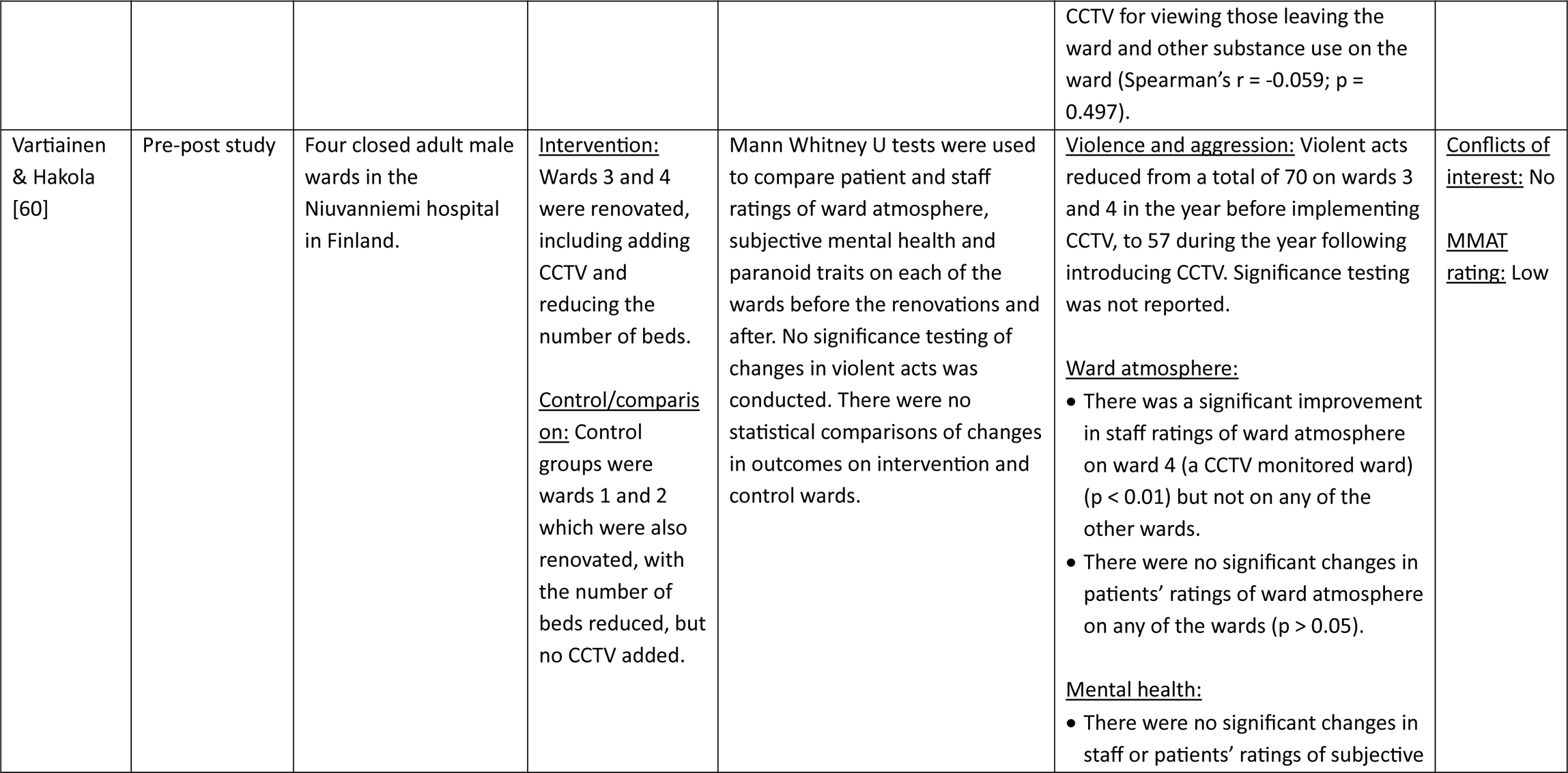
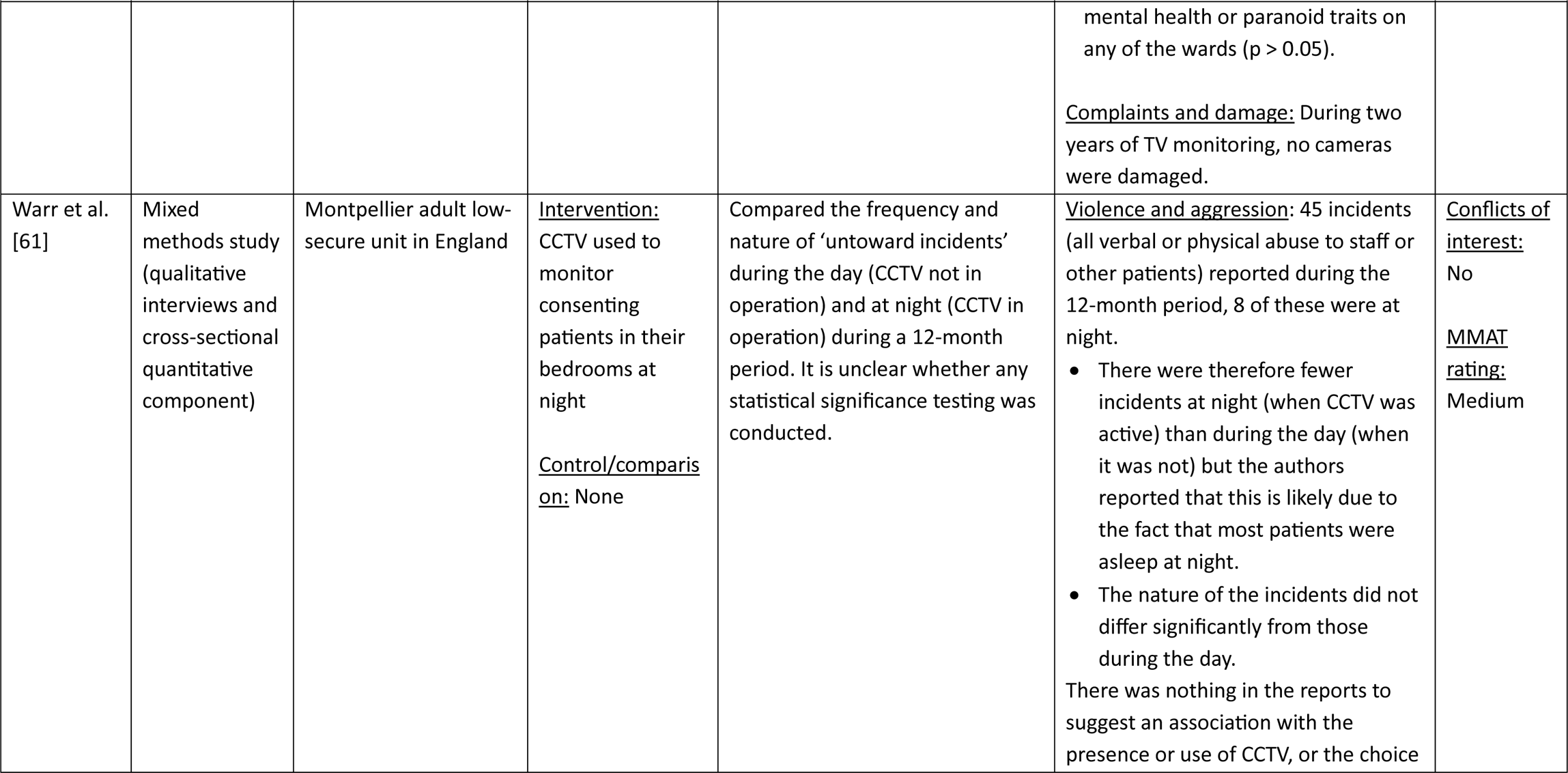
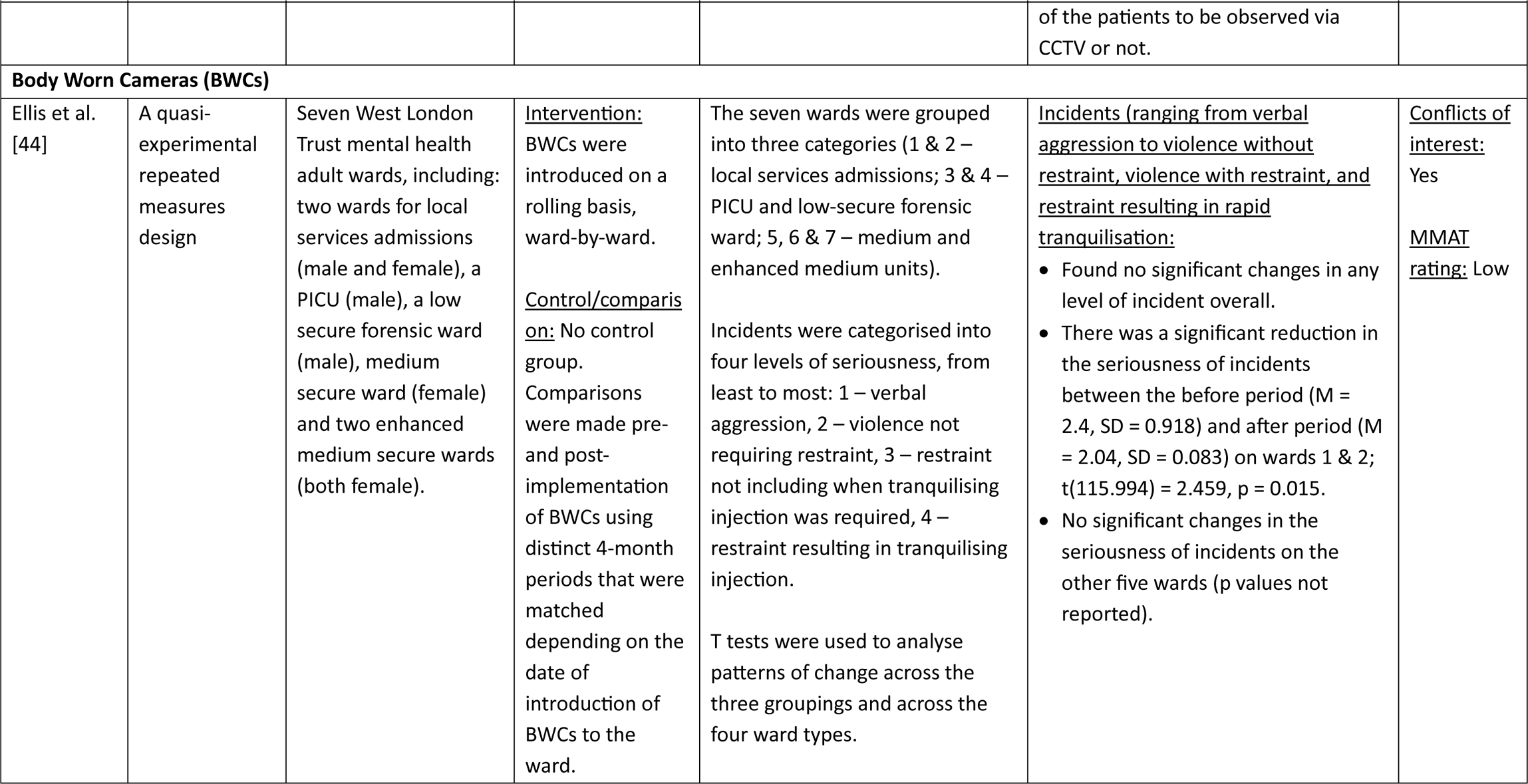
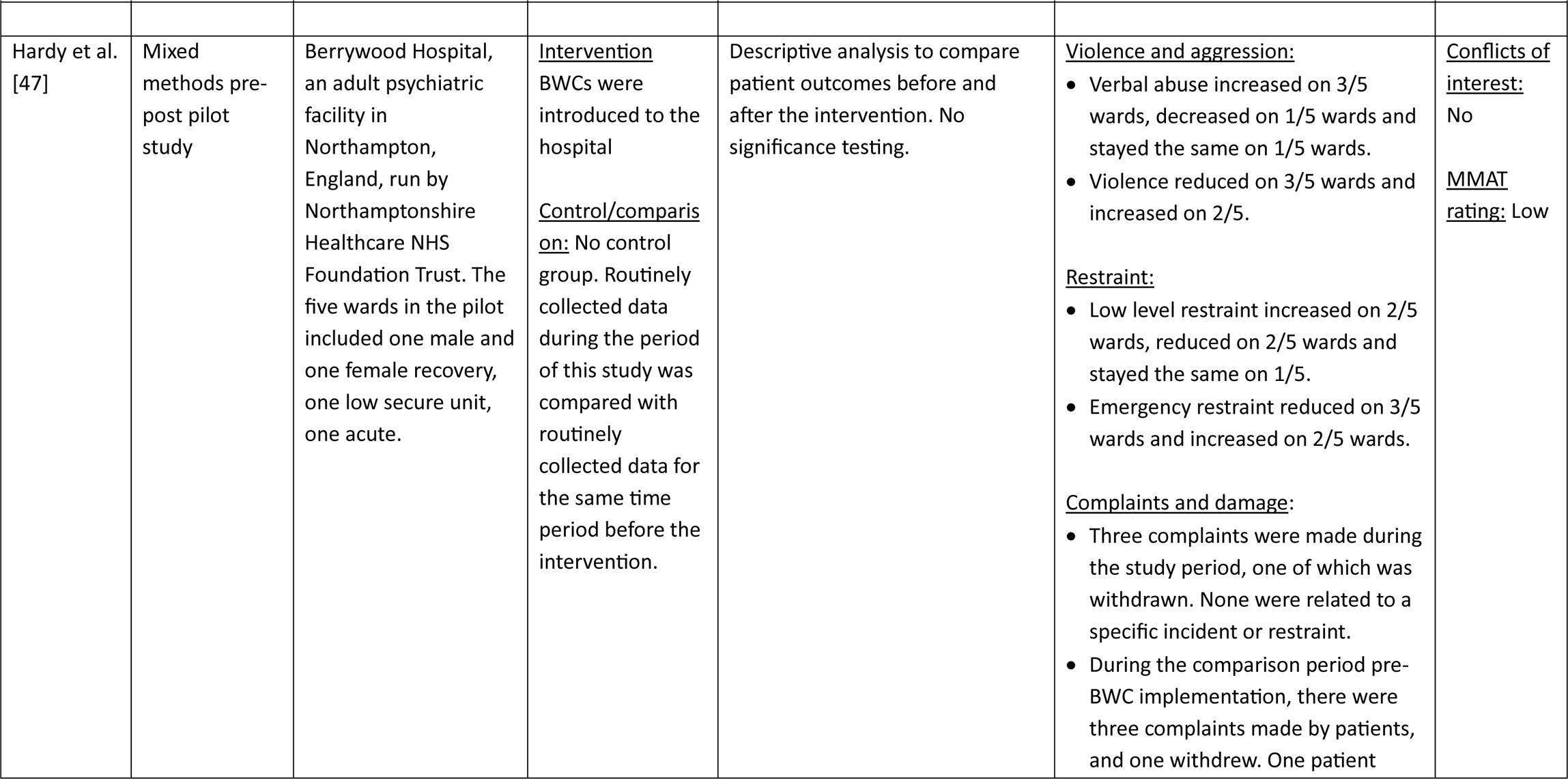
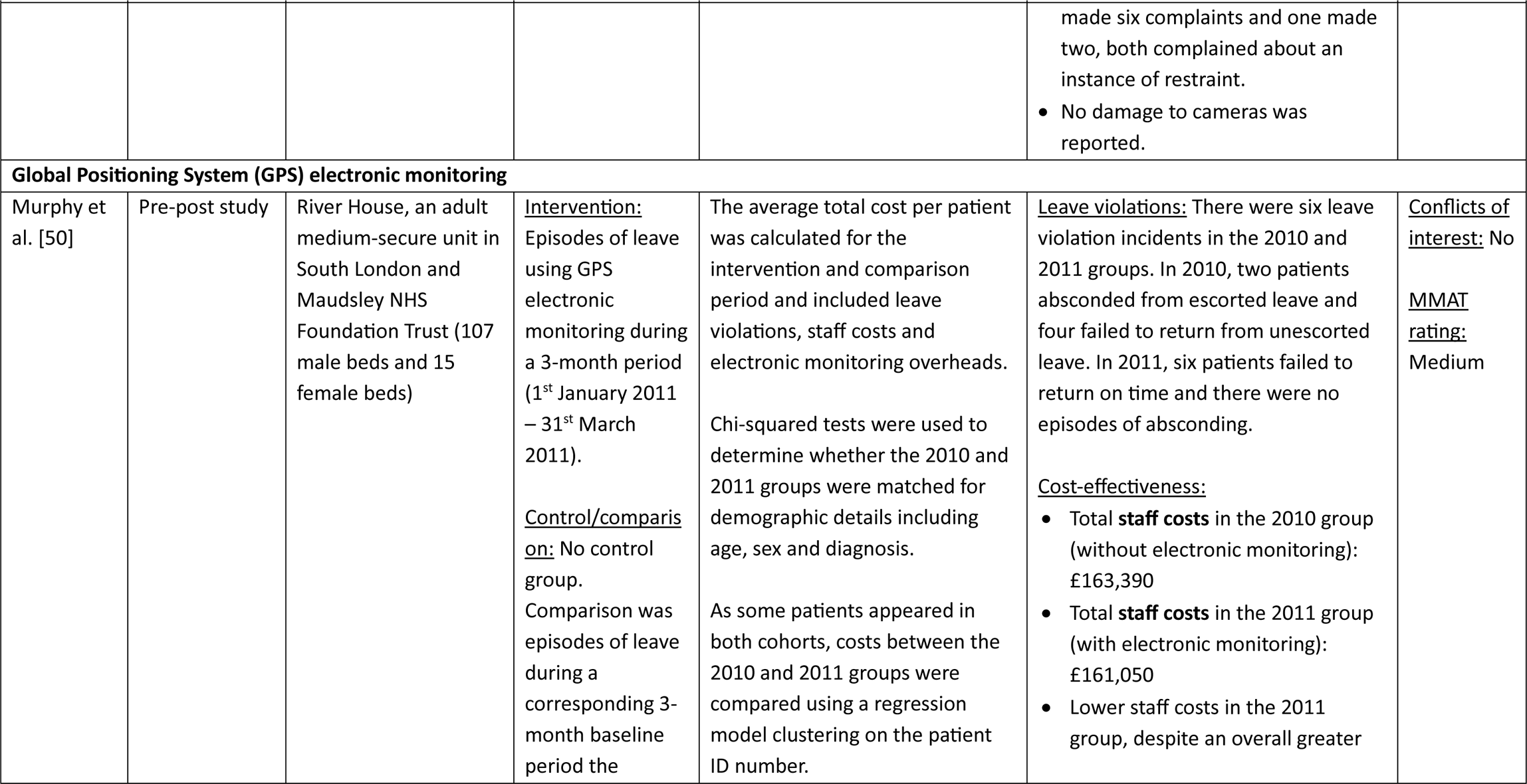
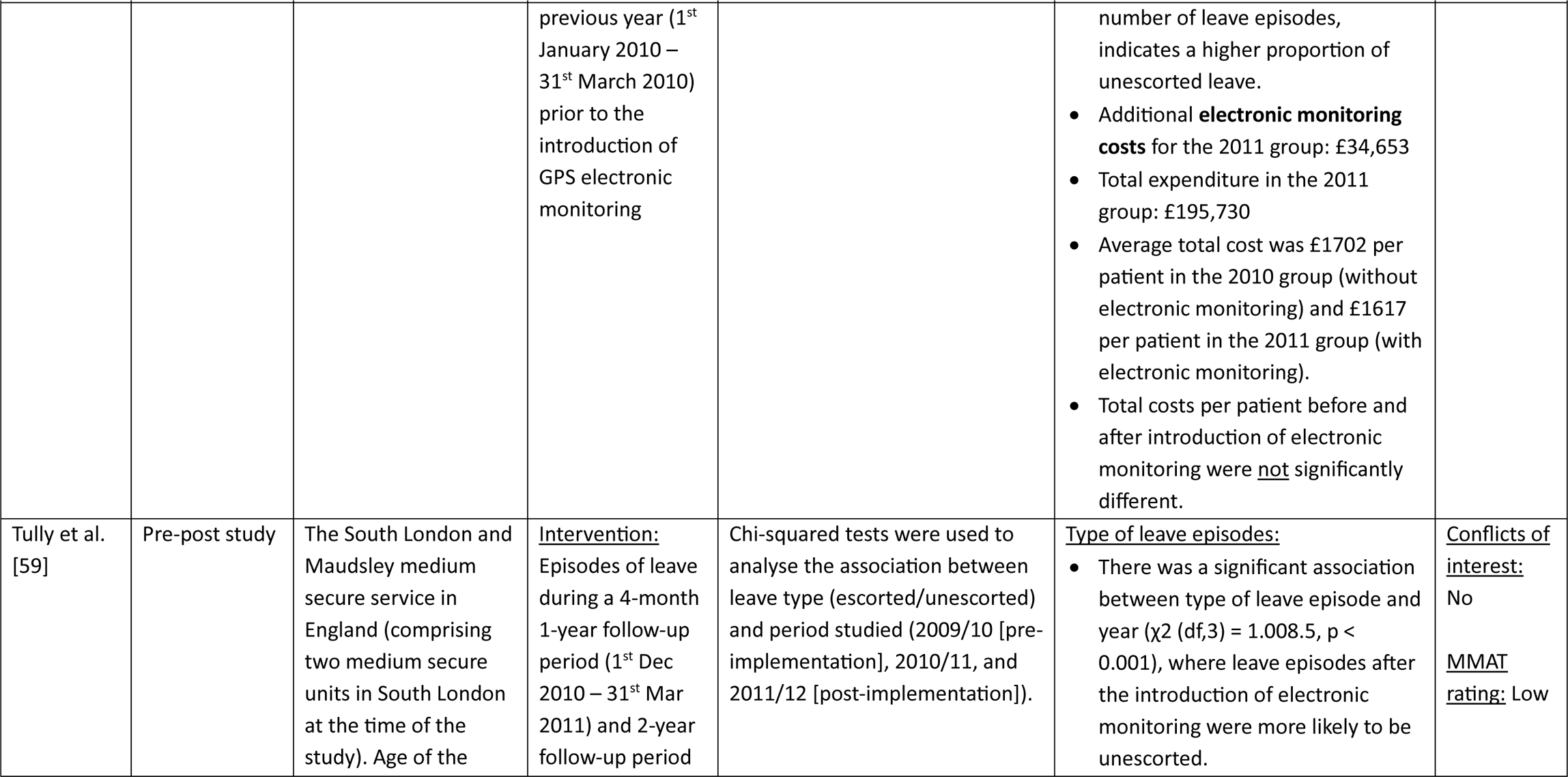
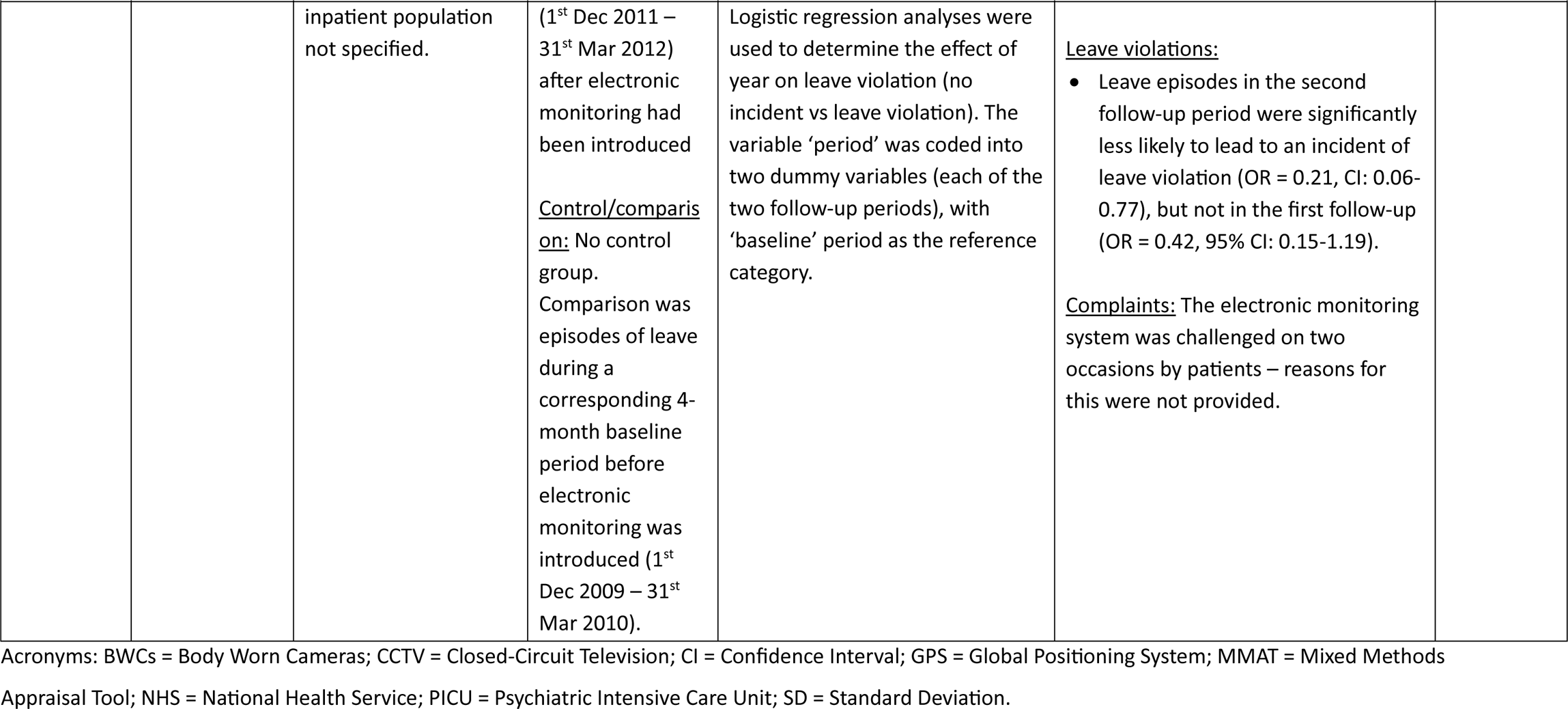
Quantitative evidence for the impact of surveillance technologies in inpatient mental health settings.

| Author and year | Study design | Setting | Intervention and control group | Analysis method | Results | Conflicts of interest and MMAT rating |
| --- | --- | --- | --- | --- | --- | --- |
| **Vision-Based Patient Monitoring and Management (VBPMM)** |  |  |  |  |  |  |
| Barrera et al. [37] | Service improvement project/feasibility study<br>(Pre-post design with concurrent control period) | An adult acute male inpatient mental health ward. | <u>Intervention:</u><br>VBPMM-assisted observations<br><u>Control/comparison:</u> In the initial implementation phase, the VBPMM-assisted observations ran in parallel to the existing observations protocol. | Pearson's correlations between measures at T1 (on admission to a bedroom with sensors) and T2 (the point of moving to a bedroom without sensors). And comparison of VBPMM-assisted observations and standard observations in the early implementation phase. | <u>Insomnia:</u> Insomnia Severity Index scores significantly decreased the longer patients slept in a bedroom with VBPMM (Pearson correlation: 0.403; two-tailed p = 0.016; n = 35).<br><u>Length of stay:</u> Significant positive correlation between nights in rooms with VBPMM and length of stay (Pearson's correlation: 0.410; two-tailed p = 0.003; n = 50). However, the duration of their hospital admission (n = 47, mean = 44.01, SD 43.62) was not significantly longer than the duration of admission of all patients admitted to the ward in the 12 months prior to VBPMM being used (n = 131; mean = 40.40; SD 35.90) (T = -0.437, df = 65.56, two-tailed p = 0.664).<br><u>Medication use:</u> No significant difference in the frequency of hypnotic<br>and anxiolytic medication use (including zopiclone, promethazine and benzodiazepines) between T1 and T2 (p value not reported).<br><u>Rapid tranquilizations:</u> No significant difference in the frequency of rapid tranquilization between T1 and T2 (p value not reported).<br><u>Care quality:</u> 100% match of vital sign reports between observations with and without VBPM.<br><u>Complaints and damage:</u> Ward incident reports showed no incidents or negative comments were reported related to VBPM. | <u>Conflicts of interest:</u><br>No<br><u>MMAT rating:</u> Low |
| Clark et al. [40] | Proof of concept quality improvement project (uncontrolled | A women's PICU in a hospital in South London. Age of the inpatient population not specified. | <u>Intervention:</u> VBPM in seclusion<br><u>Control/comparison:</u> No control group. | Mann-Whitney U and binomial tests were used to make pre-post VBPM comparisons | <u>Restraint and restrictive practices:</u> VBPM use did not significantly change seclusion session duration (p = 0.61; Mann-Whitney U test) or seclusion frequency (p = 0.49; binomial test). | <u>Conflicts of interest:</u> Yes<br><u>MMAT rating:</u> Low |
|  | pre-post design) |  | Comparison was baseline data for the three months prior to VBPM installation were used for comparison |  |  |  |
| Malcolm et al. [49] | Economic analysis study utilising a cost-calculator approach (using data from a single centre observational before and after study) | An adult PICU | <u>Intervention:</u> 12-month period where VBPM was implemented in a PICU<br><u>Control/comparison:</u> No control group. Comparison was the 12-month period before VBPM was implemented in the PICU | This cost-calculator approach to economic analysis focused on comparing the number of clinical events, observations and associated costs following the introduction of VBPM compared to standard care alone. A 12-month time horizon was used. Quality of life was not captured in the model. Scenario analysis was conducted to test the uncertainty of results using statistical significance of key inputs. | <u>Costs:</u><br>VBPM + standard care was estimated to reduce costs by £72,286 per ward per year, or £880 per patient per year, leading to an estimated cost saving to the NHS per year of £5,541,294.<br>The key driver of this was 36 hours of staff time saved per patient per year, primarily driven by a decrease in one-to-one observation hours.<br><u>Summary of the costs calculated:</u><br>(Standard care, VBPM + standard care, Difference)<br>Cost of night-time observational hours: £268, £158, **–£109** | <u>Conflicts of interest:</u><br>Yes<br><u>MMAT rating:</u> Low |
|  |  |  |  |  | • Cost of one to one observation hours: £10,749, £9,943, **-£806**<br>• Cost of assaults: £227, £167, **-£60**<br>• Cost of rapid tranquillization event: £562, £338, **-£223**<br>• Cost of VBPM £0, £319, **+£319**<br>Scenario analysis was conducted and the results were robust to statistically significant changes in input parameters. |  |
| Ndebele et al. [51] | Mixed methods non-randomised controlled pre-post evaluation within a pilot study | At Caludon Centre, Coventry & Warwickshire Partnership NHS Trust (CWPT), a purpose-built facility, based on the University Hospital Coventry and Warwickshire (UHCW) site, providing inpatient and outpatient adult mental health care | <u>Intervention group</u>: two acute wards fitted with VBPM (22-bed female and 20-bed male)<br><u>Control/comparison</u>: Control group was two acute wards without VBPM selected based on the similarity of the | Rates of self-harm and ligatures were analysed for both the observational and control wards before (baseline period) and after (active period) the VBPM was implemented on the intervention wards. Confounder analysis was conducted via interviews with ward managers.<br>The ward percentage change in incident rates between the baseline and active periods was calculated for the intervention | <u>Self-harm incidents</u>: There was a significant relative percentage change of -44% (p < 0.002, 95% CI to [-100%, -14%]) in the number of self-harm incidents in the bedroom, which includes ensuite bathrooms, in the active period on the intervention wards compared to the control wards.<br>There was a non-significant ward percentage change in incidents of self-harm in bedrooms in the active period compared to the baseline period on | <u>Conflicts of interest</u>: Yes<br><u>MMAT rating</u>: Low |
|  |  |  | patient cohort, ward size and clinical ways of working | and control wards. A relative percentage change in incident rates was calculated between the ward percentage change for the intervention wards and control wards. Incident data were normalised for ward monthly occupancy. Statistical significance was evaluated using the basic bootstrap method (aka 'Reverse Percentile Interval') with resampling applied over patients. Incident rates were calculated to assess change in self-harm and ligature incidents across the two groups. | the intervention wards (-22% (p = 0.32, 95% CI [-100, +19%])).<br><u>Ligature incidents:</u> There was a significant relative percentage change of incidents of ligatures in the bedroom in the active period on the intervention wards compared to the control wards (-48% (p < 0.001, 95% CI [-100%, -16%])).<br>There was a -68% (p < 0.001, 95% CI [-100%, -40%]) relative percentage change in ensuite bathroom ligatures in the active period across the intervention wards. |  |
| **Closed Circuit Television (CCTV)/video surveillance** |  |  |  |  |  |  |
| Simpson et al. [56] | Cross sectional survey study | 136 acute adult psychiatric wards across London, Central England and North England | No intervention or control groups – was a cross-sectional survey of psychiatric wards | Spearman's r correlations | <u>Alcohol use on ward:</u><br>No significant association between CCTV for viewing those leaving the ward and alcohol use on the ward (Spearman's r = -0.083; p = 0.345).<br><u>Other substance use on ward:</u><br>No significant association between | <u>Conflicts of interest:</u><br>No<br><u>MMAT rating:</u> High |
| | | | | | CCTV for viewing those leaving the ward and other substance use on the ward (Spearman's $r = -0.059$; $p = 0.497$). | |
| Vartiainen & Hakola [60] | Pre-post study | Four closed adult male wards in the Niuvanniemi hospital in Finland. | <u>Intervention:</u> Wards 3 and 4 were renovated, including adding CCTV and reducing the number of beds.<br><u>Control/comparison:</u> Control groups were wards 1 and 2 which were also renovated, with the number of beds reduced, but no CCTV added. | Mann Whitney U tests were used to compare patient and staff ratings of ward atmosphere, subjective mental health and paranoid traits on each of the wards before the renovations and after. No significance testing of changes in violent acts was conducted. There were no statistical comparisons of changes in outcomes on intervention and control wards. | <u>Violence and aggression:</u> Violent acts reduced from a total of 70 on wards 3 and 4 in the year before implementing CCTV, to 57 during the year following introducing CCTV. Significance testing was not reported.<br><u>Ward atmosphere:</u><br>• There was a significant improvement in staff ratings of ward atmosphere on ward 4 (a CCTV monitored ward) ($p < 0.01$) but not on any of the other wards.<br>• There were no significant changes in patients' ratings of ward atmosphere on any of the wards ($p > 0.05$).<br><u>Mental health:</u><br>• There were no significant changes in staff or patients' ratings of subjective<br>mental health or paranoid traits on any of the wards ($p > 0.05$).<br><u>Complaints and damage:</u> During two years of TV monitoring, no cameras were damaged. | <u>Conflicts of interest:</u> No<br><u>MMAT rating:</u> Low |
| Warr et al. [61] | Mixed methods study (qualitative interviews and cross-sectional quantitative component) | Montpellier adult low-secure unit in England | <u>Intervention:</u> CCTV used to monitor consenting patients in their bedrooms at night<br><u>Control/comparison:</u> None | Compared the frequency and nature of 'untoward incidents' during the day (CCTV not in operation) and at night (CCTV in operation) during a 12-month period. It is unclear whether any statistical significance testing was conducted. | <u>Violence and aggression:</u> 45 incidents (all verbal or physical abuse to staff or other patients) reported during the 12-month period, 8 of these were at night.<br>There were therefore fewer incidents at night (when CCTV was active) than during the day (when it was not) but the authors reported that this is likely due to the fact that most patients were asleep at night.<br>The nature of the incidents did not differ significantly from those during the day.<br>There was nothing in the reports to suggest an association with the presence or use of CCTV, or the choice<br>of the patients to be observed via CCTV or not. | <u>Conflicts of interest:</u> No<br><u>MMAT rating:</u> Medium |
| **Body Worn Cameras (BWCs)** |  |  |  |  |  |  |
| Ellis et al. [44] | A quasi-experimental repeated measures design | Seven West London Trust mental health adult wards, including: two wards for local services admissions (male and female), a PICU (male), a low secure forensic ward (male), medium secure ward (female) and two enhanced medium secure wards (both female). | <u>Intervention:</u> BWCs were introduced on a rolling basis, ward-by-ward.<br><u>Control/comparison:</u> No control group. Comparisons were made pre- and post-implementation of BWCs using distinct 4-month periods that were matched depending on the date of introduction of BWCs to the ward. | The seven wards were grouped into three categories (1 & 2 – local services admissions; 3 & 4 – PICU and low-secure forensic ward; 5, 6 & 7 – medium and enhanced medium units).<br>Incidents were categorised into four levels of seriousness, from least to most: 1 – verbal aggression, 2 – violence not requiring restraint, 3 – restraint not including when tranquilising injection was required, 4 – restraint resulting in tranquilising injection.<br>T tests were used to analyse patterns of change across the three groupings and across the four ward types. | <u>Incidents (ranging from verbal aggression to violence without restraint, violence with restraint, and restraint resulting in rapid tranquilisation):</u><br>• Found no significant changes in any level of incident overall.<br>• There was a significant reduction in the seriousness of incidents between the before period ($M = 2.4$, $SD = 0.918$) and after period ($M = 2.04$, $SD = 0.083$) on wards 1 & 2; $t(115.994) = 2.459$, $p = 0.015$.<br>• No significant changes in the seriousness of incidents on the other five wards ($p$ values not reported). | <u>Conflicts of interest:</u> Yes<br><u>MMAT rating:</u> Low |
| Hardy et al. [47] | Mixed methods pre-post pilot study | Berrywood Hospital, an adult psychiatric facility in Northampton, England, run by Northamptonshire Healthcare NHS Foundation Trust. The five wards in the pilot included one male and one female recovery, one low secure unit, one acute. | <u>Intervention</u><br>BWCs were introduced to the hospital<br><u>Control/comparison:</u> No control group. Routinely collected data during the period of this study was compared with routinely collected data for the same time period before the intervention. | Descriptive analysis to compare patient outcomes before and after the intervention. No significance testing. | <u>Violence and aggression:</u><br>• Verbal abuse increased on 3/5 wards, decreased on 1/5 wards and stayed the same on 1/5 wards.<br>• Violence reduced on 3/5 wards and increased on 2/5.<br><u>Restraint:</u><br>• Low level restraint increased on 2/5 wards, reduced on 2/5 wards and stayed the same on 1/5.<br>• Emergency restraint reduced on 3/5 wards and increased on 2/5 wards.<br><u>Complaints and damage:</u><br>• Three complaints were made during the study period, one of which was withdrawn. None were related to a specific incident or restraint.<br>• During the comparison period pre-BWC implementation, there were three complaints made by patients, and one withdrew. One patient<br>made six complaints and one made two, both complained about an instance of restraint.<br>No damage to cameras was reported. | <u>Conflicts of interest:</u><br>No<br><u>MMAT rating:</u> Low |
| **Global Positioning System (GPS) electronic monitoring** |  |  |  |  |  |  |
| Murphy et al. [50] | Pre-post study | River House, an adult medium-secure unit in South London and Maudsley NHS Foundation Trust (107 male beds and 15 female beds) | <u>Intervention:</u> Episodes of leave using GPS electronic monitoring during a 3-month period (1<sup>st</sup> January 2011 – 31<sup>st</sup> March 2011).<br><u>Control/comparison:</u> No control group. Comparison was episodes of leave during a corresponding 3-month baseline period the | The average total cost per patient was calculated for the intervention and comparison period and included leave violations, staff costs and electronic monitoring overheads.<br>Chi-squared tests were used to determine whether the 2010 and 2011 groups were matched for demographic details including age, sex and diagnosis.<br>As some patients appeared in both cohorts, costs between the 2010 and 2011 groups were compared using a regression model clustering on the patient ID number. | <u>Leave violations:</u> There were six leave violation incidents in the 2010 and 2011 groups. In 2010, two patients absconded from escorted leave and four failed to return from unescorted leave. In 2011, six patients failed to return on time and there were no episodes of absconding.<br><u>Cost-effectiveness:</u><br>Total **staff costs** in the 2010 group (without electronic monitoring): £163,390<br>Total **staff costs** in the 2011 group (with electronic monitoring): £161,050<br>Lower staff costs in the 2011 group, despite an overall greater | <u>Conflicts of interest:</u> No<br><u>MMAT rating:</u> Medium |
|  |  |  | previous year (1<sup>st</sup> January 2010 – 31<sup>st</sup> March 2010) prior to the introduction of GPS electronic monitoring |  | number of leave episodes, indicates a higher proportion of unescorted leave.<br>• Additional **electronic monitoring costs** for the 2011 group: £34,653<br>• Total expenditure in the 2011 group: £195,730<br>• Average total cost was £1702 per patient in the 2010 group (without electronic monitoring) and £1617 per patient in the 2011 group (with electronic monitoring).<br>• Total costs per patient before and after introduction of electronic monitoring were <u>not</u> significantly different. |  |
| Tully et al. [59] | Pre-post study | The South London and Maudsley medium secure service in England (comprising two medium secure units in South London at the time of the study). Age of the | <u>Intervention:</u><br>Episodes of leave during a 4-month 1-year follow-up period (1<sup>st</sup> Dec 2010 – 31<sup>st</sup> Mar 2011) and 2-year follow-up period | Chi-squared tests were used to analyse the association between leave type (escorted/unescorted) and period studied (2009/10 [pre-implementation], 2010/11, and 2011/12 [post-implementation]). | <u>Type of leave episodes:</u><br>• There was a significant association between type of leave episode and year ($\chi^2$ (df,3) = 1.008.5, $p < 0.001$), where leave episodes after the introduction of electronic monitoring were more likely to be unescorted. | <u>Conflicts of interest:</u><br>No<br><u>MMAT rating:</u> Low |
|  |  | inpatient population not specified. | (1<sup>st</sup> Dec 2011 – 31<sup>st</sup> Mar 2012) after electronic monitoring had been introduced<br><u>Control/comparison:</u> No control group. Comparison was episodes of leave during a corresponding 4-month baseline period before electronic monitoring was introduced (1<sup>st</sup> Dec 2009 – 31<sup>st</sup> Mar 2010). | Logistic regression analyses were used to determine the effect of year on leave violation (no incident vs leave violation). The variable 'period' was coded into two dummy variables (each of the two follow-up periods), with 'baseline' period as the reference category. | <u>Leave violations:</u><br>• Leave episodes in the second follow-up period were significantly less likely to lead to an incident of leave violation (OR = 0.21, CI: 0.06-0.77), but not in the first follow-up (OR = 0.42, 95% CI: 0.15-1.19).<br><u>Complaints:</u> The electronic monitoring system was challenged on two occasions by patients – reasons for this were not provided. |  |

Acronyms: BWCs = Body Worn Cameras; CCTV = Closed-Circuit Television; CI = Confidence Interval; GPS = Global Positioning System; MMAT = Mixed Methods
Appraisal Tool; NHS = National Health Service; PICU = Psychiatric Intensive Care Unit; SD = Standard Deviation.

#### Vision-Based Patient Monitoring and Management (VBPMM)

Four studies reported on the effect of VBPMM [37,40,49,51]. All studies reported on Oxevision by Oxehealth. All studies were rated low quality, and three declared conflicts of interest [40,49,51]. Study designs included a mixed methods non-randomised controlled pre-post evaluation within a pilot study, which compared two intervention wards with VBPMM to two control wards without VBPMM [51], an economic analysis study utilising a cost-calculator approach [49], an uncontrolled pre-post study [40] and a pre-post study with a concurrent control period [37].

##### Self-harm and ligature incidents

One study investigated VBPMM’s effect on self-harm and ligature incidents; it reported a significant relative reduction in self-harm and ligature incidents in bedrooms on the VBPMM wards *compared* to the control wards. However, when considering the VBPMM wards *alone*, there was a significant decrease in ligature incident rates, but not in self-harm rates, after introducing VBPMM [51].

##### Restrictive practices

Two studies reported on VBPMM’s effect on restrictive practices [37,40]. Barerra et al. [37] reported no significant effect on rapid tranquilization frequency, and Clark et al. [40] reported no significant impact upon seclusion session frequency or duration.

##### Clinical outcomes

One study investigated VBPMM’s effect on clinical outcomes [37]. It reported that insomnia severity significantly decreased the longer patients slept in a bedroom with VBPMM. There was a significant positive correlation between nights in rooms with VBPMM and hospital length of stay, although there was no significant difference in patients’ average hospital admission duration post-VBPMM and the average admission duration for all patients admitted to the ward in the 12 months before VBPMM introduction. There was also no significant difference in the use of hypnotic and anxiolytic medication [37].

##### Care quality

One study reported VBPMM’s effect on care quality related outcomes [37]. It reported a 100% match of vital sign reports between observations with and without sensors.

##### Cost effectiveness

One study investigated the cost-effectiveness of VBPMM [49]. It estimated that VBPMM in addition to standard care, compared to standard care alone, reduced costs by £72,286 per ward per year, or £880 per patient per year. It estimated that if rolled-out to all adult PICUs in England, VBPMM would lead to an estimated cost saving to the NHS per year of £5,541,294. The key driver of these savings was 36 hours of staff time saved per patient per year, primarily driven by a decrease in one-to-one observation hours. Scenario analyses showed that these results were robust to statistically significant changes in input parameters.

##### Complaints and damage

One study reported on VBPMM’s effect on complaints [37]; it reported that during the study period, no incidents related to VBPMM were recorded on the Trust’s online incident reporting system. During the first four nights of the new observation protocol (where VBPMM was used to conduct most observations of patients at night, instead of physical checks), eleven patients who completed questionnaires each night expressed no negative comments about the system. Details were not provided about how these patients were selected, or the format or content of the questionnaire.

#### Closed Circuit Television (CCTV)/video monitoring

Three studies [56,60,61] reported the effect of CCTV. One was rated high quality [56], one medium quality [61] and one low quality [60]. All three reported no conflicts of interest. One study had a cross-sectional design [56], one was mixed methods with a cross-sectional quantitative component [61] and one was a pre-post evaluation [60].

##### Violence and aggression

Two studies reported on CCTV’s effect on violence and aggression [60,61]. It is unclear whether Warr et al. [61], who investigated the impact of CCTV use in patients’ bedrooms at night on the frequency and nature of incidents, conducted any statistical significance testing. However, they reported that there were fewer incidents at night compared to during the day, but that there was no difference in the nature of the incidents. They also stated that there was no evidence of any association between the nature of incidents and the presence or use of CCTV, or the choice of the patient to be observed using CCTV or not. Vartiainen & Hakola [60] did not conduct any statistical significance testing but reported that violent acts reduced on the CCTV-monitored wards.

##### Clinical outcomes

Two studies reported on CCTV’s effect on clinical outcomes [56,60]. Simpson et al. [56] reported that CCTV (at exit) had no significant impact on substance or alcohol use on the ward. Vartiainen & Hakola [60] reported no significant changes in subjective mental health or paranoid traits on any of the wards (with or without CCTV).

##### Complaints and damage

One study reported on the impact of CCTV on damages [60]; it reported that no damage had occurred to cameras in two years of TV monitoring.

#### Body Worn Cameras (BWCs)

Two studies reported the effect of BWCs [44,47]. Both were rated as low quality and one declared a conflict of interest [44]. One had a quasi-experimental repeated-measures pilot study design [44], the other had a mixed methods uncontrolled pre-post pilot study design [47].

##### Violence and aggression

Both studies reported mixed results [44,47]. Ellis et al. [44] reported no significant changes in the overall numbers of violent and aggressive incidents. They reported a significant reduction in incident seriousness on two of the wards (‘local services admissions’ wards) but no significant changes on the remaining five wards. Hardy et al. [47] did not conduct statistical significance testing but stated that violence decreased on three wards and increased on two wards. They also noted an increase in verbal abuse on three wards, a decrease on one, and no change on another.

##### Restrictive practices

Both studies reported on restraint and rapid tranquilisation [44,47]. Ellis et al. [44] reported no significant change in levels of incidents requiring restraint or rapid tranquilization overall across the wards. They did report a significant decrease in rapid tranquilization on the two local services admissions wards, but not on the five remaining wards. Hardy et al. [47] did not conduct significance testing but reported an increase in low-level restraint on two wards, a decrease on two, and no change on one. Hardy et al. [47] also noted a reduction in emergency restraint on three wards and an increase on two.

##### Complaints and damage

One study reported on BWCs’ effect on complaints and damage [47]. No statistical significance testing was conducted but they reported that three complaints were made during the BWC pilot period, none of which were related to a particular incident or restraint. They stated that this was lower than in the comparison period the previous year before BWC implementation, where eight complaints were made, two of which had related to an instance of restraint.

#### Global Positioning System (GPS) electronic monitoring

Two studies reported the effect of GPS electronic monitoring technology [50,59]. Neither reported any conflicts of interest. One was rated as medium quality [50] and one low quality [59]. Both had date-matched pre-post study designs.

##### Absconding and leave violations

Both studies reported on absconding and leave violations with GPS electronic monitoring [50,59]. Tully et al. [59] reported that following the introduction of GPS monitoring, there was no significant change in the odds of a leave episode resulting in leave violation during the initial follow-up (one year later). However, during the subsequent follow-up (another year later), leave episodes were significantly less likely to lead to an incident of leave violation. Murphy et al. [50] reported no changes in the overall number of leave violations after implementing GPS electronic monitoring.

##### Complaints and damage

One study reported on complaints relating to GPS electronic monitoring [59]; it reported two events of patients challenging the use of GPS electronic monitoring. It did not report the number of patients involved and number of opportunities to challenge the use of GPS electronic monitoring.

##### Cost-effectiveness

One study reported on the cost-effectiveness of GPS electronic monitoring [50]; it reported a no significant change in the average total cost per patient following the introduction of GPS electronic monitoring.

## Discussion

### Key findings

Our paper has summarised the use of surveillance technologies on inpatient wards internationally, how these technologies are being implemented, staff, patients’ and carers’ views and experiences of them, and the impact these technologies have on quantitative outcomes such as restraint, seclusion, self-harm, violence and aggression, and absconding. There were no randomised controlled trials identified, and very few studies with control groups, meaning that causal inferences regarding the impacts of surveillance technologies cannot be drawn. Overall, there is currently insufficient evidence to suggest that surveillance technologies in inpatient mental health settings are achieving the outcomes they have been employed to achieve.

Key findings regarding implementation included a particular lack of research on certain types of surveillance technologies, such wearable sensors and GPS electronic monitoring, reflecting the novelty of these technologies in inpatient settings. Only two studies specified that they included wards with patients under the age of 18. There was more evidence of implementation of surveillance technologies in the UK than any other individual country. Most of the studies on VBPMM and BWCs were UK-based, indicating an increasing adoption of these technologies in the UK [64]. All of the studies declaring conflicts of interest were examining these technologies, with 4/6 (66.6%) VBPMM studies and 1/4 (25%) BWC studies reporting conflicts of interest.

Our lived experience researchers highlighted discrepancies between the way surveillance technologies were described as being implemented in the literature and their use in practice. For example, they noted that in their experience, staff can decide to view multiple segments of VBPMM video feed instead of it only being viewable when vital sign measurements are made. This underlines the fact that this review only captures how surveillance technologies are described as being implemented in the included papers, and so does not capture the variety of ways in which they may be implemented in practice. Furthermore, it is important to consider that the implementation of surveillance technologies is dynamic, varying across contexts and evolving over time in response to technological innovations and developments in policies and practices.

We identified minimal data regarding ‘best practice’ around the use of surveillance technologies in inpatient settings in the results sections of the included studies. As a result, there is little published evidence from empirical studies that explores such learning and provides ‘guidance’ regarding use on wards. Irrespective of this lack of empirical data, there have been numerous efforts to develop an understanding of what ‘best practice’ could look like given these technologies are already being implemented. Such guidelines have been established by healthcare regulatory bodies, professional associations and charities, as well as internal protocols by specific healthcare providers. This includes guidance around the use of surveillance technologies in general [65,66,67], as well as guidelines and recommendations for specific technologies such as BWCs [12], VBPMM [68,69] and CCTV [70,71,72]. Given the growing use of differing surveillance techniques, further research to explore these guidelines and understand their commonalities and differences (e.g., how best practice may differ across cultures and countries) could provide a better position for developing a more robust message to those institutions implementing them.

Whilst limited data existed regarding ‘best practice’ guidelines, evidence from the papers related to experiences should inform how such practice is developed. Prominent themes in qualitative results were patients’ and staff’s ethical concerns about privacy invasion, data protection, patient confidentiality and informed consent, in-line with previous literature [16,73,74,75,76,77]. These were reinforced by some quantitative evidence indicating that a substantial proportion of patients did not consent to the use of VBPMM [51] or understand the reasons for being monitored via video [48]. Only two studies specified that they included wards with patients under the age of 18, therefore the literature fails to account for the unique ethical considerations when using surveillance technologies within children and young people’s care settings. These findings highlight the danger of surveillance technology use infringing upon patients’ human rights, choice and autonomy. If surveillance technologies are to be implemented in inpatient settings, establishing best practice guidance could potentially help to regulate their use and mitigate some of these adverse effects. However, additional oversight by regulatory bodies to ensure audits of standards and adherence would be required as simply developing and implementing best practice guidelines, standards and policies does not necessarily mean that they will be adhered to in practice. This was exemplified by Warr et al. [61] who highlighted instances in their study where patients were subject to surveillance via CCTV against protocol, at times it was not meant to be in use and with patients who had not consented to its use. Similar concerns are being articulated in lived experience literature [78].

Staff, patient and carer experiences of and attitudes towards surveillance technologies on inpatient wards in the included papers were complex, with variation both within and between these groups. This mirrors findings elsewhere on surveillance technologies [75,76,77]. Qualitative literature in this review revealed some perceptions that surveillance technology could reduce violence, aggression and self-harm in inpatient settings. However, quantitative papers examining these outcomes presented inconsistent or weak results. This finding is consistent with previous systematic reviews reporting a poor and inconsistent evidence base for the use of BWCs in public sector services, including law enforcement, physical and mental healthcare settings [14,21,79,80,81]. This dissonance between qualitative perceptions of surveillance technology in inpatient settings and quantitative evidence is noteworthy; it is unclear whether it is a result of poor-quality evidence, the limitations of the surveillance methods being employed, or the complexity of the issues being addressed through surveillance and the context within which such endeavours take place. It is important to consider that perceptions of surveillance technologies are influenced not only by their effectiveness in practice, but also other external factors. These include, for example, how they are marketed by technology companies and described to people by staff, and broader societal attitudes towards surveillance, particularly amongst those more vulnerable and sensitive to close observation.

A notable discrepancy between the stated aims and the evidence base lies in assertions that surveillance technologies reduce costs [82]. Only two studies in this review explored the cost-effectiveness of surveillance technologies. One found that GPS electronic monitoring use in a forensic inpatient setting did not significantly decrease costs [50], whilst the other reported potential cost savings associated with VBPMM use in PICU settings [49]. These economic analyses had notable limitations, such as only being based on data from single centres and not considering costs such as maintenance, upgrades, wear and tear, staff training and data compliance administration. Downstream costs incurred from the impact of surveillance technologies upon outcomes such as length of inpatient stay, readmission rates and post-discharge service use were also not accounted for. Consequently, the full ongoing costs of implementing surveillance technologies in inpatient mental health settings remains unknown, meaning that claims about their cost-effectiveness are not currently robustly substantiated by the evidence base.

In the one study examining the cost-effectiveness of VBPMM, the main driver of identified potential cost savings was a reduction in one-to-one staff observations [49]. Qualitative evidence suggested that both staff and patients agreed that surveillance technologies should not replace or reduce human interaction. Indeed, research suggests that human contact, trust, support and empowerment form integral elements of therapeutic inpatient care, including during episodes of containment such as seclusion and restraint [15,19]. Malcolm et al. [49] argue that a reduction in one-to-one staff observations with VBPMM could potentially free-up resources which could be used on other, more therapeutically beneficial activities. However, in practice, there is no guarantee that this freed-up staff time may not be used for these purposes, leading to a reduction in therapeutic interaction between staff and patients [83]. There is therefore a risk that the use of surveillance technologies to reduce staffing costs could result in decreased human interaction and so quality of care in inpatient settings.

Qualitative findings revealed that staff, patient and carer perceptions and experiences were mixed across the surveillance technology types. Some of the perceived benefits of surveillance technologies included: improved staff and patient safety, enhanced monitoring and prevention of incidents (e.g., absconding, self-harm, violence and aggression), and facilitation of less intrusive observations of patients. Providing evidence to help investigate incidents and complaints was another perceived benefit, although some noted that surveillance technologies do not necessarily capture the entirety of events (e.g., due to some being turned on and off at the discretion of staff, and because they may not capture all of the events leading up to an incident). Concerns were also expressed by staff and patients that surveillance technology use could have wide-ranging negative effects, including negatively impacting patients’ recovery, privacy and dignity, decreasing feelings of safety, exacerbating distress and paranoia, reducing quality of care, damaging therapeutic relationships with staff and exacerbating power imbalances between patients and staff. Indeed, patient and service user groups, along with advocates and disability activists, have consistently voiced concerns about the potential iatrogenic harms associated with the use of surveillance technology in inpatient mental health settings [84,85]. These harms have been the subject of media attention [30,86] including recent inquest reports suggesting that “alarm fatigue” associated with surveillance technology use can even have fatal consequences [86].

However, many of the included studies did not comprehensively investigate potential impacts, including unintended consequences, quantitatively. For example, very few quantitatively investigated surveillance technology’s impact upon patients’ mental health, absconding rates, self-harm, or care quality. Further, even when these outcomes were investigated, there may have been limitations in how they were measured. For example, Ndebele et al. [51] only measured self-harm frequency in bedrooms and bathrooms, and so they did not capture any possible impact of VBPMM on rates of self-harm in communal ward spaces or on self-harm severity. This is a concern, given reports from patients that VBPMM use can worsen self-harm [78]. Many possible effects were not investigated at all in any of the included studies, such as the impact of surveillance technologies on therapeutic alliances, treatment satisfaction, staff and patient well-being, patient quality of life, recovery, engagement with services, and longer-term outcomes such as readmission rates and post-discharge mental health and service use. Therefore, this review shows that our understanding of the impact of surveillance technologies in inpatient mental health settings, including their full range of potential harms and risks, remains incomplete.

### Methodological quality of the included studies

There were significant methodological limitations in more than two fifths (44.4%) of included studies. Furthermore, there were declared conflicts of interest in nearly a fifth of studies (18.5%), all in studies examining VBPMM and BWCs, and additional potential undisclosed conflicts of interest identified. We noted that several of the studies with positive findings had declared conflicts of interest relating to the technology of interest, for example, studies being funded or conducted by the technology company themselves. This may not be surprising given their drive to demonstrate the efficacy of their technology. Many of these studies were also rated as low quality. Results therefore need to be interpreted with caution.

There was often a lack of information about how participants were recruited, and how surveys and interviews or focus groups were conducted, making it difficult to assess potential biases (e.g., risk of cherry-picking participants, excluding the most unwell patients, power imbalances inhibiting sharing of criticisms of technology by patients and staff). Consequently, the literature may underrepresent the perspectives of populations facing greater barriers to research participation (e.g., patients lacking capacity to consent, people with concerns about confidentiality, distrust towards research or facing language barriers) [88]. The lack of transparency in methodologies, e.g., no pre-registration of studies, makes it difficult to ascertain how reported outcomes were chosen, and raises questions around whether negative outcomes (such as harms, verbal aggression and property destruction) were purposefully omitted. Methodologically, no randomised controlled trials were identified, and few studies had control groups, with mainly before and after comparisons. Many papers did not adequately consider the complexity of the issues and variables surrounding surveillance, for instance, the role of confounding or contextual factors in interpreting results.

There was in general a significant lack of lived experience involvement in the implementation and evaluation of surveillance technologies, and a lack of lived experience involvement in the studies themselves. Even when it was reported, it was often poorly described, for example, lacking detail about numbers of people involved, their demographics, recruitment methods and how (and to what degree) they were involved in the research process. Furthermore, in some studies there lacked a clear distinction between the involvement of individuals with lived experience in the research process versus participation in the study by patients.

### Strengths and limitations

Our review is a comprehensive, systematic synthesis of the available literature on the implementation, experience, and impact of surveillance technologies in inpatient mental health settings. We reported information on lived experience involvement in the study design and the implementation of the surveillance, exposing significant gaps which should be addressed and prioritised. We also reported information on declared conflicts of interest and funding in the included papers, which have enhanced our ability to assess the validity and independence of the evidence presented.

We sought to identify both academic and grey literature in our review, although, due to time constraints, grey literature was only included in relation to RQ2a (exploring patient, staff and carer perceptions and experiences of the technology) and was limited to studies which included a description of their methodology. We acknowledge that there may be perspectives which are therefore underrepresented in our synthesis, including perspectives from those with lived experience of surveillance on inpatient wards. There is a risk of publication bias (i.e., studies showing positive outcomes being more likely to be published) given the number of included studies which declared conflicts of interest, although we were unable to investigate and confirm this.

### Implications for policy and practice

The findings of this review suggest that the current evidence base does not support the use of surveillance technologies as a means of improving safety, care quality or reducing costs in inpatient mental health settings.

More independent, coproduced research is needed thoroughly evaluating the impact of surveillance technologies, including their full range of potential harms, in inpatient settings. As is best practice with the implementation of any new intervention, they should only be deployed if the resulting evidence supports their use.

However, the current reality is that surveillance technologies are already being implemented across a variety of inpatient services across the globe, and it is unlikely that this will come to a complete halt. If these technologies continue to be implemented, there will be an urgent need to develop trauma-informed policies, procedures and guidelines for their use, centred around the perspectives of patients. This could contribute to developing more acceptable ways of using surveillance technologies and help maximise their potential benefits and mitigate their harms [89]. These guidelines and policies would need to be accompanied by comprehensive and ongoing training for staff, ideally coproduced with patients, and systematic monitoring and auditing of services’ adherence to them to help ensure compliance.

These policies and guidelines should comprehensively address the tensions and ethical concerns highlighted by patients, carers, and staff in this review. This includes concerns around informed consent, patient confidentiality, data protection and potential iatrogenic harms. Procedures for investigating and addressing misuse of technology and data should be incorporated. Wider systemic challenges, including issues such as staffing shortages, power imbalances and reliance on restrictive approaches to risk management, also need to be acknowledged and actively addressed.

It is essential that all stakeholders, particularly patients, are meaningfully involved in all stages of future research, implementation, evaluation and decision-making regarding surveillance technology use in inpatient mental health settings.

### Implications for research

The literature base identified in this review is largely characterised by uncontrolled and poor-quality studies presenting inconsistent results. Nearly a fifth of papers identified in this review had declared conflicts of interest, and additional potential undisclosed conflicts of interest were also identified.

Future research on surveillance in inpatient wards should be funded and conducted independently to ensure the rigour and validity of the methods and findings. Conflicts of interest should also be declared in any published reports or articles. Research led by those with lived experience of mental health inpatient care generally, and surveillance technologies specifically, would be particularly valuable in evaluating potential harms missed by academic or clinical researchers. Care should also be taken to ensure that the perspectives of those who are unwell, or may need support to express their views, are captured in any future research on technologies in these settings [90]. Further synthesis of data on surveillance from other locations where people with mental health problems present may be helpful, for example in crisis services or mental health presentations in emergency departments.

Future primary research in this area should more purposefully aim to: i) investigate the harms caused by surveillance, including a full exploration of the psychological impact and an exploration of changes in care protocols due to the technology, ii) explore and establish best practice and ethical guidelines for the use of surveillance in inpatient units (and across all mental health services and settings) which fully consider the experiences of patients who have negative views and adverse responses to surveillance, and iii) include those with lived experience in study design, analysis, interpretation, and dissemination.

### What is already known on this topic

- Surveillance technologies are increasingly being implemented in inpatient mental health settings, with the stated aim of improving safety, though their use is controversial.
- This is the first systematic review of the evidence on the implementation, experiences and effects of a range of surveillance technologies in inpatient mental health settings.

### What this study adds

- The findings of this review suggest that the current evidence base does not support the use of surveillance technologies as a means of improving safety, care quality or reducing costs in inpatient mental health settings.
- Patient, staff and carer perceptions and experiences were mixed across the surveillance technology types.
- Further independent, co-produced research is needed to thoroughly evaluate their impact, including their full range of potential harms, in inpatient mental health settings.

### Lived experience commentary, by Georgia Johnson and Rachel Rowan Olive

We are unsurprised by the poor quality and inconsistent results of the evidence. In our experience surveillance technology – like most restrictive practice – is rapidly rolled out in response to institutional anxiety following serious incidents. Surveillance technology’s illusion of control alleviates that anxiety, promising potential benefits well beyond the evidence base. Surveillance’s damage, however, is more concrete. Most researchers did not look for iatrogenic harm, thus compounding said harm by invalidating our fears and experiences.

But we know these harms intimately, because we have experienced them. These digital technologies strip away our most basic dignity, and are, by an extension, an affront to our very humanity. It is when professionals stop treating us like humans, and see only a cluster of symptoms, that restrictive practice becomes its most abusive self. Other people’s fear is not a justification for abusing us in this way.

The UK’s psychiatric system is not one where meaningful consent for surveillance can be implemented, however blithely manufacturers and evaluators state that consent is always obtained. When Oxevision was piloted on Georgia’s ward, she was not given the opportunity to consent: she only discovered the system existed after a nurse said, “Oh you’re in the bathroom, I couldn’t see you on the camera.” Staff didn’t know whether patients were allowed to refuse it. The distress caused was so great that the response team had to be called. After turning the cameras off, they were turned back on during another shift. When Georgia objected, staff said that no such cameras existed and that she was experiencing psychosis. *Would she like a cup of tea instead?* FOI data from StopOxevision [91] shows this is not an isolated event, with patient leaflets and posters frequently omitting any mention of functionalities such as camera surveillance.

Finally, we highlight the contrast in attitudes to staff surveilling patients versus patients filming staff. On being illegally detained during a mental health crisis, Rachel began recording those detaining her, knowing we are frequently disbelieved when making complaints. Outraged staff *wearing body-worn cameras* promptly insisted, “we are not here to be filmed”.

This is a common response to patients documenting poor experiences; it puts paid to any illusion that institutional surveillance could lessen the violent disbelief we face. Staff control when and how cameras are used. Surveillance within this system only cements power imbalances and causes lasting trauma.

## Acknowledgements

We are very appreciative of the NIHR MHPRU Lived Experience Researchers who contributed to and supported this work (study design, screening, data extraction, quality appraisal, synthesis, feeding back on drafts of the paper).

## Conflicts of interest

AS and UF have undertaken and published research on BWCs. We have received no financial support from BWC or any other surveillance technology companies. All other authors declare no competing interests.

## Funding

This study is funded by the National Institute for Health and Care Research (NIHR) Policy Research Programme (grant no. PR-PRU-0916-22003). The views expressed are those of the author(s) and not necessarily those of the NIHR or the Department of Health and Social Care. The funders had no role in study design, data collection and analysis, decision to publish, or preparation of the manuscript. ARG was supported by the Ramon y Cajal programme (RYC2022-038556-I), funded by the Spanish Ministry of Science, Innovation and Universities.

## Data availability statement

The template data extraction form is available in Supplementary 1. MMAT quality appraisal ratings for each included study are available in Supplementary 2. All data used is publicly available in the published papers included in this review.

## Author contribution

All authors substantially contributed to the conception or design of this study. Literature searching was conducted by JG and KS. Title and abstract screening was conducted by KS, UF, JG, AG, CR. Full text screening was conducted by KS, UF, JG, AG, CR, RC. Data extraction and quality appraisal were conducted by KS, JG, AG, RC, UF, CR. Evidence synthesis was led by JG and UF and supported by all other authors. JG, KS and UF led on drafting the manuscript with input and/or editing by all authors. All authors read and approved the final manuscript.

## Acronyms

BWCs: Body Worn Cameras
CCTV: Closed Circuit Television
CI: Confidence Interval
GPS: Global Positioning System
IT: Information Technology
MHPRU: Policy Research Unit in Mental Health
MMAT: Mixed Methods Appraisal Tool
NHS: National Health Service
NIHR: National Institute for Health and Social Care Research
PANSS: Positive and Negative Syndrome Scale
PICU: Psychiatric Intensive Care Unit
PIN: Personal Identification Number
PMVA: Prevention and Management of Violence and Aggression
PRISMA: Preferred Reporting Items for Systematic Reviews and Meta-Analyses
SD: Standard Deviation
TV: Television
UK: United Kingdom
USA: United States of America
VBPMM: Vision-Based Patient Monitoring and Management

## Notes

### Clinical Protocols

https://www.crd.york.ac.uk/prospero/display_record.php?RecordID=463993

