## Appendices for "The use and impact of surveillance-based technology initiatives in inpatient and acute mental health settings: A systematic review"

**Appendix A:** Preferred Reporting Items for Systematic Reviews and Meta-Analyses (PRISMA) checklist

| Section and Topic | Item # | Checklist item | Location where item is reported |
|---|---|---|---|
| **TITLE** | | | |
| Title | 1 | Identify the report as a systematic review. | Page 1 |
| **ABSTRACT** | | | |
| Abstract | 2 | See the PRISMA 2020 for Abstracts checklist. | Page 2 |
| **INTRODUCTION** | | | |
| Rationale | 3 | Describe the rationale for the review in the context of existing knowledge. | Pages 3-4 |
| Objectives | 4 | Provide an explicit statement of the objective(s) or question(s) the review addresses. | Page 4 |
| **METHODS** | | | |
| Eligibility criteria | 5 | Specify the inclusion and exclusion criteria for the review and how studies were grouped for the syntheses. | Pages 5-7 |
| Information sources | 6 | Specify all databases, registers, websites, organisations, reference lists and other sources searched or consulted to identify studies. Specify the date when each source was last searched or consulted. | Page 5 |
| Search strategy | 7 | Present the full search strategies for all databases, registers and websites, including any filters and limits used. | Appendix B |
| Selection process | 8 | Specify the methods used to decide whether a study met the inclusion criteria of the review, including how many reviewers screened each record and each report retrieved, whether they worked independently, and if applicable, details of automation tools used in the process. | Page 5 |
| Data collection process | 9 | Specify the methods used to collect data from reports, including how many reviewers collected data from each report, whether they worked independently, any processes for obtaining or confirming data from study investigators, and if applicable, details of automation tools used in the process. | Page 7 |
| Data items | 10a | List and define all outcomes for which data were sought. Specify whether all results that were compatible with each outcome domain in each study were sought (e.g. for all measures, time points, analyses), and if not, the methods used to decide which results to collect. | Page 6-7 and Supplementary 1 |
| | 10b | List and define all other variables for which data were sought (e.g. participant and intervention characteristics, funding sources). Describe any assumptions made about any missing or unclear information. | Supplementary 1 |
| Study risk of bias assessment | 11 | Specify the methods used to assess risk of bias in the included studies, including details of the tool(s) used, how many reviewers assessed each study and whether they worked independently, and if applicable, details of automation tools used in the process. | Page 7 |
| Effect measures | 12 | Specify for each outcome the effect measure(s) (e.g. risk ratio, mean difference) used in the synthesis or presentation of results. | Page 8 |
| Synthesis methods | 13a | Describe the processes used to decide which studies were eligible for each synthesis (e.g. tabulating the study intervention characteristics and comparing against the planned groups for each synthesis (item #5)). | Page 8 |
| | 13b | Describe any methods required to prepare the data for presentation or synthesis, such as handling of missing summary statistics, or data | Page 8 |

| Section and Topic | Item # | Checklist item | Location where item is reported |
|---|---|---|---|
| | | conversions. | |
| | 13c | Describe any methods used to tabulate or visually display results of individual studies and syntheses. | Page 8 |
| | 13d | Describe any methods used to synthesize results and provide a rationale for the choice(s). If meta-analysis was performed, describe the model(s), method(s) to identify the presence and extent of statistical heterogeneity, and software package(s) used. | Page 8 |
| | 13e | Describe any methods used to explore possible causes of heterogeneity among study results (e.g. subgroup analysis, meta-regression). | Page 8 |
| | 13f | Describe any sensitivity analyses conducted to assess robustness of the synthesized results. | N/A |
| Reporting bias assessment | 14 | Describe any methods used to assess risk of bias due to missing results in a synthesis (arising from reporting biases). | Pages 7-8 |
| Certainty assessment | 15 | Describe any methods used to assess certainty (or confidence) in the body of evidence for an outcome. | N/A |
| **RESULTS** | | | |
| Study selection | 16a | Describe the results of the search and selection process, from the number of records identified in the search to the number of studies included in the review, ideally using a flow diagram. | Page 8-10 |
| | 16b | Cite studies that might appear to meet the inclusion criteria, but which were excluded, and explain why they were excluded. | Appendix C |
| Study characteristics | 17 | Cite each included study and present its characteristics. | Pages 11-24 and Appendix D |
| Risk of bias in studies | 18 | Present assessments of risk of bias for each included study. | Pages 11-24 and Supplementary 2 |
| Results of individual studies | 19 | For all outcomes, present, for each study: (a) summary statistics for each group (where appropriate) and (b) an effect estimate and its precision (e.g. confidence/credible interval), ideally using structured tables or plots. | Pages 59-74 |
| Results of syntheses | 20a | For each synthesis, briefly summarise the characteristics and risk of bias among contributing studies. | Tables 3-10. Narrative descriptions provided throughout the results section. |
| | 20b | Present results of all statistical syntheses conducted. If meta-analysis was done, present for each the summary estimate and its precision (e.g. confidence/credible interval) and measures of statistical heterogeneity. If comparing groups, describe the direction of the effect. | N/A |
| | 20c | Present results of all investigations of possible causes of heterogeneity among study results. | N/A |

| Section and Topic | Item # | Checklist item | Location where item is reported |
|---|---|---|---|
| | 20d | Present results of all sensitivity analyses conducted to assess the robustness of the synthesized results. | N/A |
| Reporting biases | 21 | Present assessments of risk of bias due to missing results (arising from reporting biases) for each synthesis assessed. | N/A |
| Certainty of evidence | 22 | Present assessments of certainty (or confidence) in the body of evidence for each outcome assessed. | N/A |
| **DISCUSSION** | | | |
| Discussion | 23a | Provide a general interpretation of the results in the context of other evidence. | Pages 75-79 |
| | 23b | Discuss any limitations of the evidence included in the review. | Pages 75-79 |
| | 23c | Discuss any limitations of the review processes used. | Page 80 |
| | 23d | Discuss implications of the results for practice, policy, and future research. | Pages 80-82 |
| **OTHER INFORMATION** | | | |
| Registration and protocol | 24a | Provide registration information for the review, including register name and registration number, or state that the review was not registered. | Page 4 |
| | 24b | Indicate where the review protocol can be accessed, or state that a protocol was not prepared. | Page 4 |
| | 24c | Describe and explain any amendments to information provided at registration or in the protocol. | N/A |
| Support | 25 | Describe sources of financial or non-financial support for the review, and the role of the funders or sponsors in the review. | Pages 83-84 |
| Competing interests | 26 | Declare any competing interests of review authors. | Page 83 |
| Availability of data, code and other materials | 27 | Report which of the following are publicly available and where they can be found: template data collection forms; data extracted from included studies; data used for all analyses; analytic code; any other materials used in the review. | Page 84 |

**<u>Appendix B:</u>** Full search strategies

**Academic databases:**

<u>Embase (2358 results), MEDLINE (946 results) and PsycInfo (524 results) search terms:</u>
(Psychiatri* or Mental health).mp. [mp=title, abstract, heading word, drug trade name, original title, device manufacturer, drug manufacturer, device trade name, keyword heading word, floating subheading word, candidate term word]

AND

(Inpatient or Hospital* or Ward* or Unit or PICU or 136-suite or 136 suite or place* of safety).mp. [mp=title, abstract, heading word, drug trade name, original title, device manufacturer, drug manufacturer, device trade name, keyword heading word, floating subheading word, candidate term word]

AND

(Surveill* or CCTV or Body worn camera or bodycam* or body camera or body-cam or body worn video or remote technolog* or digitally assisted or tracking device* or video track* or door-lock* or non-contact monitor* or sensor-based monitor* or electronic monitor* or patient monitor* or video monitor* or vision-based monitor*).mp. [mp=title, abstract, heading word, drug trade name, original title, device manufacturer, drug manufacturer, device trade name, keyword heading word, floating subheading word, candidate term word]

<u>Pubmed (567 results) search terms:</u>
((Psychiatri*[Title/Abstract] OR (Mental health[Title/Abstract])) AND (Inpatient[Title/Abstract] OR Hospital*[Title/Abstract] OR Ward*[Title/Abstract] OR Unit[Title/Abstract] OR PICU[Title/Abstract] OR (136-suite[Title/Abstract]) OR (136 suite[Title/Abstract]) OR (place* of safety[Title/Abstract]))) AND (Surveill*[Title/Abstract] OR CCTV[Title/Abstract] OR (Body worn camera[Title/Abstract]) OR bodycam*[Title/Abstract] OR (body camera[Title/Abstract]) OR (body-cam[Title/Abstract]) OR (body worn video[Title/Abstract]) OR (remote technolog*[Title/Abstract]) OR (digitally assisted[Title/Abstract]) OR (tracking device*[Title/Abstract]) OR (video track*[Title/Abstract]) OR (door-lock*[Title/Abstract]) OR (non-contact monitor*[Title/Abstract]) OR (sensor-based monitor*[Title/Abstract]) OR (electronic monitor*[Title/Abstract]) OR (patient monitor*[Title/Abstract]) OR (video monitor*[Title/Abstract]) OR (vision-based monitor*[Title/Abstract]))system*[Title/Abstract]) OR Oxehealth[Title/Abstract] OR (tracking device*[Title/Abstract]) OR (video track*[Title/Abstract]) OR (door-lock*[Title/Abstract]))

<u>Scopus (2497) search terms:</u>
TITLE-ABS-KEY ( psychiatri* OR "Mental health" ) AND TITLE-ABS-KEY ( inpatient OR hospital* OR ward* OR unit OR picu OR "136-suite" OR "136 suite" OR "place* of safety" ) AND TITLE-ABS-KEY ( surveill* OR cctv OR "Body worn camera" OR bodycam* OR "body camera" OR "body-cam" OR "body worn video" OR "remote technolog*" OR "digitally assisted" OR "tracking device*" OR "video track*" OR "door-lock*" OR "non-contact

monitor*" OR "sensor-based monitor*" OR "electronic monitor*" OR "patient monitor*" OR "video monitor*" OR "vision-based monitor*" )

**Total references extracted from academic databases:**

All: 6892
Embase: 2358
Medline: 946
PsycInfo: 524
Scopus: 2497
Pubmed: 567
After de-duplication on Endnote: 3385

**Grey literature searching**

PsyArXiv (251 results)
(Psychiatri* OR “Mental health”) AND (Inpatient OR Hospital* OR Ward* OR Unit OR PICU OR 136-suite OR 136 suite OR place* of safety) AND (Surveill* or CCTV or Body worn camera or bodycam* or body camera or body-cam or body worn video or remote technolog* or digitally assisted or tracking device* or video track* or door-lock* or non-contact monitor* or sensor-based monitor* or electronic monitor* or patient monitor* or video monitor* or vision-based monitor* or video surveillance or remote monitor* or tracking device*)

MedRxiv (261 results)
Psychiatri* OR “Mental health” AND Inpatient AND Surveill* or CCTV or "Body worn camera" or non-contact monitor* or video track*

HMIC (263 results)
HMIC Health Management Information Consortium <1979 to July 2023>

| | | |
|---|---|---|
| 1 | exp Mental health/ or Psychiatr*.mp. or exp mental disorders/ | 33984 |
| 2 | mental health.mp. or exp Mental health/ | 26004 |
| 3 | 1 or 2 | 43925 |
| 4 | Inpatient.mp. or exp In patients/ | 4299 |
| 5 | Hospital.mp. or exp Mental health hospitals/ or exp Psychogeriatric hospitals/ | 46272 |
| 6 | exp Hospital wards/ or exp Childrens wards/ or Ward.mp. or exp wards/ | 5118 |
| 7 | Unit.mp. | 9822 |
| 8 | 136-suite.mp. | 2 |
| 9 | 136 suite*.mp. | 2 |
| 10 | Place* of safety.mp. | 98 |
| 11 | Surveill*.mp. | 3131 |
| 12 | exp psychiatric units/ or exp Closed circuit television/ or exp Security systems/ or cctv.mp. | 881 |
| 13 | Body worn camera*.mp. | 0 |
| 14 | bodycam*.mp. | 0 |
| 15 | body camera.mp. or exp Monitoring systems/ | 471 |

16 body-cam.mp. 0
17 body worn video.mp. 0
18 remote technolog*.mp. 5
19 digitally assisted.mp. 0
20 exp Assistive technology/ or exp Health technology/ or tracking device*.mp. or exp Monitoring systems/ 1613
21 video track*.mp. 0
22 door-lock*.mp. 9
23 non-contact monitor*.mp. 0
24 sensor-based monitor*.mp. 0
25 electronic monitor*.mp. 22
26 patient monitor*.mp. or exp Patient monitoring equipment/ 289
27 video monitor*.mp. 3
28 vision-based monitor*.mp. 0
29 video surveillance.mp. 11
30 remote monitor*.mp. 56
31 tracking device*.mp. 5
32 picu.mp. 61
33 4 or 5 or 6 or 7 or 8 or 9 or 10 or 32 56717
34 11 or 12 or 13 or 14 or 15 or 16 or 17 or 18 or 19 or 20 or 21 or 22 or 23 or 24 or 25 or 26 or 27 or 28 or 29 or 30 or 31 5694
35 3 and 33 and 34263
36 exp Mental health/ or Psychiatr*.mp. or exp mental disorders/ 33984
37 mental health.mp. or exp Mental health/ 26004
38 36 or 37 43925
39 Inpatient.mp. or exp In patients/ 4299
40 Hospital.mp. or exp Mental health hospitals/ or exp Psychogeriatric hospitals/ 46272
41 exp Hospital wards/ or exp Childrens wards/ or Ward.mp. or exp wards/ 5118
42 Unit.mp. 9822
43 136-suite.mp. 2
44 136 suite*.mp. 2
45 Place* of safety.mp. 98
46 Surveill*.mp. 3131
47 exp psychiatric units/ or exp Closed circuit television/ or exp Security systems/ or cctv.mp. 881
48 Body worn camera*.mp. 0
49 bodycam*.mp. 0
50 body camera.mp. or exp Monitoring systems/ 471
51 body-cam.mp. 0
52 body worn video.mp. 0
53 remote technolog*.mp. 5
54 digitally assisted.mp. 0
55 exp Assistive technology/ or exp Health technology/ or tracking device*.mp. or exp Monitoring systems/ 1613
56 video track*.mp. 0

57 door-lock*.mp. 9
58 non-contact monitor*.mp. 0
59 sensor-based monitor*.mp. 0
60 electronic monitor*.mp. 22
61 patient monitor*.mp. or exp Patient monitoring equipment/ 289
62 video monitor*.mp. 3
63 vision-based monitor*.mp. 0
64 video surveillance.mp. 11
65 remote monitor*.mp. 56
66 tracking device*.mp. 5
67 picu.mp. 61
68 39 or 40 or 41 or 42 or 43 or 44 or 45 or 67 56717
69 46 or 47 or 48 or 49 or 50 or 51 or 52 or 53 or 54 or 55 or 56 or 57 or 58 or 59 or 60 or 61 or 62 or 63 or 64 or 65 or 66 5694
70 38 and 68 and 69 263

**Appendix C:** Excluded full texts and reasons for exclusion

| Reference | Reason for exclusion | Full Reference |
|---|---|---|
| 1. Alezrah et al. (2018) | Wrong design | Alezrah C, Lafont M, Bouet R, Depaigne-Loth A, Paindavoine C, Laurence M, Rodde-Dunet MH, Senon JL. Les dernières recommandations de bonnes pratiques sur l'isolement et la contention [The latest best practice guidelines relating to seclusion and restraint]. Soins Psychiatr. 2018 Jul-Aug;39(317):16-19. French. doi: 10.1016/j.spsy.2018.04.005. PMID: 30047452. |
| 2. Arnetz et al. (2011) | Not a surveillance tech | Arnetz JE, Aranyos D, Ager J, Upfal MJ. Development and application of a population-based system for workplace violence surveillance in hospitals. Am J Ind Med. 2011 Dec;54(12):925-34. doi: 10.1002/ajim.20984. Epub 2011 Jul 7. PMID: 21739469. |
| 3. Baker (2004) | No access | Baker, S. (2004). Holding Psychiatric Patients. *Journal of Emergency Nursing, 30 (1),* 73-75. |
| 4. Banja (2017) | Wrong design | Banja JD. Failures of foreseeability: Risk management considerations in reducing allegations of sexual violence in psychiatric units. J Healthc Risk Manag. 2017 Jan;36(3):21-25. doi: 10.1002/jhrm.21238. PMID: 28099795. |
| 5. Borges & Nourani-Vatani (2011) | No usable data | Borges PV, Nourani-Vatani N. Vision-based detection of unusual patient activity. Stud Health Technol Inform. 2011;168:16-23. PMID: 21893907. |
| 6. Bowers et al (2010) | Not a surveillance tech | Bowers, L., Haglund, K., Muir-Cochrane, E., Nijman, H., Simpson, A. & Van Der Merwe, M. (2010), Locked doors: a survey of patients, staff and visitors. Journal of Psychiatric and Mental Health Nursing, 17: 873-880. https://doi.org/10.1111/j.1365-2850.2010.01614.x |
| 7. Bowers et al. (2008) | Not a surveillance tech | Bowers, L., Allan, T., Haglund, K., Muir-Cochrane, E., Nijman, H., Simpson, A. & Van Der Merwe, M. (2008). The City-128 extension: locked doors in acute psychiatry, outcome and acceptability. Available at: https://njl-admin.nihr.ac.uk/document/download/2027549 [Accessed 18/03/24]. |
| 8. Cheng et al. (2013) | No usable data | Cheng, C-Y., Lan, T-H, & Chan, C-H. (2013). An improved localization algorithm with wireless heartbeat monitoring system or patient safety in psychiatric wards. *Engineering Applications of Artificial Intelligence, 26 (2),* 905-912. |
| 9. Cheng et al. (2019) | No usable data | K. Cheng, M. S. Khokhar, Q. Liu, R. Tahir and M. Li, "Data-Driven Logical Topology Inference for Managing Safety and Re-Identification of Patients Through Multi-Cameras IoT," in IEEE Access, vol. 7, pp. 159466-159478, 2019, doi: 10.1109/ACCESS.2019.2951164. |

| | | |
|---|---|---|
| 10. Curtis et al. (2007) | Not a surveillance tech | Curtis, S., Gesler, W., Fabian, K., Francis, S., & Priebe, S. (2007). Therapeutic Landscapes in Hospital Design: A Qualitative Assessment by Staff and Service Users of the Design of a New Mental Health Inpatient Unit. Environment and Planning C: Government and Policy, 25(4), 591-610. https://doi.org/10.1068/c1312r |
| 11. Desai (2010) | Wrong design | Desai, S. (2010). Violence and Surveillance: Some Unintended Consequences of CCTV Monitoring within Mental Health Hospital Wards. Surveillance & Society 8(1): 85-92. |
| 12. Desai (2011) | Wrong design | Desai, S. (2011). Violence and surveillance in mental health wards. *Criminal Justice Matters, 83 (1),* 4-5. |
| 13. Efkemann et al. (2019) | Not a surveillance tech | Efkemann SA, Bernard J, Kalagi J, Otte I, Ueberberg B, Assion HJ, Zeiß S, Nyhuis PW, Vollmann J, Juckel G, Gather J. Ward Atmosphere and Patient Satisfaction in Psychiatric Hospitals With Different Ward Settings and Door Policies. Results From a Mixed Methods Study. Front Psychiatry. 2019 Aug 30;10:576. doi: 10.3389/fpsyt.2019.00576. PMID: 31543830; PMCID: PMC6728825. |
| 14. Esc et al. (1979) | Wrong setting | Esc, E.M., Gayral, L.F. & Goldberger, E. (1979). Handling the emergency psychiatric case at the general hospital. *Revue de Medicine de Toulouse, 15 (0),* 293-298. |
| 15. Frank (2013) | Wrong design | Frank,U. (2013). Video monitoring in psychiatry: pro and contra. Psychiatr Prax, 40:117–118 |
| 16. Goerdeler (2014) | No access | Goerdeler, J. (2014). Data Protection in forensic psychiatric institutions. *Recht und Psychiatrie, 32 (3),* 129-141. |
| 17. Haglund et al. (2006) | Not a surveillance tech | Haglund K, von Knorring L, von Essen L. Psychiatric wards with locked doors--advantages and disadvantages according to nurses and mental health nurse assistants. J Clin Nurs. 2006 Apr;15(4):387-94. doi: 10.1111/j.1365-2702.2006.01489.x. PMID: 16553751. |
| 18. Hall (2004) | Not a surveillance tech | Hall JE. Restriction and control: the perceptions of mental health nurses in a UK acute inpatient setting. Issues Ment Health Nurs. 2004 Jul-Aug;25(5):539-52. doi: 10.1080/01612840490443473. PMID: 15204895. |
| 19. Hutton et al. (2021) | Not a surveillance tech | Hutton A, Wilson R, Foureur M. Comfort Equals Nurturing: Young People Talk About Mental Health Ward Design. HERD: Health Environments Research & Design Journal. 2021;14(4):258-269. doi:10.1177/19375867211022684 |
| 20. Igoumenou (2020) | Wrong design | Igoumenou, A. (2020). Ethical Issues in Clinical Forensic Psychiatry. Springer Nature: Swizerland. |
| 21. Jehel et al. (2010) | Wrong design | Jehel L, Simon G. Critères d'évaluation et de surveillance pour la prise en charge de la crise suicidaire [Criteria for evaluation and surveillance in managing a suicidal crisis]. Soins Psychiatr. 2010 Jan-Feb;(266):37-41. French. PMID: 20162967. |
| 22. Kmietowicz (2017) | Wrong design | Kmietowicz, Z. (2017). Body cameras on staff reduced attacks in psychiatric wards, finds study. *BMJ,* 357 doi: https://doi.org/10.1136/bmj.j2268 |

| | | |
|---|---|---|
| 23. Kroll et al. (2020) | Wrong setting | Kroll, L., Böhning, N., Müßigbrodt, H. *et al.* Non-contact monitoring of agitation and use of a sheltering device in patients with dementia in emergency departments: a feasibility study. *BMC Psychiatry* 20, 165 (2020). https://doi.org/10.1186/s12888-020-02573-5 |
| 24. Li et al. (20160 | Wrong design | Li, C.Y., Lee, Y.H., You, C.W., Huang, M.C., Chuang, Y., Chu, H.H., Lin, Y.F., Chen. L.L. (2016). Challenges and opportunities for designing new technology to reveal early warning signs in acute psychiatry units. *UbiComp – Proceedings for the 2016 ACM International Joint Conference on Pervasive and Ubiquitous Computing,* 145-148. |
| 25. Lusignan (1998) | No access | Lusignan, R. (1998). Victimization of clinicians in psychiatric settings. *American Journal of Forensic Psychiatry 19,* 53-66. |
| 26. Male et al. (1991) | No usable data | Male, B., Clark, C. & El Komy, A. (1991). Electronic alert system for mentally handicapped adults incapable of consent: civilise technology or civil rights abuse. *Psychiatric Bulletin, 15, 605-606.* |
| 27. McClean et al. (2011) | Not a surveillance tech | McClean RJ. Assessing the security needs of patients in medium secure psychiatric care in Northern Ireland. *The Psychiatrist*. 2010;34(10):432-436. doi:10.1192/pb.bp.109.027672 |
| 28. Missouridou et al. (2021) | Not a surveillance tech | Missouridou, E., Resoulai, A., Sakavara, I., Fradelos, E.C., Kritsiotakis, E., Mangoulia, P., Kasidi, E., Stefanou, E., Liapis, C., Segredou, E., Koutelekos, J. & Evagelou, E. (2021). Psychiatric Care in Acute Care Units with Locked Doors: nursing care providers' perceptions and experiences. *Advances in Experimental Medicine and Biology, 1337,* 99-106. |
| 29. Missouridou et al. (2020) | Not a surveillance tech | Missouridou E , Zartaloudi A , Dafogianni C , et al. Locked versus open ward environments and restrictive measures in acute psychiatry in Greece: Nursing students' attitudes and experiences. *Perspect Psychiatr Care*. 2021; 57: 1365-1375. https://doi.org/10.1111/ppc.12699 |
| 30. Morris (2021) | Wrong design | Morris, N.P. (2021). Digital Technologies and Coercion in Psychiatry. Psychiatr Serv. 2021 Mar 1;72(3):302-310. doi: 10.1176/appi.ps.202000427. PMID: 33430653. |
| 31. Muir-Cochrane et al. (20120 | Not a surveillance tech | Muir-Cochrane, E., van der Merwe, M., Nijman, H., Haglund, K., Simpson, A. and Bowers, L. (2012), Investigation into the acceptability of door locking to staff, patients, and visitors on acute psychiatric wards. International Journal of Mental Health Nursing, 21: 41-49. https://doi.org/10.1111/j.1447-0349.2011.00758.x |
| 32. Nolan et al. (2006) | Wrong design | Nolan KA, Volavka J. Video recording in the assessment of violent incidents in psychiatric hospitals. J Psychiatr Pract. 2006 Jan;12(1):58-63. doi: 10.1097/00131746-200601000-00010. PMID: 16432448. |

| | | |
|---|---|---|
| 33. Novitsky et al. (2009) | Wrong design | Novitsky, M.A., Julius, R.J. & Dubin, W.R. (2009). Non-pharmacologic management of violence in psychiatric emergencies. *Primary Psychiatry, 16,* 49-53. |
| 34. Olsen (1998) | Wrong design | Olsen, D.P. (1998). Ethical considerations of video monitoring psychiatric patients in seclusion and restraint. *Archives of psychiatric nursing, 12,* 90-94 |
| 35. Patfield (2000) | Wrong design | Patfield, M. (2000). Creeping Custodialism. Australasian Psychiatry, 8, *370-372.* |
| 36. Quirk et al. (2004) | Not a surveillance tech | Quirk A, Lelliott P, Seale C. Service users' strategies for managing risk in the volatile environment of an acute psychiatric ward. Soc Sci Med. 2004 Dec;59(12):2573-83. doi: 10.1016/j.socscimed.2004.04.005. PMID: 15474210. |
| 37. Quirk et al. (2005) | Not a surveillance tech | Quirk, A., Lelliott, P. & Seale, C. (2005) Risk management by patients on psychiatric wards in London: An ethnographic study, Health, Risk & Society, 7:1, 85-91, DOI: 10.1080/13698570500034683 |
| 38. Rademeyer et al. (2009) | Wrong design | Rademeyer AJ, Blanckenberg MM, Scheffer C. Wireless physiological monitoring system for psychiatric patients. Annu Int Conf IEEE Eng Med Biol Soc. 2009;2009:5134-7. doi: 10.1109/IEMBS.2009.5334576. PMID: 19965038. |
| 39. Salzmann-Erikson et al. (2012) | Not a surveillance tech | Salzmann-Erikson M, Eriksson H. Panoptic power and mental health nursing-space and surveillance in relation to staff, patients, and neutral places. Issues Ment Health Nurs. 2012 Aug;33(8):500-4. doi: 10.3109/01612840.2012.682326. PMID: 22849776. |
| 40. Salmann-Erikson et al. (2011) | Not a surveillance tech | Martin Salzmann-Erikson , Kim Lützén , Ann-Britt Ivarsson & Henrik Eriksson (2011) Achieving Equilibrium within a Culture of Stability—Cultural Knowing in Nursing Care on Psychiatric Intensive Care Units, Issues in Mental Health Nursing, 32:4, 255-265, DOI: 10.3109/01612840.2010.549603 |
| 41. Shah et al. (1993) | Wrong design | Shah, A.K. & Ganesvaran, T. (1997) Inpatient Suicides in an Australian Mental Hospital. *Australian and New Zealand Journal of Psychiatry* 31:2, pages 291-298. |
| 42. Simpson (2023) | Wrong design | Simpson, A. (2023) Surveillance, CCTV and body-worn cameras in mental health care, Journal of Mental Health, 32:2, 369-372, DOI: 10.1080/09638237.2023.2194988 |
| 43. Slemon et al. (2017) | Wrong design | Slemon A, Jenkins E, Bungay V. Safety in psychiatric inpatient care: The impact of risk management culture on mental health nursing practice. *Nurs Inq*. 2017, 00:e12199. https://doi.org/10.1111/nin.12199 |
| 44. Stanley et al. (2020) | Wrong design | Stanley B, Mann JJ. The Need for Innovation in Health Care Systems to Improve Suicide Prevention. JAMA Psychiatry. 2020 Jan 1;77(1):96-98. doi: 10.1001/jamapsychiatry.2019.2769. PMID: 31577340. |

| | | |
|---|---|---|
| 45. Steinauer et al. (2017) | No usable data | Steinauer R, Huber CG, Petitjean S, Wiesbeck GA, Dürsteler KM, Lang UE, Seifert C, Andreeff K, Krausz M, Walter M, Vogel M. Effect of Door-Locking Policy on Inpatient Treatment of Substance Use and Dual Disorders. Eur Addict Res. 2017;23(2):87-96. doi: 10.1159/000458757. Epub 2017 Mar 29. PMID: 28351023. |
| 46. Stolovy et al. (2015) | Wrong design | Stolovy T, Melamed Y, Afek A. Video Surveillance in Mental Health Facilities: Is it Ethical? Isr Med Assoc J. 2015 May;17(5):274-6. PMID: 26137651. |
| 47. Tao et al. (2021) | Wrong setting | Tao X, Shaik TB, Higgins N, Gururajan R, Zhou X. Remote Patient Monitoring Using Radio Frequency Identification (RFID) Technology and Machine Learning for Early Detection of Suicidal Behaviour in Mental Health Facilities. Sensors (Basel). 2021 Jan 24;21(3):776. doi: 10.3390/s21030776. PMID: 33498893; PMCID: PMC7865785. |
| 48. Temkin et al. (2004) | Not a surveillance tech | Temkin TM, Crotty M. Suicide and Other Risk Monitoring in Inpatient Psychiatry. Journal of the American Psychiatric Nurses Association. 2004;10(2):73-80. doi:10.1177/1078390304263042 |
| 49. Theile & Snyder (2009) | No access | Theile D, Snyder B. The role of security in the pediatric psychiatric environment. J Healthc Prot Manage. 2009;25(2):58-65. PMID: 19711792. |
| 50. Tully et al. (2016) | Wrong design | Tully, J., Fahy, T., & Larkin, F. (2016). New technologies in the management of risk and violence in forensic settings. CNS Spectrums, Available on CJO 2015 doi:10.1017/S1092852915000279 |
| 51. Voigtlaender et al. (2016) | Wrong design | Voigtlaender, B.S., Scholzel, D., Schonherr, A., Schlobhayer, J. & Barth, T. (2016). Recognizing high-risk behavioural patterns in emergency psychiatry: From surveillance to technical assistance, insights into an innovative project* from the point of view of potential users. *European Psychiatry, 33,* S172. |
| 52. Voskes et al. (2014) | No access | Voskes Y, Kemper M, Landeweer EG, Widdershoven GA. Preventing seclusion in psychiatry: a care ethics perspective on the first five minutes at admission. Nurs Ethics. 2014 Nov;21(7):766-73. doi: 10.1177/0969733013493217. Epub 2013 Sep 12. PMID: 24036666. |
| 53. Tilt et al. (2003) | Wrong design | Tilt R. High-security hospitals. *British Journal of Psychiatry*. 2003;182(6):548-548. doi:10.1192/bjp.182.6.548 |
| 54. No authors listed | No access | Dealing with mental hospital violence and abuse. Hosp Secur Saf Manage. 1999 Jul;20(3):5-8. PMID: 10539575. |
| 55. Unknown | No access | Ganz e Rolfes prigione psichiatrica, Berlino |
| 56. Unknown | No access | Further measures for increased safety in high security hospitals |
| 57. Burns (1998) | Wrong design | Burns, T . (1998). Not just bricks and mortar. Report of the Royal College of Psychiatrists working party on the size, staffing, structure, siting, and security of new acute adult psychiatric in-patient units. *Psychiatric Bulletin, 22,* 465-466. |

| | | |
|---|---|---|
| 58. Commission on Acute Adult Psychiatric Care (2016) | Wrong design | Commission on Acute Adult Psychiatric Care (2016). The Commission to Review the Provision of Acute Inpatient Psychiatric Care for Adults in England, Wales and Northern Ireland. Background Briefing Paper. URL: www .evidence.nhs.uk/Search?om= %5B%7B%22toi %22:%5B%22Evidence %20Summaries%22%5D%7D %5D&q=acute+inpatient+mental+health&s =Relevance |
| 59. Department of Health and Social Care (2018) | Wrong design | Department of Health and Social Care (2018). The High Security Psychiatric Services (Arrangements for Safety and Security) Directions and associated guidance. Available at: chrome-extension://efaidnbmnnnibpcajpcglclefindmkaj/https://assets.publishing.service.gov.uk/media/5b0fca1bed915d2cd5d01a9c/consultation-on-the-high-security-psychiatric-services-directions.pdf [Accessed 18/03/24]. |
| 60. Gooding (2019) | Wrong design | Gooding, P. (2019). Mapping the rise of digital mental health technologies: Emerging issues for law and society. *International journal of law and psychiatry*, *67*, 101498. |
| 61. Greer (2021) | Wrong design | Greer, B. (2021). *Using Remote Monitoring to Improve Prediction and Develop Understanding of Aggression in Inpatient Mental Health Services* (Doctoral dissertation, King's College London). |
| 62. Johnson et al. (2023) | Wrong design | Johnson, E. A., Dudding, K. M., & Carrington, J. M. (2023). When to err is inhuman: An examination of the influence of artificial intelligence-driven nursing care on patient safety. Nursing Inquiry, e12583. |
| 63. Gooding & Clifford (2021) | Wrong design | Gooding, P. M., & Clifford, D. M. (2021). Semi-automated care: Video-algorithmic patient monitoring and surveillance in care settings. Journal of bioethical inquiry, 18(4), 541-546. |
| 64. Appenzeller et al. (2020) | Wrong design | Appenzeller, Y.E., Appelbaum, P.S. & Trachsel, M. (2020) Ethical and practical issues in video surveillance of psychiatric units. Psychiatric Services, 71(5), 480–486. https://doi.org/10.1176/appi.ps.201900397 |
| 65. Cowman et al. (2017) | Not a surveillance tech/ wrong design | Cowman, S., Björkdahl, A., Clarke, E., Gethin, G., Maguire, J. & European Violence in Psychiatry Research Group (EViPRG). (2017) A descriptive survey study of violence management and priorities among psychiatric staff in mental health services, across seventeen european countries. BMC Health Services Research, 17(1), 59. Available from https://doi.org/10.1186/s12913-017-1988-7 |
| 66. Cowman & Bowers (2009) | Not a surveillance tech/ wrong design | Cowman, S. & Bowers, L. (2009) Safety and security in acute admis-sion psychiatric wards in Ireland and London: a comparative st udy. Journal of Clinical Nursing, 18(9), 1346–1353. Available from: https://doi.org/10.1111/j.1365-2702.2008.02601.x |
| 67. Stephens (2007) | Wrong design | Stephens, M. 2007. "Risk, Security and Surveillance: The Care and Control of People with Serious Mental Health Problems." Security Journal 20: 211-221 |

| | | |
|---|---|---|
| 68. Irish & Shubert (2015) | Wrong design | Irish, A. and Shubert, T. (2015) Body worn camera use in health care facilities. IAHSS-F RS-15-01. Evidence Based Healthcare Security Research Series. chrome-extension://efaidnbmnnnibpcajpcglclefindmkaj/https://iahssf.org/assets/IAHSS-Foundation-Body-Worn-Cameras.pdf |
| 69. Tully et al. (2014) | Wrong design | Tully, J., Hearn, D., & Fahy, T. (2014). Can electronic monitoring (GPS 'tracking') enhance risk management in psychiatry? The British Journal of Psychiatry, 205, 83–85. |
| 70. Lloyd-Jukes et al. (2021) | Wrong design | Lloyd-Jukes, H., Gibson, O. J., Wrench, T., Odunlade, A., & Tarassenko, L. (2021). Vision-based patient monitoring and management in mental health settings. Journal of Clinical Engineering, 46(1), 36–43. doi: 10.1097/JCE.0000000000000447. |
| 71. Ayers et al. (1988) | No surveillance tech | Ayers, J., Maloney, C., Nimlos, K. (1988). Videotape feedback in an inpatient psychiatric service. Hospital and Community Psychiatry, 39, 777-780. |
| 72. Crowner et al. (1991) | No access | Crowner, M. L., Douyon, R., Convit, A., & Volavka, J. (1991). Videotape recording of assaults on a state hospital inpatient ward. Journal of Neuropsychiatry, 3(suppl), 9-14. |
| 73. Barnard & Neill (2006) | Wrong design | Barnard, M. & Neill, P. (2006). Report of the external review of John Meyer War Following the death of Eshan Charrun. NHS London. Available at: chrome-extension://efaidnbmnnnibpcajpcglclefindmkaj/https://hundredfamilies.org/wp/wp-content/uploads/2013/12/JASON_CANN_ward_review_LON.pdf [Accessed on 18/03/24] |
| 74. Bergmann & McGregor (2011) | Wrong design | Bergmann, J., and McGregor, A. (2011). Body-worn sensor design: What do patients and clinicians want? Ann. Biomed. Eng. 39(9):2299–2312. |
| 75. Bureau of Justice Assistance (2015) | Wrong setting | Bureau of Justice Assistance. (2015). Body-Worn CameraToolkit. [Cited 01 06 2019]. Available from: URL https://www.bja.gov/bwc/ |
| 76. Desai (2009) | Wrong design | Desai, S. (2009). The new stars of CCTV: What is thepurpose of monitoring patients in communal areas ofpsychiatric hospital wards, bedrooms and seclusion rooms? Diversity & Equality in Health and Care,6(1), 12 |
| 77. Malek et al. (2023) | Wrong design | Malek S, Hearn D, Fahy T, Tully J, Exworthy T. Legal and human rights issues in the use of electronic monitoring (using GPS 'tracking' technology) in forensic mental health settings in the UK. Medicine, Science and the Law. 2023;63(4):309-315. doi:10.1177/00258024231174820 |

| | | |
|---|---|---|
| 78. Laurie et al. (2021) | No usable data | J. Laurie, N. Higgins, T. Peynot, L. Fawcett, and J. Roberts, 'An evaluation of a video magnification-based system for respiratory rate monitoring in an acute mental health setting', *International Journal of Medical Informatics*, 2021, doi: 10.1016/j.ijmedinf.2021.104378. |
| 79. McDonald (2021) | Wrong design | McDonald, K. (2021). AI-enabled CCYV in aged care trial gets underway. PULSE+IT Available at: https://www.pulseit.news/aged-community-disabled-care/ai-enabled-cctv-in-aged-care-trial-gets-underway/ [Accessed on 18/03/24] |
| 80. West (2021) | Wrong design | West, K. (2021). Oxehealth. NHS Health Innovation Network. Available at: https://nhsaccelerator.com/innovation/oxehealth/ [Accessed on 18/03/24]. |
| 81. Gooding & Kariotis (2021) | Wrong design | P. Gooding and T. Kariotis, 'Ethics and Law in Research on Algorithmic and Data-Driven Technology in Mental Health Care: Scoping Review', *JMIR Mental Health*, vol. 8, no. 6, p. e24668, Jun. 2021, doi: 10.2196/24668. |
| 82. De Looff et al. (2019) | Not surveillance tech | de Looff P, Noordzij ML, Moerbeek M, Nijman H, Didden R, Embregts P. Changes in heart rate and skin conductance in the 30 min preceding aggressive behavior. Psychophysiology. 2019 Oct;56(10):e13420. doi: 10.1111/psyp.13420. Epub 2019 Jun 11. PMID: 31184379. |
| 83. Rahman (2019) | Wrong design | Rahman, T. (2019). Should Trackable Pill Technology Be Used to Facilitate Adherence Among Patients Without Insight. *AMA Journal of Ethics, 21 (4),* E332-336 |
| 84. Wright & Swaran (2022) | Wrong setting | Wright, Kay BSc, BPS, PhD; Singh, Swaran FRCPsych, DM. Reducing Falls in Dementia Inpatients Using Vision-Based Technology. Journal of Patient Safety 18(3):p 177-181, April 2022. \| DOI: 10.1097/PTS.0000000000000882 |
| 85. Michard & Kalkman (2021). | Wrong setting | Michard, F. & Kalkman, C.J. (2021). Rethinking Patient Surveillance on Hospital Wards. *Anesthesiology* 135:531–540 doi: https://doi.org/10.1097/ALN.0000000000003843 |
| 86. Lewis et al. (2021) | Wrong design | Lewis, A., Gandhi, V., Venugopal, B., Bowen, J., Polkey, M. & Gibson, O. (2021). Non contact patient monitoring and management system (Oxevision) in patient management. *Physiotherapy, O15, 114 (supp 1 E13).* |
| 87. Abbe & O'Keeffe (2021) | Wrong setting | Abbe JR, O'Keeffe C. Continuous Video Monitoring: Implementation Strategies for Safe Patient Care and Identified Best Practices. J Nurs Care Qual. 2021 Apr-Jun 01;36(2):137-142. doi: 10.1097/NCQ.0000000000000502. PMID: 32658001; PMCID: PMC7899219. |
| 88. De Silva (2015) | Wrong design | De Silva, P. (2015). Transforming mental health wards utilising new technologies? *Progress in Neurology and Psychiatry, 19(60:* 6-8. |
| 89. Royal College of Nursing (2018) | Wrong design | Royal College of Nursing (2018). Body Worn Cameras: would they make it safer for nurses and patients? Available at: https://www.rcn.org.uk/magazines/Bulletin/2018/May/Body-cameras [Accessed 18/03/24]. |

| | | |
|---|---|---|
| 90. Victorian Government (2018) | Wrong design | Victorian Government (2018). 'Chief Psychiatrist's guideline: Surveillance and privacy in designated mental health services'. Available: https://www.health.vic.gov.au/sites/default/files/migrated/files/collections/policies-and-guidelines/s/surveillance-and-privacy-in-designated-mental-health-services-pdf.pdf [Accessed 18/03/24]. |
| 91. Wilson et al. (2021) | Wrong design | Wilson, K., Eaton, J., Foye, U., Ellis, M., Thomas, E. and Simpson, A. (2022), What evidence supports the use of Body Worn Cameras in mental health inpatient wards? A systematic review and narrative synthesis of the effects of Body Worn Cameras in public sector services. Int J Mental Health Nurs, 31: 260-277. https://doi.org/10.1111/inm.12954 |
| 92. Lum et al. (2019) | Wrong design | Lum C, Stoltz M, Koper CS, Scherer JA. Research on body-worn cameras: What we know, what we need to know. *Criminology & Public Policy*. 2019; 18: 93–118. https://doi.org/10.1111/1745-9133.12412 |

**Appendix D:** Detailed study characteristics table

| Author, year and country | Aim of study | Surveillance description | Study design | Inpatient setting | Sample detail (including control group) | Lived experience involvement | MMAT quality rating | Conflicts of interest |
|---|---|---|---|---|---|---|---|---|
| Barrera et al. 2020<br><br>Country:<br>England | Establish whether it is safe to conduct nursing observations remotely from the nursing office using VBPMM. | VBPMM; Oxevision by Oxehealth | Service improvement study/ feasibility study | An adult acute male inpatient mental health ward. | Patients, staff and relatives<br><br>Patients n = not reported<br>Staff n = 18<br>Relatives n = 10<br><br>Control:<br>During the initial implementation phase, sensor-assisted observations were compared to the existing observation protocol ('treatment as usual'). | Meetings with former patients, patients' relatives and front-line nursing staff, where they were asked (e.g., about patients' safety, data confidentiality and impact on staffing levels). Their feedback was used to modify the project. | Low | No |
| Bowers et al. 2002<br><br>Country:<br>England | Describe current safety and security measures used on acute psychiatric wards in London, and to explore the relationships between them. | CCTV/video surveillance; CCTV for security (location on ward not specified); brand(s) not specified | Quantitative survey | An adult acute male inpatient mental health ward. | N = 87 hospital wards<br><br>Mean number of beds per ward: 20.6<br><br>84% of wards were mixed:<br>10% were female only, 6% male only.<br><br>87% wards were within the NHS. | None reported | Low | None declared |
| Clark et al. 2021 | Primary aim: improve the quality of physical health monitoring by making accurate vital sign measurements more frequently available.<br><br>Secondary aims:<br>explore the clinical | VBPMM; Oxevision by Oxehealth | Proof of concept quality improvement project | A women's PICU in a hospital in South London. Age of the inpatient population not specified. | Staff, patients and carers<br><br>Patient in pre-implementation focus group n = 12<br><br>Patients surveyed post-surveillance in seclusion n = 12<br><br>Carers surveyed post surveillance in seclusion n = 6 | Focus groups were held with patients (n = 12) and staff, and a questionnaire was given to staff, before implementing VBPMM to explore their views. Adaptations were made to how it was | Low | Yes<br><br>Funding: This project was a jointly funded and supported collaboration between South London and Maudsley NHS Foundation Trust and Oxehealth Limited. |

| | | | | | | | | |
|---|---|---|---|---|---|---|---|---|
| | experience of integrating a technological innovation with routine clinical care. | | | | Staff n = not reported | implemented based on this feedback. | | Declaration of interest: DB is Head of Insights & Benefits Realisation at Oxehealth Ltd. OG is Director for Research & Development at Oxehealth Ltd. CW is Director for Mental Health at Oxehealth Ltd. |
| Curtis et al. 2013<br><br>Country: England | Evaluate a purpose built NHS inpatient mental health care facility, the 'New Hospital'. | CCTV/video surveillance; CCTV cameras in common areas; brand(s) not specified | Qualitative evaluation | The 'New Hospital' had 318 inpatient beds to care for patients with acute psychiatric illnesses, geriatric conditions, learning difficulties, and a significant number of forensic cases. Age of the inpatient population not specified. | Staff, patients, family and carers<br><br>Results are reported from 19 group or individual meetings, representing a subset from a total of 40 conversations in the wider study. It is unclear why this subset was selected.<br><br>Number of participants = not reported | None reported | High | None declared |
| Dewa et al. 2023<br><br>Country: UK | Conduct a qualitative service evaluation to explore both staff and patient perspectives on the use of Oxehealth technology in a high-secure forensic psychiatric hospital. | VBPMM; Oxevision by Oxehealth | Qualitative study | Broadmoor Hospital in South England within West London NHS Trust – an adult high-secure forensic inpatient service. | Staff and patients<br><br>Patients n = 12<br>Staff n = 12<br>Total n = 24<br><br>Gender:<br>Male patients: 12 (100%)<br>Male staff: 6 (50%) | Two patients (from a group of clinically stable patients on the ward approached by the research team) joined the research team as patient advisors. One patient advisor left part-way | High | None declared |

| | | | | | | |
|---|---|---|---|---|---|---|
| | | | | | Female patients: 0<br>Female staff: 3 (25%)<br>Prefer not to say patients: 0<br>Prefer not to say staff: 3 (25%)<br><br><u>Age:</u><br>Patients had a mean age of 37.7 (SD 10.9), and staff were slightly younger (34.2 ± 12.0)<br><br><u>Patients' primary diagnosis:</u><br>Paranoid schizophrenia: 5 (42%)<br>Bipolar affective disorder: 2 (17%)<br>Emotionally unstable personality disorder: 2 (17%)<br>Schizotypal disorder: 1 (1%)<br>Paranoid personality disorder: 1 (1%)<br>Mental and behavioural disorders due to multiple drug use and substance misuse: 1 (1%)<br><br><u>Ethnicity</u><br>*White-British*<br>Patients: 4 (33%)<br>Staff: 4 (33%)<br>*Asian-British*<br>Patients: 0<br>Staff: 2 (17%)<br>*Black or Black British – Caribbean*<br>Patients: 2 (1%)<br>Staff: 0<br>*Black or Black British – African*<br>Patients: 2 (17%)<br>Staff: 1 (1%) | through the project due to project delays and was replaced by another patient.<br><br>The patient advisors reviewed and edited the consent form through an iterative approach with researchers. The interview topic guides were by the researchers and patient advisors. One of the patient advisors co-created the initial coding framework and initial themes with researchers. The other patient advisor provided feedback on this in a 1-1 meeting with a researcher and this informed the next analysis stage. Both patient advisors were paid in accordance with hospital pay guidance (£5 per interview).<br><br>Researchers met regularly with the project advisors throughout the study, |

| | | | | | | |
|---|---|---|---|---|---|---|
| | | | | | *Black or Black British – British*<br>Patients: 1 (1%)<br>Staff: 0<br>*Black or Black British – Other/Unspecified*<br>Patients: 2 (2%)<br>Staff: 0<br>*White – Irish*<br>Patients: 1 (1%)<br>Staff: 1 (1%)<br>*Prefer not to say*<br>Patients: 0<br>Staff: 3 (25%)<br><br>Patients' average length of stay (days): 1052.8 (701.2)<br>Staff average length of service at Broadmoor Hospital (years): 9.3 (11.1)<br>Staff average length of career in mental health (years): 11.8 (11.6)<br><br>Ward level dependency:<br>*Assertive Rehabilitation*<br>Patients: 10 (83%)<br>Staff: 1 (1%)<br>*Increased support and assertive treatment*<br>Patients: 2 (17%)<br>Staff: 2 (17%)<br>*Admissions*<br>Patients: 0<br>Staff: 5 (42%)<br>*Not available*<br>Patients: 0<br>Staff: 3 (25%) | holding meetings face-to-face where possible. |

| | | | | | | | | |
|---|---|---|---|---|---|---|---|---|
| | | | | | Staff previous use of Oxehealth:<br>Yes: 2 (17%)<br>No: 10 (83%) | | | |
| Due et al. 2012<br><br>Country:<br>Australia | Explore the potential relationship between surveillance techniques, the enactment of security measures, and patient violence in mental health wards. | CCTV/video surveillance; CCTV or surveillance cameras in all areas on each ward except bedrooms and bathrooms; brand(s) not specified | Ethnographic case study | The mental health unit of a large public hospital in South Australia. The buildings comprised both a secure or 'locked' ward, and an open ward. Age of the inpatient population not specified. | Patients, staff and visitors. Further detail not reported. | None reported | High | None declared |
| Ellis et al. 2019 | Conduct a pilot project to evaluate whether issuing BWCs to mental health ward nurses was associated with a reduction in violence and aggression in recorded incident interventions. | BWCs; brand was Reveal trading as Calla | A quasi-experimental repeated measures design | Seven West London Trust mental health adult wards, including: two wards for local services admissions (male and female), a PICU (male), a low secure forensic ward (male), medium secure ward (female) and two enhanced medium secure wards (both female). | Staff and patients<br><br>Staff<br>N=63 filled out the pre-pilot questionnaire<br><br>Roles:<br>Security nurses, nurses in charge, response nurses<br><br>In this study there were 4 female wards and 3 male wards<br><br>Total number of beds across all wards 94 | None reported | Low | Yes<br><br>Funding:<br>The lead author's expenses incurred in attending meetings were reimbursed by the camera company, but the evaluation was carried out, analysed and written up independently. |

| | | | | | | | | |
|---|---|---|---|---|---|---|---|---|
| Greer et al. 2019 | Explore the attitudes of staff toward passive remote monitoring technology for risk of aggression in inpatient forensic mental health services, with a focus on the potential benefits that this technology could provide and barriers to implementation. | Wearable sensors; brands were E4 (Empatica Srl) and Everion (Biovotion Ltd) | Qualitative study using focus groups | Medium-secure forensic mental health service in South London, UK. Age of the inpatient population not specified. | Staff (n = 25)<br><br><u>Job roles</u><br>Staff nurse: 20/25<br>Student nurse: 3/25<br>Ward manager: 2/25<br><br><u>Age:</u><br>Mean age of staff was 42.7 years (SD = 11.6), range: 22-64<br><br><u>Sex:</u><br>Female: 16/25<br>Male: 9/25<br><br><u>Ethnicity:</u><br>Black African: 16/25<br>Black Caribbean: 5/25<br>White British: 4/25<br><br><u>Highest educational attainment:</u><br>Higher-level qualification (e.g. university degree, professional qualification): 22/25<br>Secondary (A-level equivalent): 3/25<br><br><u>Time in post</u><br>Average: 6 years 6 months (SD = 3 years 5 months) | The focus group topic guide was informed by consultation with two service user–caregiver advisory groups. | High | None declared |
| Hakimzada et al. 2020<br><br><u>Country:</u> UK | Explore the attitudes of psychiatric nursing staff towards the use of BWCs on psychiatric inpatient wards. | BWCs; brand(s) not specified | Quantitative and qualitative survey questionnaire | Seven inpatient wards in one Mental Health Trust in South West London, including a PICU, | Nursing staff (n = 60)<br><br><u>Gender:</u><br>50% female<br>41.7% male<br>8.3% did not report | None reported | Medium | None declared |

| | | | | | | | | |
|---|---|---|---|---|---|---|---|---|
| | | | | two acute wards and four secure wards. Age of the inpatient population not specified. | Age:<br>21.7% 20-30 years, 21.7% 31-40 years, 18.3% 41-50 years, 6.7% 51+, 31.7% did not report<br><br>Job contract:<br>Full-time: 70%<br>Part-time: 3.3%<br>Agency: 1.7%<br>No answer: 25%<br><br>Grade:<br>Band 2: 1.7%<br>Band 3: 25%<br>Band 4: 5%<br>Band 5: 30%<br>Band 6: 13.3%<br>Band 7: 3.3%<br>No answer: 21.7% | | | |
| Hardy et al. 2017<br><br>Country: England | Overall aim: examine the feasibility of using BWCs in an inpatient mental health setting.<br><br>Specific objectives:<br>• Find out whether wearing the camera is comfortable and if it causes restriction<br>• Test and refine information technology support and security requirements | BWCs; brand was Reveal trading as Calla | Mixed methods pre-post study | Berrywood Hospital, an adult psychiatric facility in Northampton, England, run by Northamptonshire Healthcare NHS Foundation Trust. The five wards in the pilot included one male and one female recovery, one low secure unit, one acute. | Staff and patients<br><br>Including all nursing staff on the wards where BWCs were being used, and staff in the response team using BWCs.<br><br>Reported number of staff and patient respondents to each survey question, but not overall numbers of participants. | None reported | Low | None declared<br><br>However, in the ‘acknowledgements’ section they stated: "We would like to thank Reveal trading as Calla (www.calla.co) for providing the cameras and the training." |

| | | | | | | | | |
|---|---|---|---|---|---|---|---|---|
| | • Determine the level of training and support required by staff using the BWCs<br>• Explore the experience of staff using BWCs, practical issues faced, their perceptions of its usefulness<br>• Explore the experience of staff who work alongside colleagues who are wearing BWCs, practical issues faced, their perceptions of its usefulness<br>• The acceptability of staff wearing BWCs to patients<br>• Observe any change in the level of reported incidents<br>• Examine the costs of utilising BWCs | | | | | | | |
| Krieger et al. 2018 | Assess patients' preferences regarding prevalent specific forms of coercive interventions, their accompanying emotions, and their understanding of the experience as measured at different | CCTV/video surveillance; the part of the questionnaire specifically asking patients about their preferences | Naturalistic trial | Three PICUs at the Asklepios Clinic North in Hamburg, Germany. Age of the inpatient population not specified. However, can be inferred that | Patients<br><br>Patients in coercive intervention group n = 213<br><br>Patients in control group (voluntary admission with no coercive treatment) n = 51<br><br><u>Sex (female/male (%))</u> | None reported | Medium | None declared |

| | | | | | | | | |
|---|---|---|---|---|---|---|---|---|
| | sites and different points in time using both interviews and self-assessments. | specified video surveillance in seclusion; brand(s) not specified | | patient participants included adults and children. | Coercive intervention group: 46.2/53.8<br>Control group: 43.1/56.9<br><br><u>Age:</u><br>Coercive intervention group, M(SD): 42.57 (13.05)<br>Control group, M(SD): 42.45 (12.42)<br><br><u>Nationality (native German/other):</u><br>Coercive intervention group: 78.4%/21.6%<br>Control group: 72.5%/27.5%<br><br><u>Years of formal education:</u><br>Coercive intervention group, M(SD): 10.52 (1.74)<br>Control group, M(SD): 10.54 (1.64) | | | |
| Malcolm et al. 2022 | The objective of this early economic evaluation was to explore the impact of introducing VBPMM (vision-based patient monitoring and management) with standard care, versus standard care alone on health and economic outcomes in PICUs across England | VBPMM; Oxevision by Oxehealth | Economic analysis study utilising a cost-calculator approach (using data from a single centre observational before and after study) | An adult PICU | Patients (n = not reported)<br><br><u>Age:</u><br>Mean: 36 years<br><br><u>Diagnosis:</u><br>35% schizophrenia, 24% mood affective disorders, 15% substance misuse. Other conditions included personality disorders, as well as neurosis and stress related disorders. | None reported | Low | Yes<br><br>Funding:<br>RM, JS and AS are employed by York Health Economics Consortium (YHEC). YHEC was funded by Oxehealth to develop the economic model and manuscript.<br><br>Author contributions: 'FN and KW were the chief and principal investigators respectively in the |

| | | | | | | | | |
|---|---|---|---|---|---|---|---|---|
| | | | | | | | | clinical study of Oxevision.' |
| Murphy et al. 2017 | To compare the costs of using GPS electronic monitoring (EM) in forensic psychiatric patients on leave from a medium-secure service by comparing the average total cost per patient with EM against the average total cost per patient without EM. | GPS electronic monitoring; brand(s) not specified | Retrospective observational study | River House, an adult medium-secure unit in South London and Maudsley NHS Foundation Trust (107 male beds and 15 female beds) | Patients<br><br>Intervention group n = 121<br>Control group n = 96<br>Total patients n = 175<br><br>Control group was patients who had used leave during a 3-month period in 2010 (no EM). Intervention group was patients who had used leave in the corresponding period in 2011 (during which EM had been implemented).<br><br><u>Sex:</u><br>Control group:<br>Female n = 13 (13.5%)<br>Male n = 78 (81.3%)<br>Unavailable: n = 5 (5.2%)<br><br>EM group:<br>Female n = 15 (12.4%)<br>Male n = 77 (63.6%)<br>Unavailable n = 29 (24%)<br><br><u>Age in EM group:</u><br>18-29yrs n=23 (19%);<br>30-39yrs n= 26 (21.5%);<br>40-49yrs n=31 (25.6%);<br>50-59yrs n=6 (5%);<br>60-69yrs n=5 (4.1%);<br>70-79yrs n=1 (0.8%)<br><br><u>Diagnosis in EM group:</u><br>Psychosis n=72 (59.5%); | None reported | Medium | None declared |

| | | | | | | | | |
|---|---|---|---|---|---|---|---|---|
| | | | | | Affective disorder n=8 (6.6.%);<br>Personality Disorder n=23 (19%);<br>Substance use Disorder n=9 (7.4%)<br><br><u>Other diagnoses in EM group</u>:<br>PTSD n=1 (0.8%); Borderline LD n=1; Mild LD n=1; Mental disorder NOS n=0;<br>No other diagnosis n=88 (72.2%) | | | |
| Ndebele et al. 2023<br><br><u>Country:</u><br>England | To examine the effect of adopting the contact-free vision-based patient monitoring and management (VBPMM) system into existing clinical practice on the number of incidents of self-harm in bedrooms (all types and ligatures specifically) on acute mental health inpatient wards. A minor aspect of the study was to include patient and staff feedback. | VBPMM; Oxevision by Oxehealth | Mixed methods non-randomized controlled before-and-after evaluation within a pilot study | At Caludon Centre, Coventry & Warwickshire Partnership NHS Trust (CWPT), a purpose-built facility, based on the University Hospital Coventry and Warwickshire (UHCW) site, providing inpatient and outpatient adult mental health care | Staff and patients<br><br>Number of patients in total = not reported<br><br><u>Intervention group:</u> two acute wards fitted with VBPMM (22-bed female and 20-bed male)<br><br><u>Control wards:</u> two acute wards without VBPMM selected based on the similarity of the patient cohort, ward size and clinical ways of working<br><br><u>Age of patients:</u><br>*Intervention group:*<br>Mean: 37 years<br>Range: 18-82<br>*Control group:*<br>Mean = 39 years<br>Range = 18-84<br><br><u>Diagnosis (intervention/ control):</u><br>Mood affective disorders 35%/32%;<br>Personality Disorder 28%/25%;<br>Disorders due to psychoactive substance use 14%/18%; | None reported | Low | Yes:<br><br>Paper stats that: 'The authors time was funded by their respective employers (CWPT for authors 1 and 2, Oxehealth Limited for authors 3 and 4). In accordance with Taylor & Francis policy and the ethical obligation as researchers, it is being disclosed that authors 3 and 4 have a financial interests in the supplier of the technology used in this study.' |

| | | | | | | | | |
|---|---|---|---|---|---|---|---|---|
| | | | | | Neurotic, stress-related and somatoform disorders 6%/8%;<br>Other 18%/18%. | | | |
| Nijman et al. 2011<br><br><u>Country:</u><br>UK | To investigate the prevalence of door locking and the use of other (additional) exit security measures on psychiatric admission wards in the UK, and to empirically study the associations between locking ward exit doors and absconding rates. | CCTV/video surveillance; brand(s) not specified | Cross sectional study | 133 adult acute psychiatric wards in London, Central England and Northern England which participated in the City-128 study (Bowers et al., 2007). | Staff answered surveys about the participating wards | None reported | High | None declared |
| Oxevision, 2022<br><br><u>Country:</u><br>England | Not clearly stated | VBPMM; Oxevision by Oxehealth | Mixed methods study | 13 wards, including the following services: female working age acute, male working age acute, mixed working age acute and psychiatric intensive care units (age not specified). | Patients (n = “over 75”)<br><br>Number of patients rating each statement ranged from 60-78. ‘No opinion’ responses were not included in these counts. Specific overall number of participants not stated. | None specifically described in relation to this report. However, the report describes generally how Oxehealth aims to involve patient and carers throughout the Oxevision implementation process and tries to gain feedback from them to improve care. States that patients’ and carers’ views are represented in Oxehealth through their partnerships with NHS Trusts, academic institutions and leading research | Low | Yes<br><br>This is an Oxehealth publication about Oxevision |

| | | | | | | | | |
|---|---|---|---|---|---|---|---|---|
| | | | | | | centres (including co-design, co-production and testing of new developments). | | |
| Peek-Asa et al. 2009 | Compare the workplace violence prevention programs in a sample of psychiatric units and facilities in New Jersey and California. The units and facilities were compared on four components: training, policies and procedures, environmental safeguards, and security. | CCTV/video surveillance; brand(s) not specified | Cross sectional survey | 83 psychiatric units within acute care hospitals and psychiatric facilities in New Jersey and California. Age of the inpatient populations not specified. | Psychiatric units were the individual unit of analysis<br><br>53 in California<br>30 in New Jersey<br><br><u>Unit types (California/New Jersey):</u><br>Psychiatric units in trauma hospitals: 12/7<br>Psychiatric unit in general acute care facility with more than 300 beds: 11/7<br>Psychiatric unit in general acute care with fewer than 300 beds: 12/11<br>Psychiatric facilities: 18/5 | None reported | Low | None declared |
| Shetty et al. 2023 | Explore the patients' experiences of different observation methods in seclusion and their influence on their connection and relations to staff, by patients in an Irish forensic mental health hospital, in order to inform future seclusion practices. | CCTV/video surveillance; video camera in seclusion room; brand(s) not specified | Retrospective phenomenological qualitative study | Medium secure wards (three male, one female) at an adult forensic mental health hospital in Ireland | Patients (n = 10)<br><br><u>Sex:</u><br>Male n = 8<br>Female n = 2<br><br>All had experienced seclusion in the last five years. | None reported | High | None declared |
| Simpson et al. 2011<br><br><u>Country:</u><br>England | Discover whether rates of drug/ alcohol use on acute psychiatric wards were related to levels | CCTV/video surveillance; CCTV brand(s) not npecified | Cross sectional study | 136 acute adult psychiatric wards across London, Central England and North England | Same as Nijman et al. 2011. | None reported | High | None declared |

| | | | | | | | | |
|---|---|---|---|---|---|---|---|---|
| | and intensity of exit security measures. | | | | | | | |
| Steinert et al. 2014<br><br><u>Country:</u><br>Germany | Conduct an online survey on the current practice of coercive measures in German psychiatric hospitals, in light of regional legal prohibition of video surveillance (Nordrhein-Westfalia) in 2011. | CCTV/video surveillance; video monitoring used during restraint; brand(s) not specified | Cross-sectional survey (online questionnaire) | 88 psychiatric hospitals in Germany<br><br>This includes 36 specialist hospitals, 41 departments within general hospitals and 13 university hospitals.<br><br>These included general psychiatry hospitals, as well as those for addictions, forensic psychiatry and old-age psychiatry.<br><br>Age of the inpatient populations not specified.<br><br>Hospitals had on average 151 beds (range 40-545); 20 hospitals also had forensic beds, on | Staff at medical director level responded to the online survey (n = 88) | An online questionnaire was developed together with the working group for the prevention of violence and coercion in psychiatry and the regional association of psychiatry-experienced people in Baden-Wuerttemberg. Sent first to working group (mostly professional) then to patient group. Changes were made in both rounds of consultation. | High | None declared |

| | | | | | | | | |
|---|---|---|---|---|---|---|---|---|
| | | | | average 101 (range 5-384). All but 3 had compulsory responsibility for a specific geographical area and took patients who were compulsorily admitted. | | | | |
| Tapp et al. 2016<br><br><u>Country:</u> Multi-country (including UK, Canada, Norway, China, Finland and The Netherlands) | Establish whether experts with clinical and/or research experience in this setting could reach consensus on elements of high-security hospital services that would be essential to the rehabilitation of forensic patients. | CCTV/video surveillance; CCTV brand(s) not npecified | Three-round Delphi study | Forensic high security inpatient mental health services. Age of the inpatient population not specified. | Staff (n = 54)<br><br><u>Roles (Round 1)</u><br>Nursing n=15 (27.8%);<br>Psychiatry n=15 (27.8%);<br>Psychology n=14 (25.9%); Occupational therapy n=4 (7.4%);<br>Social work n=2 (3.7%);<br>Security services n=1 (1.9%) Pharmacy n=2 (3.7%);<br>Speech and language therapy n=1 (1.9%).<br><br><u>Roles (Round 3):</u> Nursing n=8 (22.9%);<br>Psychiatry n=8 (22.9%);<br>Psychology n=11 (31.4%); Occupational therapy n=4 (11.4%);<br>Social work n=2 (5.7%);<br>Security services n=0;<br>Pharmacy n=2 (5.7%);<br>Speech and language therapy n=0. | None reported | Medium | None declared |
| Tron et al. 2018<br><u>Country:</u> Israel | i) Develop and evaluate a framework for using wearable devices to facilitate | Wearable sensor; smartwatch (GeneActiv) | Quantitative evaluation | Closed adult inpatient wards at Shaar-Meashe | Patients<br><br>Patients n = 25 (after 2 dropouts) | None reported | Low | None declared |

| | | | | | | | | |
|---|---|---|---|---|---|---|---|---|
| | continuous motor deficits monitoring in schizophrenia patients in a natural setting<br>ii) Help characterise subtypes of schizophrenia to better understand its causes and develop more personalised treatments. | worn by patients | | mental health centre. | Age:<br>Range: 21 to 58 (mean: 37.48)<br><br>Diagnosis (DSM-5 criteria):<br>Schizophrenia: 21/27<br>Paranoid schizophrenia: 3/27<br>Schizoaffective disorder: 2/27<br>Psychotic state cannabinoids: 1/27<br><br>Course of illness:<br>Ranged from 0 (first hospitalisation) to 37 years (mean: 16.9 years) | | | |
| Tully et al. 2016<br><br>Country: England | Determine whether the introduction of Electronic Monitoring (EM) using GPS 'tracking' led to a reduction in episodes of leave violation. They also aimed to assess the extent to which EM affected the amount of overall leave and the proportion of leave that was unescorted. They hypothesised that the introduction of EM would reduce leave violations, increase overall leave and increase the proportion of unescorted leave. | GPS electronic monitoring; brand was 'Buddi Tracker' | Observational pre-post study | The South London and Maudsley (SLaM) medium-secure service, which commenced in April 2010, comprising 120 beds. At the time of the study, these beds were located in two medium-secure units in South London. Age of the inpatient population not specified. | N/A | None reported | Low | None declared |

| | | | | | | | | |
|---|---|---|---|---|---|---|---|---|
| Vartianinen & Hakola, 1994<br><br><u>Country:</u> Finland | To study, with a questionnaire, the effects of TV monitoring on patients and personnel. | CCTV/video surveillance; CCTV brand(s) not npecified | Pre-post study using a survey | Four closed adult male wards in the Niuvanniemi hospital in Finland. | Staff and patients<br><br>Staff n = 97 (90% of all staff)<br><br>Patients n = 77<br>Patients' sex: 100% male<br><br><u>Response rates on wards 3 and 4 (where CCTV was introduced):</u><br>>90% return rate of the first questionnaire (pre-CCTV)<br>85% response rate for ward 3 and 75% response rate for ward 4 after CCTV was introduced<br><br><u>Response rates on wards 1 and 2 (no CCTV):</u><br>100% response rate for the first questionnaire<br>After the removal of rooms, return rates were 81% on ward 1 and 86% on ward 2 after bedrooms were removed<br><br>Approximately 70% of patients had committed violent crime<br><br><u>Patients' diagnosis:</u><br>Schizophrenia: approximately 90% of the patients in wards 2, 3, and 4, 70% in ward 1 | None reported | Low | None declared |
| Warr et al. 2005<br><br><u>Country:</u> England | Determine the acceptable use of CCTV surveillance in a mental health inpatient unit and | CCTV/video surveillance; CCTV in bedrooms; brand(s) not specified | Qualitative interview study | Montpellier adult low-secure unit in England | Staff and patients<br><br>Staff n = 10<br>All staff had experience of using CCTV for observation at night | None reported | Medium | None declared |

| | | | | | | | | |
|---|---|---|---|---|---|---|---|---|
| | whether it benefits patient care. | | | | Patients n = 6<br><br><u>Patients' age:</u><br>21-45 years<br><br>All patients had a primary diagnosis of schizophrenia with lengthy histories of previous admissions to hospital and penal institutions. Sample included those being observed by CCTV and those who were not. | | | |
| Wilson et al. 2023<br><br><u>Country:</u><br>England | Explore the perspectives of patients, mental health staff, and senior management to identify the possible impacts of body-worn cameras in inpatient mental health settings. | BWCs; brand(s) not specified | Explorative qualitative study | Five NHS acute adult inpatient wards across England.<br><br><u>Site A:</u> Four hospital sites in London with 17 acute wards; staff not using BWCs.<br><br><u>Site B:</u> Hospital in Southwest England with three acute wards; BWCs in use only by staff in the health based place of safety room.<br><br><u>Site C:</u> One hospital site in London with three | Staff and patients<br><br>Total n = 64<br>Staff n = 25<br>Patients n = 24<br>Service users from Twitter n = 9<br>Mental health nursing directors n = 6<br><br><u>Staff roles:</u><br>Clinical support worker, mental health nurse, occupational therapist, head of nursing and quality, physical health team, security/reception, clinical team leader, student nurse, healthcare assistant, violence prevention trainer, ward manager, assistant psychologist, mental health nurse director<br><br><u>Gender:</u><br>Staff overall:<br>Male = 19, female = 7, not reported = 1. | Yes. A lived experience consultant and a lived experience research assistant, both with personal experience of inpatient care, were key members of the research team. Both helped form and facilitate a Lived Experience Advisory Panel for this research, which consisted of eight people with diverse lived experience of inpatient mental healthcare including some with experience of BWCs. The involvement group developed a plain | High | None declared |

| | | | | | | | | |
|---|---|---|---|---|---|---|---|---|
| | | | | acute wards; staff using BWCs.<br><br><u>Site D:</u> Two hospital sites in Northeast England with six acute wards; staff using BWCs.<br><br><u>Site E:</u> One hospital site in London with three acute wards; staff using BWCs. | Mental health nurse directors: Male = 3, female = 3.<br><br>Patients:<br>Male = 6, female = 11, transgender = 1, not reported = 1. | language glossary for this paper, included in Appendix I. | | |
| Zakaria & Ramli, 2018<br><br><u>Country:</u><br>Malaysia | Identify patients' perception on physical privacy dimensions proposed by Carew and Stapleton | CCTV/video monitoring; brand(s) not specified | Qualitative study | Psychiatric wards at a teaching hospital in Malaysia (included child and adult inpatients) | Patients (n = 25)<br><br><u>Sex:</u><br>Male n = 14<br>Female n = 11<br><br><u>Age:</u><br>Range 14-54 years | None reported | High | None declared |

Acronyms: BWCs = Body Worn Cameras; CCTV = Closed Circuit Television; EM = Electronic Monitoring; GPS = Global Positioning System; MMAT = Mixed Methods Appraisal Tool; NHS = National Health Service; PICU = Psychiatric Intensive Care Unit; UK = United Kingdom; USA = United States of America; VBPMM = Vision-Based Patient Monitoring and Management.

**<u>Appendix E:</u>** Table summarising how surveillance technologies in inpatient mental health settings were implemented in the included studies

| **Surveillance technology and brand names** | **How the technology was implemented and intended functions** | **Settings** | **Informed consent procedures** | **Lived experience involvement in implementation** |
|---|---|---|---|---|
| Vision-Based Patient Monitoring and Management<br><br>(Barerra et al., 2020; Clarke et al., 2021; Dewa et al., 2023; Malcolm et al., 2022; Ndebele et al., 2023; Oxehealth, 2022)<br><br>(6 papers)<br><br><u>Brands :</u><br>All Oxevision | Oxevision is a wall-mounted system with an optical sensor (a Class IIa medical device) which enables contact-free monitoring of patients' pulse rate and breathing rate through photoplethysmography and chest wall/abdomen movements (Barerra et al., 2020; Oxehealth, 2022). An infrared-sensitive camera is used so that the technology can be used in any lighting conditions (Clark et al., 2021; Ndebele et al., 2023). These remote vital sign observations are made at regular intervals (e.g., every five minutes in Clark et al. (2021), but are only available when the patient is sufficiently still for them to be accurate (Clark et al., 2021). Video can be viewed for up to 15 seconds when taking vital sign measurements or responding to alerts. In the latter case, only deidentified blurred video is available (Ndebele et al., 2023). The device also monitors patients' movement and generates location and activity-based alerts to notify staff when patients may need assistance, for example, if they have spent a long time in their bathroom, if others have entered their bedroom, or if they have left their room at night (Ndebele et al., 2023; Oxehealth, 2022). It therefore allows staff to continuously monitor patients (Malcolm et al., 2022). Nurses can access the system via monitors at the nurses' station and portable tablets (Malcolm et al., 2022). If staff have concerns, they should check on the patient in-person (Barrera et al., 2020). The system is intended as an adjunct to usual care, not as a replacement for therapeutic interaction or personal care (Barerra et al., 2020; Oxehealth, 2022).<br><br><u>Stated aims/functions:</u><br>The Oxevision system aims to avoid the need for staff to disturb patients for checks (e.g., by entering their room and turning on | <u>Countries:</u><br>• All UK-based<br><br><u>Ward types :</u><br>• Acute wards (Barerra et al., 2020 ; Ndebele et al., 2023 ; Oxehealth, 2022).<br>• PICUs (Clark et al., 2021 ; Malcolm et al., 2022; Oxehealth, 2022).<br>• High-secure forensic service (Dewa et al., 2023).<br><br><u>Location within wards:</u><br>• In five studies, the Oxevision system was located in patients' bedrooms (Barerra et al., 2020; Dewa et al., 2023; Malcolm et al., 2022; Ndebele et al., 2023; Oxehealth, 2022).<br>• In one study it was located in a PICU seclusion room (Clark et al., 2021) | Methods of staff informing patients about the Oxevision system included: discussion (Clark et al., 2021; Malcolm et al., 2022; Ndebele et al., 2023; Oxehealth, 2022), information leaflets (Malcolm et al., 2022; Ndebele et al., 2023), and posters on the ward (Clark et al., 2021). Malcolm et al. (2022) also reported informing carers, where appropriate, about the system.<br><br>Ndebele et al. (2023) specified that consent was sought from patients to use Oxevision in their bedroom. Decisions were made by consultees (e.g., the patient's carer or the ward's Consultant Psychiatrist) in cases where patients lacked capacity. Consent was sought from the patient once they had capacity to make this decision. Where consent was not given, the system remained switched off in the patient's bedroom for the duration of their stay.<br><br>Barrera et al. (2020) and Dewa et al. (2023) did not report | In Clark et al.'s (2021) study, a patient focus group (n = 12) was held before implementing Oxevision to explore their views and address concerns prior to implementation.<br><br>Oxehealth's (2022) report emphasises early and continuous patient and carer involvement throughout the implementation process, and their own research, through collaborating with NHS Trusts and research institutions to ensure patient and carer feedback shapes the technology's development.<br><br>Barrera et al. (2020) stated: "there were extensive PPI activities, including meetings with former patients and patients' relatives. Similarly, meetings were held with front-line nursing staff, where the project was modified following suggestions. Patients as well as staff members asked challenging questions, including concerns about |

| | | | | |
|---|---|---|---|---|
| | the lights) (Barerra et al., 2020; Dewa et al., 2023). It also enables observations to be made without requiring patients' cooperation (Barerra et al., 2020). Other reported intended functions include: helping staff respond to patient needs more quickly and efficiently (Malcolm et al., 2022), facilitating monitoring and mitigation of self-harm risk (Ndebele et al., 2023), preventing incidents (Dewa et al., 2023), supporting care planning (Oxehealth, 2022), supporting compassionate and dignified care (Oxehealth, 2022) and reducing NHS mental health care costs (Malcolm et al., 2022). | | procedures for obtaining consent for the use of Oxevision. | patients' safety, data confidentiality and impact on staffing levels."<br><br>Lived experience involvement in implementation was not reported in the remaining Oxehealth studies (Dewa et al., 2023; Malcolm et al., 2022; Ndebele et al., 2023). |
| CCTV/video surveillance<br><br>(Bowers et al., 2002; Curtis et al., 2013; Due et al., 2012; Krieger et al., 2018; Nijman et al. 2011, Simpson et al., 2011; Steinert et al., 2014 Tapp et al., 2016; Vartiainen & Hakola, 1994; Peek-Asa et al., 2009 Zakaria & Ramli, 2018; Warr et al., 2005; Shetty et al., 2023)<br><br>(13 papers overall)<br><br><u>Brands:</u><br>None specified | In the included studies, CCTV was implemented in communal areas (Curtis et al., 2013; Due et al., 2012; Nijman et al., 2011; Vartiainen & Hakola, 1994), including corridors (Vartiainen & Hakola, 1994), and at exit doors (Nijman et al,. 2011; Simpson et al, 2011). It has also been implemented in seclusion rooms (Vartiainen & Hakola, 1994), and patients' bedrooms (Warr et al., 2005). Some studies specified that it was not used in patients' bedrooms (Curtis et al., 2013; Due et al., 2012) or bathrooms (Due et al,. 2012). Due et al. (2012) and Warr et al. (2005) described how screens located in each duty station enabled staff to monitor patients continuously. In Warr et al. (2005), CCTV in patients' bedrooms was only intended to be monitored during nights.<br><br>Warr et al. (2005) stated that the cameras were fitted with a sensitive audio facility capable of detecting breathing in most people whilst asleep. They also used infrared cameras to enable observation without any visible light. They specified that images and sound were not recorded.<br><br>Studies have shown no association between CCTV use and other security measures (Bowers et a., 2002; Nijman et al., 2011). Bowers et al. (2002) also found no significant differences in CCTV use on wards between private and NHS facilities, or by gender of occupants. | <u>Countries:</u><br>• UK (Bowers et al., 2002; Curtis et al., 2013; Nijman et al,. 2011)<br>• Australia (Due et al., 2012)<br>• Germany (Krieger et al., 2018; Steinert et al., 2014)<br>• Finland (Vartiainen & Hakola, 1994)<br>• USA (Peek-Asa et al., 2009)<br>• Malaysia (Zakaria & Ramli, 2018)<br>• International - (Delphi study) (Tapp et al., 2016)<br><br><u>Ward types:</u><br>• Acute wards (Curtis et al., 2013)<br>• PICUs (Krieger et al., 2018) | Not reported on in the included studies. | None reported in the included studies. |

| | | | | |
|---|---|---|---|---|
| | <u>Stated aims/functions:</u><br>The described functions of CCTV/video surveillance in inpatient settings include: continuously monitor both patients' behaviour [Curtis et al., 2013; Nijman et al., 2011; Steinert et al., 2014; Zakaria & Ramli, 2018) and staff's (Curtis et al., 2013) behaviour; making observation less disruptive (Warr et al., 2005), monitoring who is leaving the ward (Nijman et al., 2011), monitoring safety during mechanical restraint (Steinert et al., 2014), reducing institutional incidents (Tapp et al., 2016) and preventing violence (Peek-Asa et al., 2009). In Krieger et al. (2018) it was considered a coercive intervention. | • Forensic high-secure wards (Tapp et al,. 2016)<br>• Inpatient mental health facility with beds for acute psychiatric illnesses, geriatric conditions, learning difficulties and a significant number of forensic cases (Curtis et al., 2013).<br>• Unspecified (Bowers et al., 2002; Due et al., 2012; Peek-Asa et al., 2009; Steinert et al., 2014; Vartiainen & Hakola, 1994; Zakaria & Ramli, 2018)<br><br><u>Location within wards:</u><br>Communal areas (e.g., corridors, at exit doors), seclusion rooms, patients' bedrooms. Some studies specified not within private areas (e.g., bedrooms, bathrooms). | | |
| Body Worn Cameras (BWCs)<br><br>(Ellis et al., 2019; Hakimzada et al., 2020; Hardy et al., 2017; Wilson et al,. 2023) | Body worn cameras (BWCs) are recording devices used to document interactions between staff and patients on psychiatric inpatient wards. They can make audio and video recordings. BWCs are small, securely attached to staff uniforms (e.g., using a harness) and worn continuously by trained staff. BWCs are manually activated by staff. Some papers described BWCs only being activated when incidents occurred (Ellis et al., 2019; Hardy et al., 2017). Ellis et al. (2019) further specified that | <u>Countries:</u><br>All UK-based<br><br><u>Ward types:</u><br>• Acute wards (Ellis et al., 2019 ; Hardy et al,. 2017; Wilson et al., 2023) | In Hardy et al.'s (2017) study, all wards were provided with fair processing notices in the form of posters which were displayed in areas of high visibility. These stated that: the cameras record video and audio information, but only when activated by the | None reported in any of the studies. |

| | | | |
|---|---|---|---|
| (4 papers)<br><br><u>Brands:</u><br>• Calla (Ellis et al., 2019; Hardy et al., 2017)<br>• Unspecified (Hakimzada et al,. 2020; Wilson et al,. 2023) | BWCs use was only for incidents/interventions "that would normally be the subject of an IR1 or witness statement". However, other papers were less specific in describing activation timing.<br><br>BWC activation is signalled, for example, by a light and audible beep (Hardy et al., 2017). Staff should also inform patients before recording (Hakimzada et al,. 2020). In Hardy et al.'s (2017) study, staff were advised to inform patients and other staff that the wearing of the camera was for safety. They were also trained to talk to the camera to share what they can see and what they intend to do. If they decided to turn the camera off because it was exacerbating the situation, they were advised to inform patients and tell them why. Staff could turn the camera around if it was appropriate to record sound, but not visuals (Hardy et al., 2017).<br><br><u>Data storage, transfer and access</u><br>Access to camera footage is protected by a PIN so data cannot be downloaded if the camera is lost (Hardy et al., 2017). Cameras may display the date, time and storage time. In Hardy et al.'s (2017) study, cameras were docked, recharged and data was uploaded to a secure cloud via a computer in the reception area. Footage was kept for 31 days unless there was an incident that needed to be reviewed. Only ward matrons were able to access footage, and from their ward only. In Ellis et al.'s (2019) study, footage was stored on a secure cloud for 30 days and then automatically deleted unless secured for a specific purpose, including internal investigation, staff reflection and training exercises, and/or evidence related to a criminal investigation.<br><br>BWCs may only be worn by certain members of staff during shifts, not all (e.g., in Hardy et al. (2017) they were worn by a member of nursing staff on each ward, a member of the | • Low-secure forensic units (Ellis et al., 2019; Hardy et al., 2017)<br>• Medium and medium enhanced secure forensic wards (Ellis et al,. 2019)<br>• Male and female recovery wards (Hardy et al., 2017)<br>• PICUs (Ellis et al., 2019; Hardy et al., 2017)<br>• Health-based place of safety room at a psychiatric hospital (Wilson et al., 2023)<br><br>Hakimzada et al. (2020) explored perceptions of BWCs amongst staff on two acute wards, four secure wards and a PICU which had <u>not</u> yet implemented BWCs. | wearer; staff wearing the cameras will clearly let people know when they begin any recording; cameras will be activated if staff believe that safety may be compromised when responding to incidents; and all recorded data will be processed in accordance with the Data Protection Act 98. The posters were regularly replaced if removed by patients. Staff verbally informed patients about the cameras by including prompts in morning meetings, patient experience groups and community meetings.<br><br>In Hardy et al.'s (2017) study, 68% of patients surveyed reported that they had been made aware that some of the nurses were wearing BWCs on the ward. The patients who reported that they had not been made aware were from three of the wards with half of these being from one ward. Methods of being informed about BWCs identified by patients (n = 39):<br>• Posters: 20 (51%)<br>• Informed on admission: 5 (13%)<br>• Informed at morning meeting: 15 (38%)<br>• Given written information: 5 (13%) |

| | | | | |
|---|---|---|---|---|
| | Prevention and Management of Violence and Aggression (PMVA) team, and the night manager).<br><br><u>Training and preparation:</u><br>In Hardy et al.'s (2017) study, before implementation of BWCs, the necessary policies, IT infrastructure and information governance compliance were established. A full privacy impact assessment and self-assessment tool from the surveillance camera commissioner were completed. Patients and visitors were informed. A 90-minute training was delivered to staff by Reveal trading as Calla (the BWC supplier). Aspects covered included: teaching about the purpose of BWCs, how they work; practical aspects of collecting, using and returning the camera; data security how data is stored and how to search for footage; and how to send footage to the police if necessary. Ward managers and/or a member of the PMVA team cascaded this training to ward staff. Members of the PMVA team were trained to act as administrators by Reveal trading as Calla.<br><br><u>Stated aims/functions:</u><br>Intended functions of BWCs in inpatient mental health settings include: increasing transparency; resolving incidents and complaints by providing accurate incident records; improving staff performance by using footage for training and monitoring; and improving staff conduct and the behaviour of patients being recorded and preventing confrontational situations, reducing incidents of aggression, improving safety and countering false allegations (Ellis et al., 2019; Hakimzada et al., 2020; Hardy et al., 2017; Wilson et al., 2023). | | • Other: 1 (3%) | |
| GPS electronic monitoring<br><br>(Murphy et al., 2017; Tully et al., 2016) | In medium-secure forensic inpatient mental health settings, GPS monitoring has been implemented to monitor the whereabouts of patients on leave (Murphy et al., 2017; Tully et al., 2016). This includes monitoring leave violations, such as absconding and failure to return (Murphy et al., 2017). | <u>Countries:</u><br>Both UK-based<br><br><u>Ward types:</u> | Both studies stated that wearing the device was voluntary and only used with patients who consented (Murphy et al,. 2017; Tully et al., 2016), but an exception was made for high-risk patients requiring | Tully et al. (2016) states that the introduction of the technology was discussed with patients and legal advisors, and consent and information forms were developed. No further detail was provided. |

| | | | |
|---|---|---|---|
| (2 papers)<br><br><u>Brands :</u><br>• Buddi Tracker (Tully et al., 2016)<br>• Unspecified (Murphy et al. 2017) | Tully et al. (2016) stated that it was primarily intended to be used with patients in the early stages of leave, when risk of leave violation is highest. Murphy et al. [50] stated that clinical decisions about the appropriateness of GPS electronic monitoring for individual patients are made following a specific risk assessment protocol. Both studies stated that the device was only used with patients who consented to its use. In Murphy et al. (2017), an exception was high-risk patients who required urgent hospital or court transfer.<br><br>In both studies, the devices were attached to patients' ankles. In Tully et al. (2016), each patient had their own allocated 'Buddi Tracker' device, which could be attached to their ankle using an individually measured secure strap whilst on leave. The strap incorporates cabling to make it non-removable, and optic fibres to provide anti-tamper alarms. The device works anywhere that a GPS signal can be received, and its location is determined by a series of signals routed via satellites. The device transmits its location periodically to the monitoring software via a mobile phone network. Geographical parameters, known as 'geo-fences', can be set enabling the creation of exclusion and inclusion zones (a common sanction for forensic patients). Should a wearer breach a geofence, an alarm is created which vibrates the device on the ankle. An alert signal is also sent through the in-built monitoring software. Information from each device is monitored by a security company and breaches of agreed terms and conditions trigger a predetermined alert to relevant parties and a risk management plan (Tully et al,. 2016).<br><br><u>Intended aims/functions:</u><br>Described as being used to monitor the whereabouts of patients on leave. Murphy et al. (2017) hypothesised that the technology would reduce leave violations, increase overall leave, and increase the proportion of unescorted leave. | Both based in medium-secure inpatient mental health settings. | urgent hospital or court transfer in Murphy et al. (2017).<br>Information about this consent process was not provided.<br>Consent rates were not provided. |

| | | | | |
|---|---|---|---|---|
| Wearable sensors<br><br>(Greer et al., 2019; Tron et al., 2018)<br><br>(2 papers)<br><br><u>Brands:</u><br>• GeneActiv (Tron et al., 2018)<br>• E4 (Empatica Srl) and Everion (Biovotion Ltd) (Greer et al., 2019) | In Tron et al.'s (2018) study, psychiatric inpatients with psychosis wore GeneActiv smartwatches equipped with accelerometers, light and temperature sensors. Their high frequency output (50Hz) was stored on memory cards in the device. Medical staff managed the placement and removal of these devices and uploaded the data from the memory card to a central storage location for analysis.<br><br>Greer et al.'s (2019) study explored staff perceptions of two remote monitoring devices – E4 (worn around the wrist) and Everion (worn around the upper arm). It explored their views of using them to monitor patients' physiological signals in real-time to predict aggression in inpatient settings. No further detail was provided about how these devices operate.<br><br><u>Intended aims/functions:</u><br>In Tron et al.'s (2018) study, GeneActiv's purpose was to monitor high-level movement parameters, correlate them with patients' mental state and facilitate continuous motor deficits tracking. They stated that they hoped this could help to develop an automatic system for evaluating symptom severity in schizophrenia, to characterise schizophrenia subtypes, gain insights into underlying causes, and enable more personalised treatment.<br><br>In Greer et al. (2019) the intended aims of the E4 and Everion devices were to monitor patients' physiological signals in real-time to predict aggression in inpatient settings. | <u>Countries:</u><br>• Israel [Tron et al,. 2018)<br>• UK (Greer et al., 2019)<br><br><u>Ward types:</u><br>• "Psychiatric inpatient facility" (Tron et al., 2018).<br>• Medium secure inpatient ward (Greer et al (2019) did not actually implement, just explored staff views on the devices. | Tron et al. (2018) stated that patients had to consent to participate in the study, but no further detail was provided about the consent process. | None reported. |

Acronyms: BWCs = Body Worn Cameras; CCTV = Closed Circuit Television; EM = Electronic Monitoring; GPS = Global Positioning System; IT = Information Technology; MMAT = Mixed Methods Appraisal Tool; NHS = National Health Service; PICU = Psychiatric Intensive Care Unit; PIN = Personal Identification Number; PMVA = Prevention and Management of Violence and Aggression; TV = Television; UK = United Kingdom; USA = United States of America; VBPMM = Vision-Based Patient Monitoring and Management.
